# Vaccine optimization for COVID-19: who to vaccinate first?

**DOI:** 10.1101/2020.08.14.20175257

**Authors:** Laura Matrajt, Julia Eaton, Tiffany Leung, Elizabeth R. Brown

## Abstract

A vaccine, when available, will likely become our best tool to control the current COVID-19 pandemic. Even in the most optimistic scenarios, vaccine shortages will likely occur. Using an age-stratified mathematical model paired with optimization algorithms, we determined optimal vaccine allocation for four different metrics (deaths, symptomatic infections, and maximum non-ICU and ICU hospitalizations) under many scenarios. We find that a vaccine with effectiveness ≥50% would be enough to substantially mitigate the ongoing pandemic provided that a high percentage of the population is optimally vaccinated. When minimizing deaths, we find that for low vaccine effectiveness, irrespective of vaccination coverage, it is optimal to allocate vaccine to high-risk (older) age-groups first. In contrast, for higher vaccine effectiveness, there is a switch to allocate vaccine to high-transmission (younger) age-groups first for high vaccination coverage. While there are other societal and ethical considerations, this work can provide an evidence-based rationale for vaccine prioritization.

## Introduction

As of 22 September 2020, over 960 thousand people have died due to the ongoing SARS-CoV-2 pandemic (*1*). Different countries have enacted different containment and mitigation strategies, but the world awaits impatiently for the arrival of a vaccine as the ultimate tool to fight this disease and to allow us to resume our normal activities. There are over 100 vaccines under development (*2, 3*), with some currently undergoing phase 3 clinical trials (*3*). However, there are many unknowns surrounding a potential vaccine, including how effective it would be, how long it would be protective, how effective it would be in older individuals, how many doses would be immediately available, and how long scaling up the vaccine production would take. Furthermore, should early vaccines have low effectiveness, what are the potential trade-offs between using a low-effectiveness vaccine and waiting for a vaccine with a more desirable vaccine effectiveness? With the hope of producing a vaccine in the near future comes the difficult task of deciding whom to vaccinate first as vaccine shortages are inevitable (*4–6*). Here we utilized a mathematical model paired with optimization algorithms to determine the optimal use of vaccine for 100 combinations of vaccine effectiveness (VE) and number of doses available under a wide variety of scenarios.

## Methods

Briefly, we developed a deterministic age-structured mathematical model of SARS-CoV-2 transmission with a population stratified into 16 age groups (Fig. S1, SM). Because, historically, vaccine is distributed to each state in the United States (US) proportional to its population, and the allocation strategy is then determined at the state level (*7*), we chose a state level model with a population size similar to Washington State and demographics similar to those to the general US population; however, our results are generalizable to other populations.

We assumed that children were less susceptible to infection than middle-aged adults (20 to 65 years old), while older adults (older than 65) were relatively more susceptible (*8*) (we also analyzed a scenario assuming equal susceptibility across age groups, see SM). We assumed that both natural and vaccine-induced immunity last at least one year (our time horizon). At the beginning of our simulations, 20% of the population have already been infected and are immune (additional results for 10, 30 and 40% of the population can be found in the Results section and in the SM) and all social distancing interventions have been lifted. To keep our results as general as possible, as each state/country will have different vaccination rates, we did not model the vaccination campaigns in the main analysis. Hence, we assumed that at the beginning of our simulations, vaccination has been carried out and that vaccinated individuals have reached the full protection conferred by the vaccine. However, we also analyzed, in a separate scenario, how optimal allocation strategies changed when we modeled vaccination campaigns explicitly. Here, we consider that front-line health care workers and other essential personnel (e.g. firefighters, police) who should obviously be prioritized, have already been vaccinated.

For the main analysis, we considered a vaccine having an effect of reducing susceptibility to infection (referred to VE throughout the text) only. In addition, as separate analysis, we considered a vaccine that would also reduce the probability of COVID-19 disease (referred as VE*_COV_* below). This effect against COVID-19 disease was modeled as a combination of the vaccine effectiveness to prevent infection and the vaccine effectiveness to prevent disease given infection. Because current vaccine clinical trial protocols have an expected vaccine effectiveness against COVID-19 disease of 60% (*9, 10*), for this analysis we set VE_*COV*_ =60 while varying the relative contributions of the other two vaccine effects (see SM for full description).

For the vaccine optimization, we collated the 16 age groups into five vaccination groups: children (aged 0–19), adults between 20 and 49 years old, adults between 50 and 64 years old, adults between 65 and 74 years old, and those 75 and older. This stratification reflects our current knowledge of disease severity and mortality based on age (*11, 12*).

We developed an optimization routine that combined a coarse global search algorithm with a fast optimizer to explore the entire space of possible combinations of vaccine allocation. We compared the optimal allocation strategy given by the optimizer to a pro-rata allocation, where the vaccination coverage to each vaccination group is distributed proportionally to population size in each group. We considered VE ranging from 10% to 100% and vaccination coverage ranging from 10% to 100% of the total population. We evaluated four objective functions reflecting different metrics of disease burden that could be considered by decision makers: minimization of the total number of symptomatic infections, total number of deaths, number of cases requiring hospitalization (non-ICU) at the epidemic peak, and number of cases requiring ICU hospitalization at the epidemic peak. We chose to minimize symptomatic infections as a key metric because symptomatic individuals are the ones who are easier to identify and presumably, particular interventions will be targeted to this group. In addition, minimizing symptomatic individuals minimizes the transmission of SARS-CoV-2, and this is in line with current vaccine trials endpoints (*9, 10*). The last two objective functions were chosen because hospital bed (non-ICU and ICU) occupancy is a key metric currently used to determine county/state/country readiness to move between different interventions strategies. Here, we utilized the total number of licensed ICU beds in WA state and its current goal of staying below 10% of hospital beds occupied by COVID-19 cases (*13, 14*) as references when interpreting our results. Full details of the methods can be found in the SM.

## Results

### Epidemic mitigation and containment

Our model suggests that, for a basic reproduction number *R*_0_ = 3, herd immunity will be achieved once 60% of the population is infected (equivalently 40% vaccinated with a perfect vaccine under the optimal allocation strategy for minimizing infections assuming 20% of the population has immunity already) (Fig. 1J, Fig. S2 and Fig. 2A).

**Figure 1:**
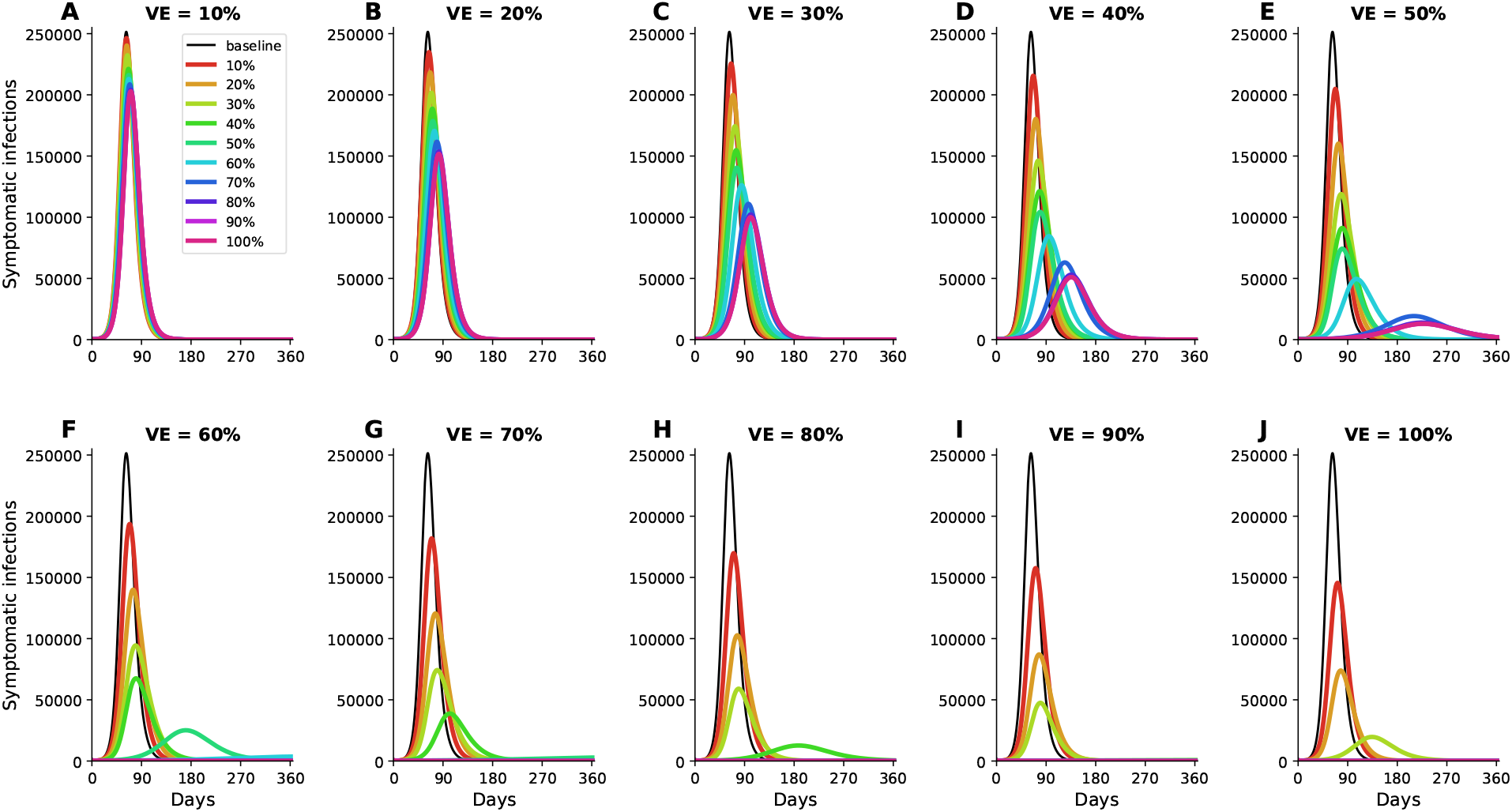
Simulated prevalence of symptomatic COVID-19 infections for VE ranging from 10% (A) to 100% (J) in 10% increments. For each VE and each vaccination coverage, the optimal vaccine allocation for minimizing symptomatic infections was used in these simulations. Colors represent different vaccination coverage, ranging from 0 (black, “baseline”) to 100% (magenta). For clarity, we present here epidemic curves for the main set of parameters only and show a complete figure with uncertainty bounds in Fig. S2.

**Figure 2:**
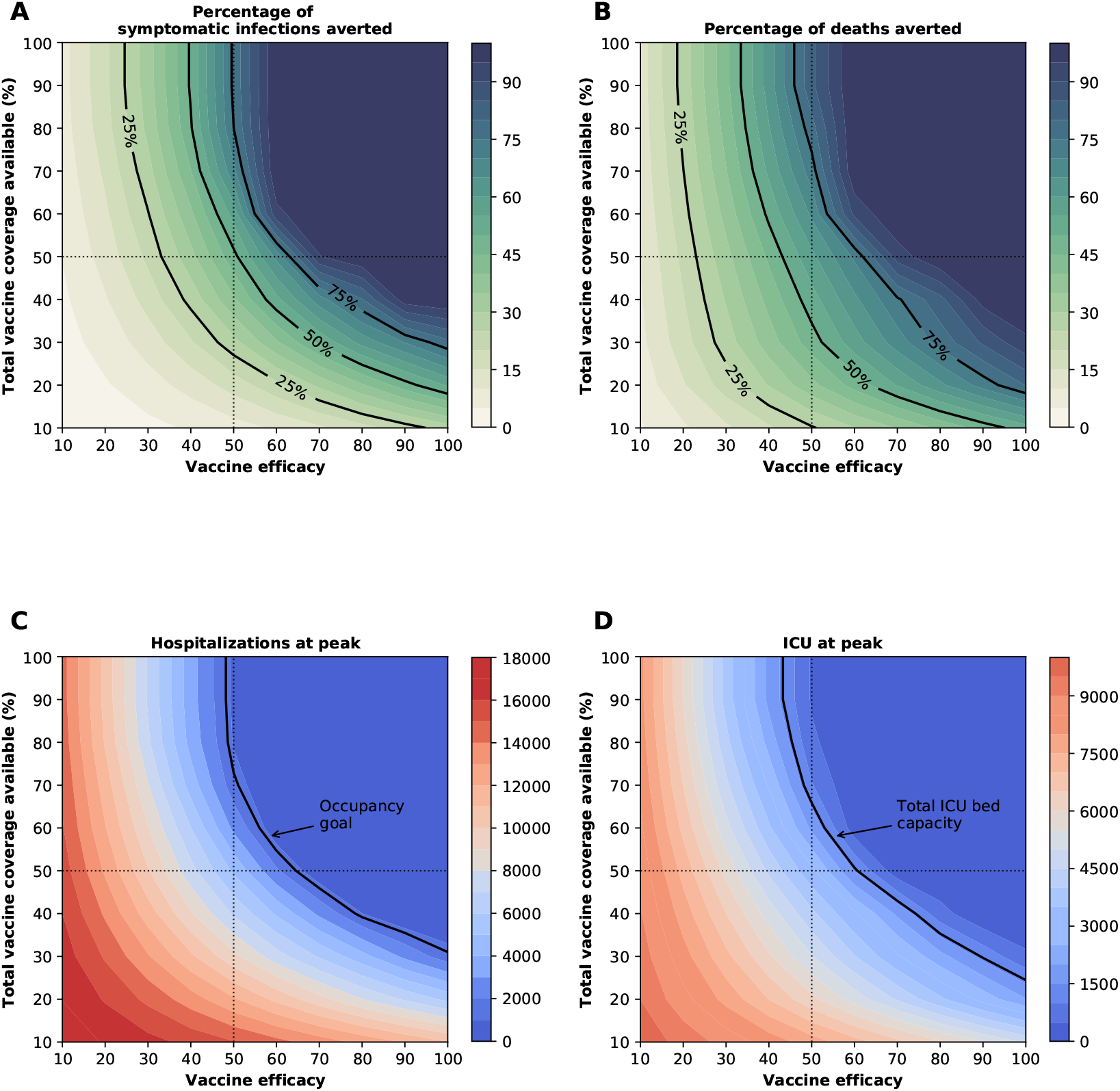
Four key metrics of COVID-19 burden under optimal distribution of vaccine. Percentage of symptomatic infections (A) and deaths (B) averted, number of maximum non-ICU (C) and ICU (D) hospitalizations as a function of VE and vaccination coverage (total vaccine available as a percentage of the population). The dotted lines correspond to VE = 50% and vaccine available to cover 50% of the population. The isoclines indicate the current goal for WA state of having 10% of licensed general (non-ICU) hospital beds occupied by COVID-19 patients in (C) and total ICU licensed hospital beds in WA state in (D).

The epidemic can be substantially slowed with any vaccine with a VE ≥ 50% as long as a majority of the population is vaccinated (Fig. 1E, Fig. 2A), and over 50% of deaths could be averted (in comparison to no vaccination and no non-pharmaceutical intervention) with as little as 35% of the population optimally vaccinated (Fig. 2A, B). If VE = 60%, the epidemic is completely contained if we optimally vaccinate 70% of the population (or 50% for higher VE = 70%) (Fig. 2A and Fig. 1F, I). In our model, only vaccines with VE ≥50% can maintain the number of non-ICU hospitalizations below the established goal (≤10% hospital-bed occupancy by COVID-19 patients) and can prevent an overflow of the ICUs. With VE = 60%, optimally vaccinating 54% satisfies both hospital-bed occupancy goals (Figs. 2C, D, S3F and S4F), compared to 67% under the pro-rata allocation (Figs. S5–S9). The optimal allocation strategy outperforms the pro-rata allocation most under low vaccine availability, with a maximum difference of 32% deaths averted (for VE = 100% and with enough vaccine to cover 20% of the population) and 32% symptomatic infections averted (for VE = 60% and vaccination coverage of 60%) when compared with a pro-rata allocation strategy (Figs. 3, S5, S10). As VE increases, both strategies tend to perform similarly as vaccination coverage increases (Fig. 3, S5 S10).

**Figure 3:**
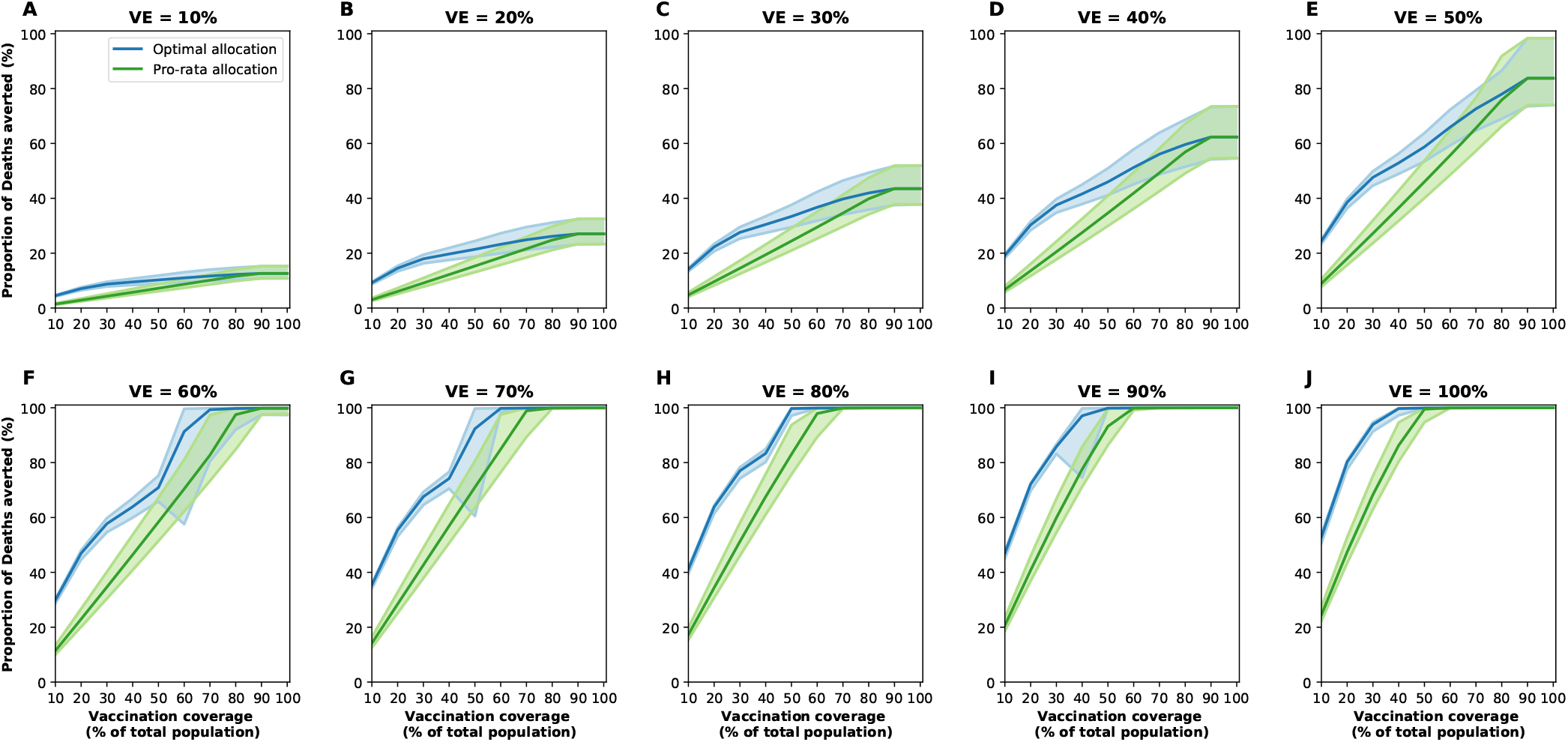
Percentage of deaths averted for the optimal allocation strategy (blue) and the pro-rata strategy (green) for VE ranging from 10% (A) to 100% (J) in 10% increments and vaccination coverage ranging from 10% to 100% of the total population. The shaded areas represent results of 1,000 simulations with the top and bottom 2.5% simulations removed.

### Optimal vaccine allocation changes with VE and vaccination coverage

The optimal allocation strategy to minimize deaths is identical for vaccine effectiveness between 10% and 50%: with low vaccination coverage, it is optimal to allocate vaccine first to the highest risk group (people over 75 years old) and then to the younger vaccination groups as more vaccine becomes available (Fig. 4A–E). In contrast, there is a threshold phenomenon observed for VE≥60%: for low coverage, the optimal allocation is still to vaccinate the high-risk groups first, but when there is enough vaccine to cover roughly half of the population (60% for VE = 60%, 50% for VE = 70%, and 40% for VE≥ 80%), there is a switch to allocate vaccine to the high-transmission groups (those aged 20-50 and children in our model) first. This is because directly vaccinating those who are driving the epidemic results in a much slower epidemic curve and hence in fewer deaths (Fig. 1F–H). As more vaccine becomes available the optimizer allocates it to high-risk groups again (Fig. 4F–J).

**Figure 4:**
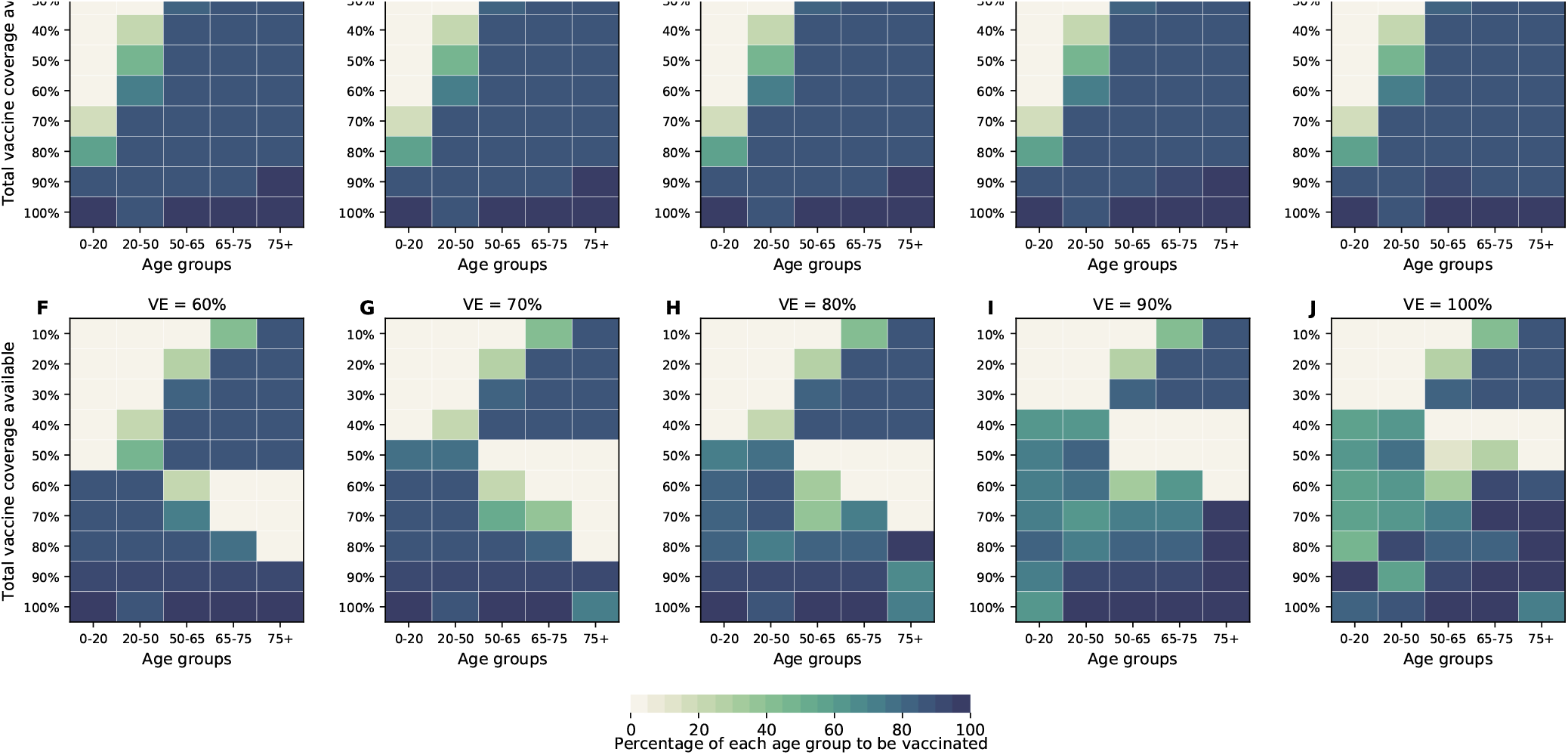
Optimal allocation strategies for minimizing deaths for VE ranging from 10% (A) to 100% (J) in 10% increments (additional figures for minimizing symptomatic infections, number of non-ICU hospitalizations at peak and number of ICU hospitalizations at peak are given in SM). For each plot, each row represents the total vaccination coverage available (percentage of the total population to be vaccinated) and each column represents a different vaccination group. Colors represent the percentage of the population in a given vaccination group to be vaccinated.

### Optimal vaccine allocation differs for different objective functions

Next, we investigated how the optimal allocation strategy changed for different objective functions and present results for VE = 60%. The optimal vaccine allocation for the four objectives differed the most when fewer vaccines are available (enough vaccine to cover less than 30% of the total population). When minimizing symptomatic infections and peak non-ICU hospitalizations, priority was given to the younger vaccination groups, as they have the most contacts in our model and hence drive transmission (Fig. 5A,B, Fig. S11, S12). As we move toward more severe outcomes (ICU hospitalizations at peak and deaths), for which older individuals are most at risk, the optimal allocation strategy shifts toward those vaccination groups (Fig. 5C,D, S13). Once more vaccine becomes available, the optimal allocation strategies are very similar for all objective functions. In fact, they are nearly identical for all the objective functions when there is enough vaccine to cover 60% and 70% of the population. For high coverage, the optimal allocation strategies for all objective functions shifted towards the high-transmission groups. To note, we did not impose the optimizer to use all the available vaccine. As a result, the optimizer found allocation strategies utilizing less than the total vaccine available while performing equally well. This was very prominent when VE and vaccination coverage were very high. For example, when minimizing peak ICU hospitalizations and VE = 90%, the optimizer utilized vaccine to cover 75% of the population even though there was vaccine available to cover the entire population. This is expected, complete containment is attained once a high proportion of the population is vaccinated and any vaccine used above that threshold will result in the same mathematical outcome.

**Figure 5:**
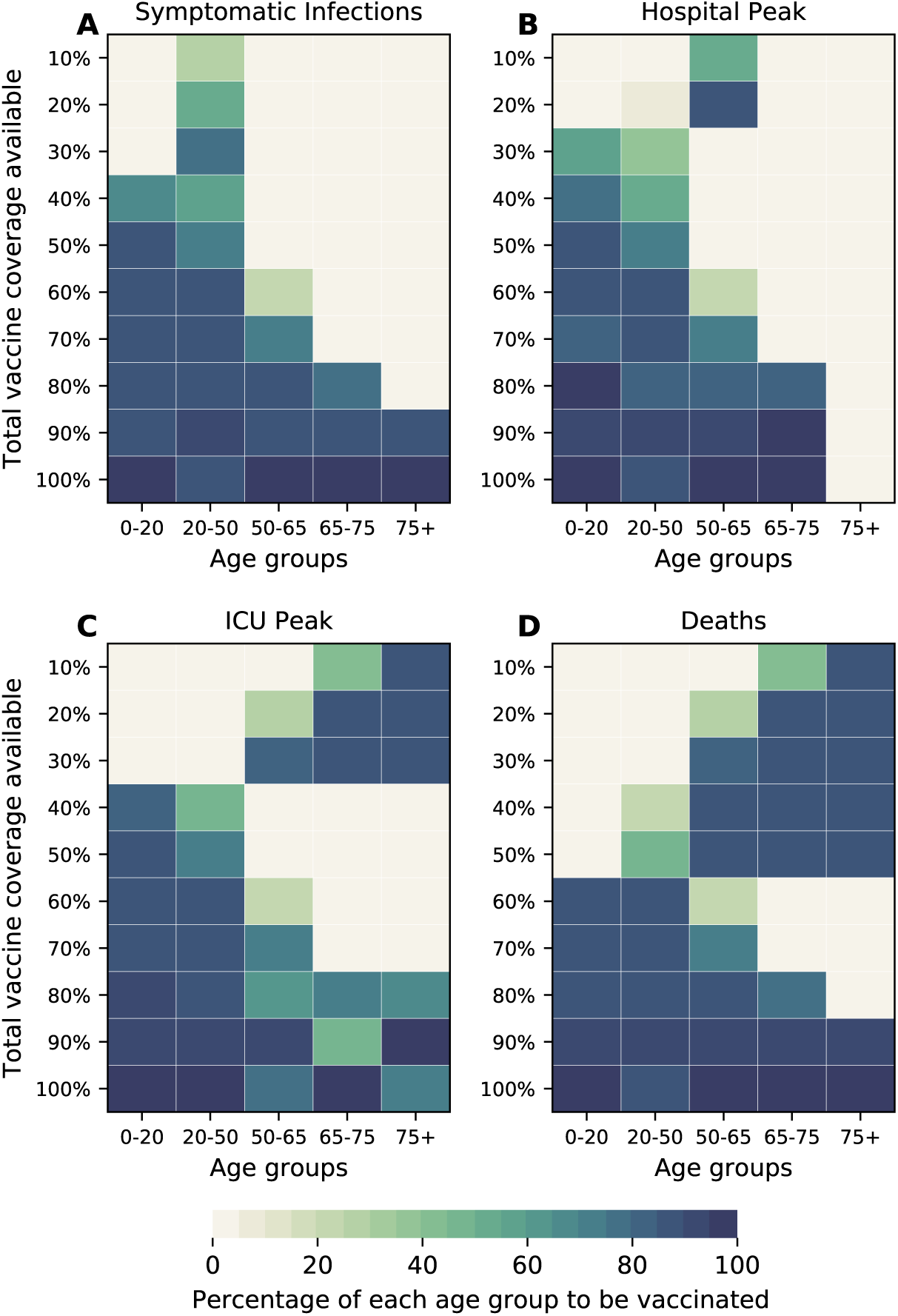
Optimal allocation strategies for minimizing: symptomatic infections (A), Number of non-ICU hospitalizations at peak (B), Number of ICU hospitalizations at peak (C) and total number of deaths (D). Here, we assumed VE = 60%. For each plot, each row represents the total vaccination coverage available (percentage of the total population to be vaccinated) and each column represents a different vaccination group. Colors represent the percentage of the population in a given vaccination group to be vaccinated.

### Optimal vaccine allocation assuming vaccine effectiveness against COVID-19 disease (VE_*COV*_)

In this section, we considered a vaccine that in addition to reducing the probability of acquiring infection, it would reduce the probability of COVID-19 disease (see SM for full details). We considered VE_*COV*_ = 60%, in line with expected vaccine effectiveness in the current trial protocols (*9, 10*). The optimal allocation strategies assuming VE_*COV*_ = 60% are nearly identical to the ones without this effect (Fig. 6, S14-S16). As expected, including this effect against COVID-19 disease has a huge impact on symptomatic infections and hospitalizations, even when this vaccine is marginally effective against preventing infection (Fig. 7). For example, if VE = 10% it can prevent 50% of the symptomatic infections when we optimally vaccinate 64% of the population (Fig. 7A). Further, for this VE and this vaccination coverage, peak hospitalizations are substantially reduced (6,840 and 3,392 non-ICU and ICU hospitalizations respectively; Fig. 7B, C) when compared to a vaccine without an effect in COVID-19 disease (15,091 and 8,405 non-ICU and ICU hospitalizations respectively; Fig. 2B, C).

**Figure 6:**
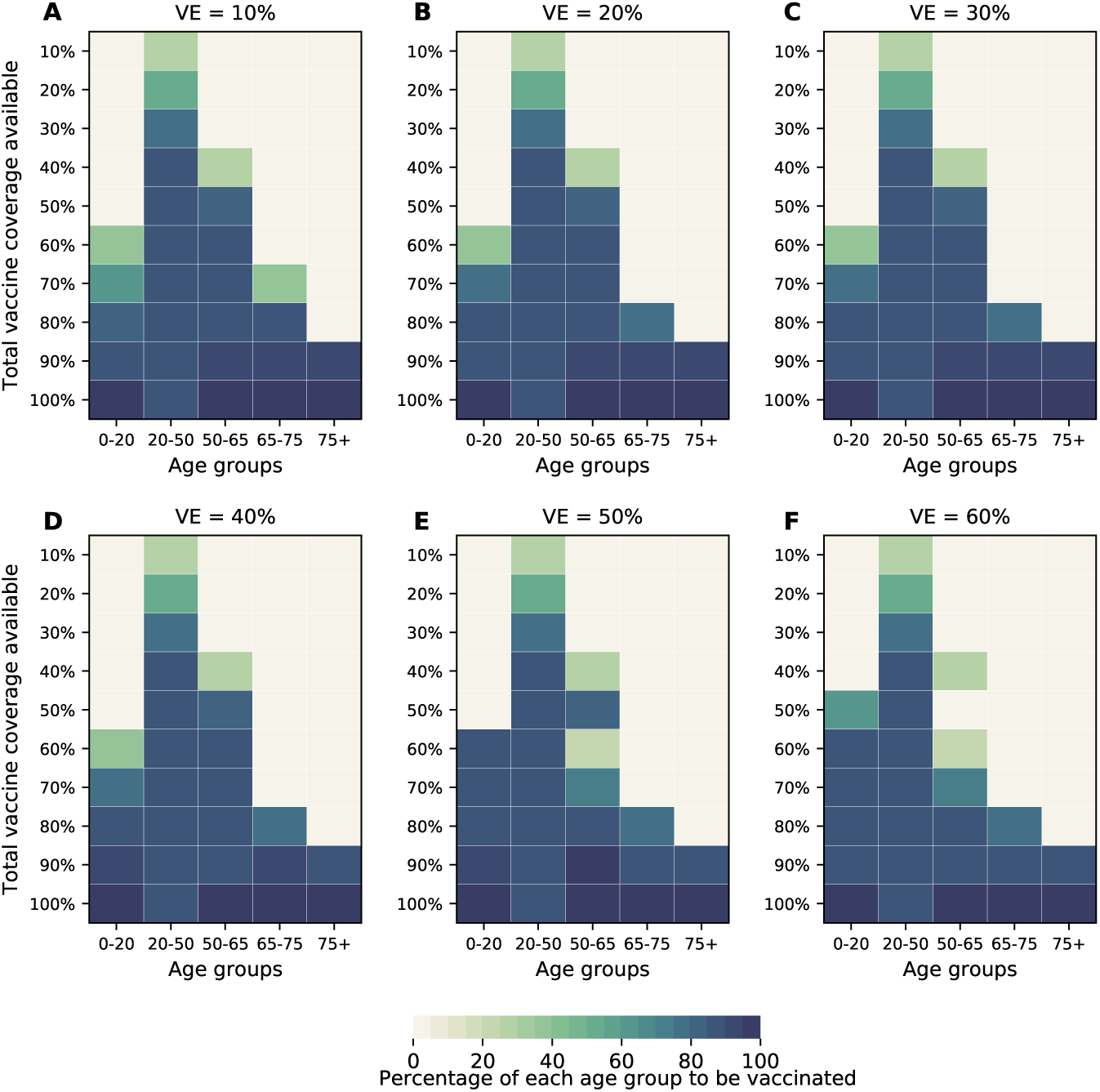
Optimal allocation strategies for minimizing total symptomatic infections for VE ranging from 10% (A) to 60% (F) in 10% increments for VE_*COV*_ = 60%. For each plot, each row represents the total vaccination coverage available (percentage of the total population to be vaccinated) and each column represents a different vaccination group. Colors represent the percentage of the population in a given vaccination group to be vaccinated.

**Figure 7:**
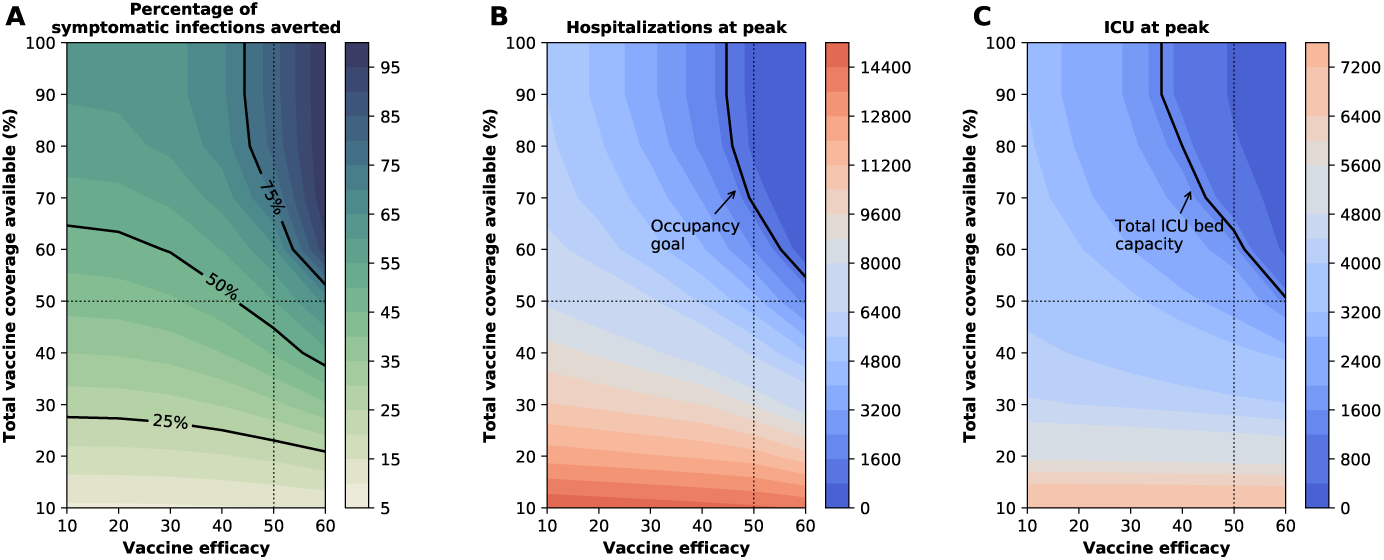
Three key metrics of COVID-19 burden under optimal distribution of vaccine for VE_*COV*_ = 60%. Percentage of symptomatic infections averted (A), number of maximum non-ICU (C) and ICU (D) hospitalizations as a function of VE and vaccination coverage (total vaccine available as a percentage of the population). The dotted lines correspond to VE = 50% and vaccine available to cover 50% of the population. The isoclines indicate the current goal for WA state of having 10% of licensed general (non-ICU) hospital beds occupied by COVID-19 patients in (C) and total ICU licensed hospital beds in WA state in (D).

### Optimal vaccine allocation as a function of pre-existing immunity to SARS-CoV-2

As the COVID-19 pandemic dynamics have been dramatically different across the globe, we expect to see a range of population-level naturally-acquired immunity when vaccination campaigns start. Hence, we investigated the optimal use of vaccine with 10%, 30%, and 40% of the population already immune at the beginning of the simulations. For all of these, the same pattern is observed when minimizing deaths: for low coverage, it is optimal to allocate all of the vaccine to the high-risk groups; for higher coverage, the optimal vaccination strategy switches to allocate more vaccine to the high-transmission groups. This threshold however varies with the degrees of pre-existing immunity in the population. When only 10% of the population has natural immunity, the switch occurs at 80% vaccination coverage, but when 40% has natural immunity, the threshold is observed at 40% vaccination coverage (Fig. 8). In addition, under low pre-existing immunity to SARS-CoV-2, the optimal strategy favors vaccinating the older vaccination groups (Fig. 8A), while under higher pre-existing immunity, the optimal allocation strategy tends to distribute vaccine more evenly across vaccination groups (Fig. 8D).

**Figure 8:**
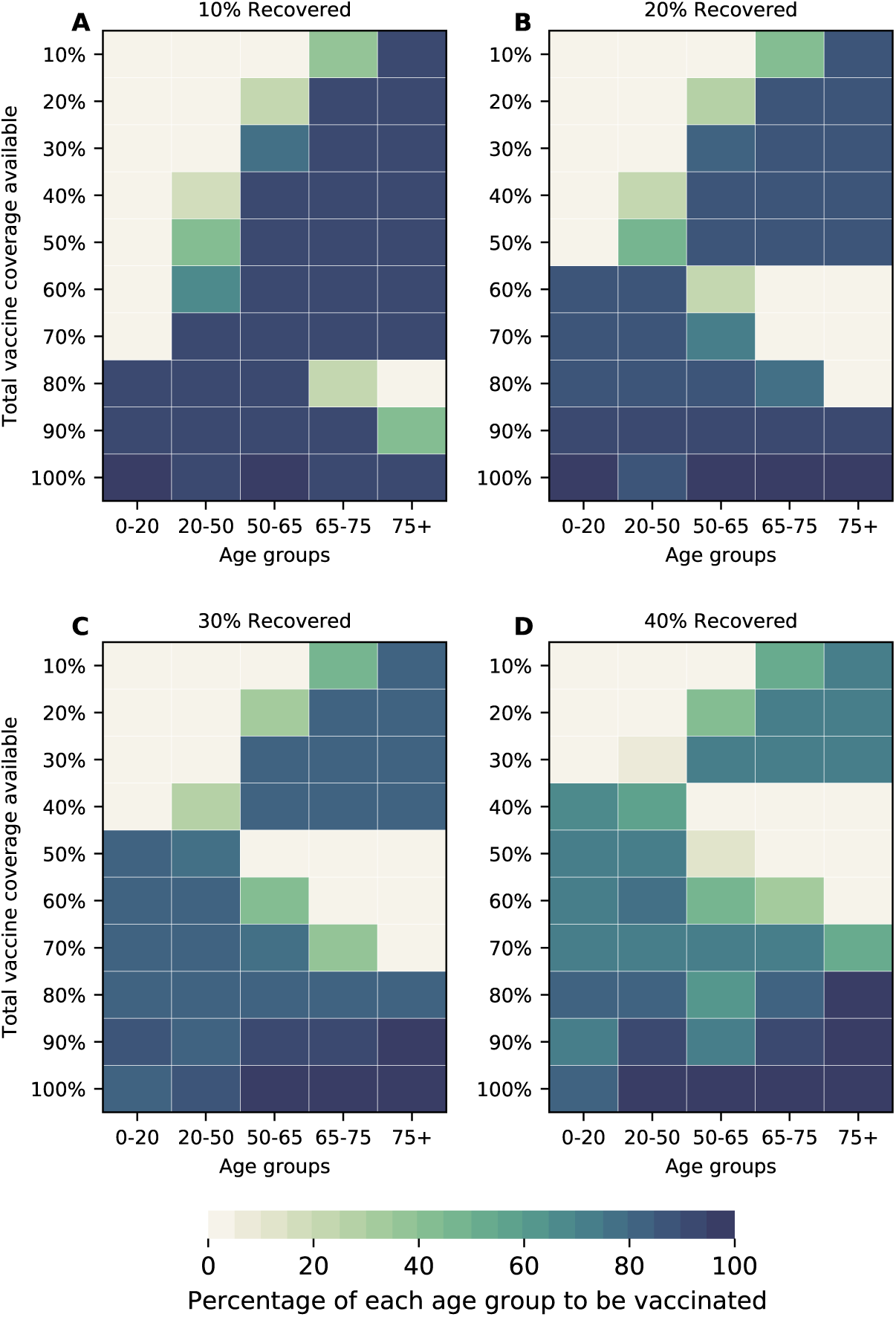
Optimal allocation strategies for minimizing deaths assuming 10% (A), 20%(B) 30% (C) and 40% (D) of the population has natural immunity to COVID-19 at the start of the simulations. Here, we assumed VE = 60%. For each plot, each row represents the total vaccination coverage available (percentage of the total population to be vaccinated) and each column represents a different vaccination group. Colors represent the percentage of the population in a given vaccination group to be vaccinated.

### Modeling the vaccination campaign

In this section we modeled the vaccination campaign and determined the optimal vaccine allocation. We extended our time horizon to two years and considered administering 75, 150, or 300 thousand doses of vaccine per week (denoted by 75K, 150K and 300K respectively). This corresponds to vaccinating the entire population in 101, 50 or 25 weeks respectively. The first vaccination rate was chosen to roughly match the vaccination rate during the 2009 H1N1 pandemic in the US (0.87% of the population weekly (*15*)). It is important to note that in order to avoid confounding, as in the rest of this work, we did not assume any social distancing intervention. Similar to previous results, when the vaccination campaign is modeled, the optimal vaccine allocation strategies were very different depending on the objective function: while the older age groups are prioritized when minimizing deaths and maximum ICU-hospitalizations (Fig. 9), the optimizer allocated vaccine to younger age groups when minimizing symptomatic infections or maximum non-ICU hospitalizations (Fig. S17). Further, older age groups were prioritized when vaccination rate was slow (75K doses administered per week, when minimizing deaths and ICU peak hospitalization). For faster vaccination campaigns, the optimizer allocated vaccine to younger age groups in addition to the older age groups. (Fig. 9). There were also some important differences. First, for any VE and any vaccination rate, we did not observe any threshold in vaccination coverage. In addition, because the time frame of the vaccination campaigns occurs at a much slower speed than the epidemic, most of the vaccine under this scenario is given after the epidemic (this was especially true when considering the 75K campaign). This resulted in optimal allocation strategies that for a given VE, were identical beyond a certain vaccination coverage (Fig. 9, Fig. S17) and the measured outcomes did not improve beyond that coverage (Fig. S18). This points to the fact that even in the very optimistic scenario of having a highly efficacious vaccine given at very high rates, additional interventions would be necessary to control the epidemic while the vaccination campaign takes place.

**Figure 9:**
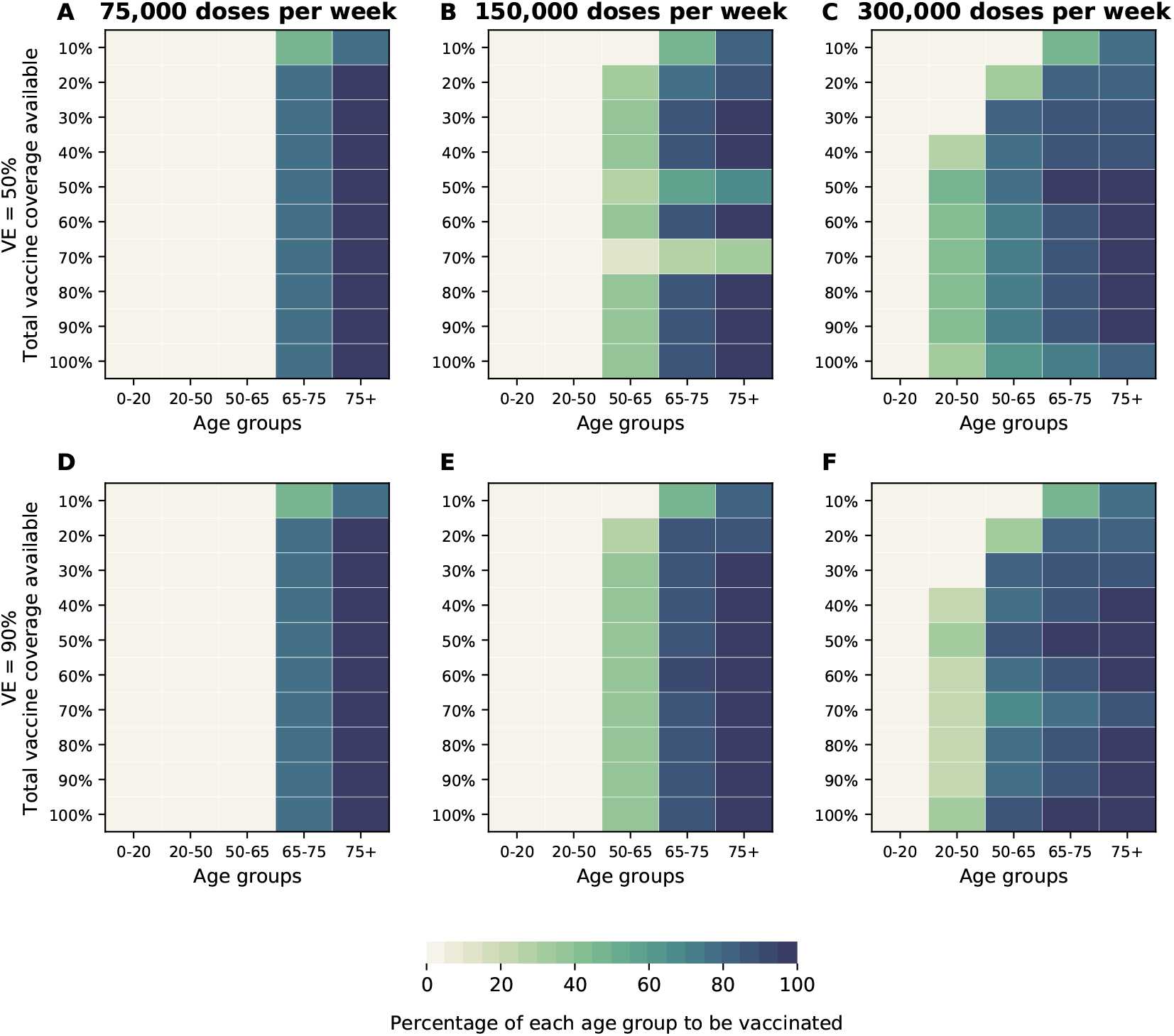
Optimal allocation strategies for minimizing deaths for two VE = 50% (A-C) and VE = 90% (D-F) and for three different vaccination rates: 75K (A and D), 150 (B and E) and 300 (C and F) thousand vaccine doses administered per week. For each plot, each row represents the total vaccination coverage available (percentage of the total population to be vaccinated) and each column represents a different vaccination group. Colors represent the percentage of the population in a given vaccination group to be vaccinated.

### Robustness of optimal allocation strategies around major parameters

#### One-way robustness analysis

We explored the robustness of the optimal allocation strategies around key features of the transmission and natural history of SARS-CoV-2. For each of these features, we investigated how changing that particular parameter would change the optimal allocation strategy.

##### Susceptibility to infection

Because the effect of age on susceptibility to SARS-CoV-2 infection remains unclear, we compared the optimal allocation strategy under the assumption of differential susceptibility, as suggested in (*8, 16*) (presented throughout the text), to one assuming equal susceptibility across age groups (Figs. S19–S22), as suggested in (*17, 18*). The optimal allocation strategies under both equal and differential susceptibility were remarkably consistent, but as expected, assuming equal susceptibility resulted in strategies allocating slightly more vaccine to children (assuming equal susceptibility) as VE increases (VE≥ 60) and more vaccine becomes available (coverage to vaccinate 70% or higher) (Figs. S19–S22). The major differences were observed when minimizing peak non-ICU hospitalizations for low VE and low vaccination coverage (less than, assuming equal susceptibility resulted in favoring adults aged 50-65 over younger adults (Fig. S21).

##### Susceptibility to symptomatic disease

While we know that children are much less susceptible to develop severe disease (*19*), the role of age in the probability of developing any kind of COVID-19 symptoms remains currently unclear. Hence we compared optimal allocation strategies assuming assuming equal probability of all age groups to develop symptoms (presented throughout the text) as suggested in (*19, 20*) to ones assuming different probabilities by age (children being less susceptible and older adults, more susceptible), as suggested in (*21*). The optimal allocation strategies for minimizing deaths and peak ICU are very consistent (Figs. S23, S24), but, as expected, we observed some changes when minimizing symptomatic infections and peak non-ICU hospitalizations (Figs. S25, S26). When we minimized symptomatic infections, the optimal allocation strategies were very similar for vaccines with VE≥ 60. For lower VE, the optimal allocation strategies tended to favor more the middle- and older-age adults groups (Fig. S25). When minimizing maximum non-ICU hospitalizations, the optimal allocation strategies were nearly identical for high vaccination coverage (≥ 70%) but switched to adults aged 50-75 (as opposed to younger adults) if vaccination coverage is lower (Fig. S26).

##### Basic reproduction number *R*_0_

Many regions in the world have controlled the epidemic using social distancing interventions and have reduced the effective reproduction number *R*_*t*_ to hover around 1. Hence, we analyzed the optimal allocation strategies in this section with *R*_0_ set to 1.5, 2 and 2.5, assuming still that at the beginning of our simulations, 20% of the population has been infected and are now recovered. For minimizing deaths and ICU peak hospitalizations, when *R*_0_ = 1.5, the optimal allocation strategy favors vaccinating children for low VE (Fig. 10A, S27), is more equally distributed if VE = 60% and favors vaccinating the older age groups for higher VE (Fig. 10D, G). If *R*_0_ = 2, we observe the same threshold phenomenon described for higher *R*_0_, but at a lower VE: for VE = 30% the optimal use of vaccine is to allocate it to high-risk groups under low vaccine coverage and to the younger groups once there is enough vaccine to cover 60% of the population. As VE is higher, this threshold moves up (Fig. 10C, G, K). Finally, for *R*_0_ = 2.5, the optimal allocation strategy is identical to the one for *R*_0_ = 3. For minimizing symptomatic infections and non-ICU peak hospitalizations, the optimal allocation strategies were more homogeneous, favoring more the younger age groups in alignment with the results presented in the main text (Fig. S28, S29). It is important to note here that social distancing interventions have kept the effective reproduction number low, but that, if those measures are lifted, then *R*_*t*_ would go up again. Hence, any vaccination program should take this into account.

**Figure 10:**
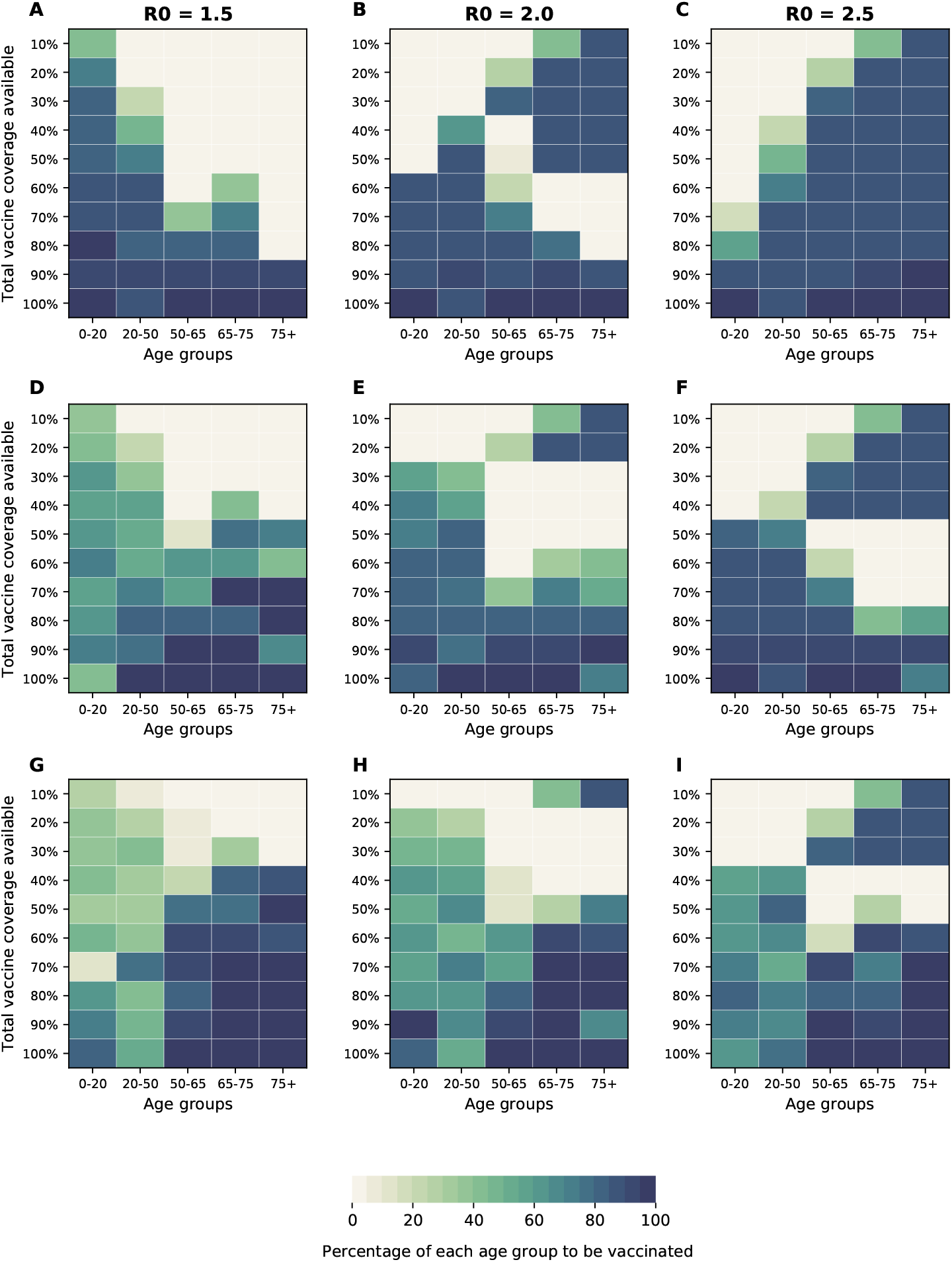
Optimal allocation strategies for minimizing deaths for three different VE: 30% (A-C), 60%(D-F) and 90% (G-I) for three different values of *R*_0_ = 1.5, 2, and 2.5 (additional figures for minimizing symptomatic infections, number of non-ICU hospitalizations at peak and number of ICU hospitalizations at peak are given in SM). For each plot, each row represents the total vaccination coverage available (percentage of the total population to be vaccinated) and each column represents a different vaccination group. Colors represent the percentage of the population in a given vaccination group to be vaccinated.

##### Distribution of pre-existing immunity in the population

Here, we assumed a different Distribution of pre-existing immunity in the population. For this section of the analysis, we ran the simulations without any vaccination until the recovered compartments reached 20% of the population. We then used this composition of the population as the initial distribution of pre-existing immunity (this composition will depend on the contact matrix and the demographics used). The optimal allocation strategies were very similar to those obtained in the main analysis, with some notable differences. First, under this scenario, the optimal allocation strategies tended to protect vaccination groups in full before prioritizing other groups. This was very apparent when minimizing deaths or peak ICU hospitalizations: for low VE (irrespective of vaccination coverage) or high VE but low vaccination coverage, the optimal allocation strategies prioritized the highest-risk age groups (individuals aged 65-75 and those over 75 years old) to get fully before allocating to other groups (Figs. S30, S31). Second, the threshold observed when minimizing deaths was occurred at higher vaccination coverage under this scenario. For example, if VE = 60%, this threshold occurs when there is enough vaccine to cover 70% of the population (Fig. S30F-J). Lastly, when minimizing symptomatic infections or non-ICU peak hospitalizations, the optimal allocation strategies shifted away from young adults towards adults in the 50-65 vaccination groups (Figs. S32, S33).

##### Duration of the incubation period

Based on early studies (*17, 22–24*), we presented results assuming an incubation period of 5.1 days. However, a new study (*25*) has suggested that the incubation period for COVID-19 might be longer (7.76 days). We found no difference in the optimal allocation strategies assuming this longer incubation period (Fig. S34).

##### Number of current infections

We compared the optimal allocation strategies when the simulations were started with a higher number of infected individuals (10,000 current infections). This would reflect a situation where the epidemic is in full exponential growth when vaccination becomes available. The optimal allocation strategy was surprisingly robust under this scenario, with nearly identical allocation strategies for all objective functions (Fig. S35).

#### Multi-way robustness analysis

In addition, we selected four parameters (*R*_0_, proportion of infections that are asymptomatic, relative infectiousness of asymptomatic infections, relative infectiousness of pre-symptomatic infections) for which there is the most uncertainty and re-ran the optimization routine for several combinations of them (full details in SM). The optimal allocation strategies were very robust under this analysis (Supplemental Files SF1–SF4).

## Discussion

The COVID-19 pandemic has devastated families and societies around the world. A vaccine, when available, would most likely become our best tool to control the spread of SARS-CoV-2. However, in the short term, even in the most optimistic scenarios, vaccine production would likely be insufficient. In this work, we paired a mathematical model of SARS-CoV-2 transmission with optimization algorithms to determine optimal vaccine allocation strategies. Given the current uncertainties surrounding such a vaccine (we do not yet know if and when this vaccine would be available, how efficacious it will be, or the number of doses immediately available) we explored 100 combinations of VE and vaccination coverage under a wide variety of scenarios minimizing four metrics of disease burden.

Our results suggest that, assuming *R*_0_ = 3, any vaccine with medium to high effectiveness (VE ≥ 50%) would be able to considerably slow the epidemic while keeping the burden on healthcare systems manageable, as long as a high proportion of the population is optimally vaccinated. Moreover, once VE = 70%, full containment of the epidemic would be possible. This is in agreement with vaccine modeling studies (*26, 27*). Further, we showed that much can be achieved even with low vaccination coverage; indeed, with medium VE, over half of deaths can be averted by optimally vaccinating only 35% of the population. When minimizing deaths, for low VE and a low supply of vaccine, our results suggest that vaccines should be given to the high-risk groups first. For high VE and high vaccination coverage, the optimal allocation strategy switched to vaccinating the high-transmission groups (younger adults and children). This remained true under equal or reduced susceptibility to infection for children, pointing to the importance of children as key players in disease transmission. This finding is consistent with previous work for other respiratory viruses (*28–30*) that found that protecting the high-transmission groups indirectly protects the high-risk groups and is the optimal use of resources. Further, the optimal allocation strategies were identical when we considered a vaccine that would also reduce symptomatic infections, but the impact of such a vaccine would of course be greater in reducing COVID-19 disease and healthcare burden. Our results show that even if this vaccine had a marginal effect in preventing infection, it would still be very beneficial to reduce the number of hospitalizations and symptomatic infections. It is expected that once a vaccine is proven to be effective, more information about its mechanisms of action, including how it affects the viral load trajectory and the relationship between that trajectory and infectiousness will be available allowing us to expand and refine our projections.

Here, we utilized mathematical optimization to determine the optimal vaccine allocation and by design, did not impose any restrictions in the allocation strategies. However in practice, implementation of optimal strategies must also account for other factors (ethical, political, and societal). When large quantities of vaccine are available, a feasible solution could involve first vaccinating the high-risk groups and then allocating the remaining vaccine to the high-transmission groups.

This study has several limitations. Our model assumes that both natural and vaccine-acquired immunity will last for at least one year. We do not yet know how long immunity against SARS-CoV-2 will last. There is some evidence that neutralizing antibodies become undetectable a few weeks following infection (***?***), though it is unclear how this correlates with immunity. If immunity were short-lived, then these results would only be applicable for that duration. Further, we assumed that asymptomatic and symptomatic infections would confer equal immunity. However, it is conceivable that asymptomatic infections might result in a weaker immune response (*31*). We chose four metrics of disease burden to minimize. However, other metrics, such as minimization of asymptomatic infections, or a combination of all of these metrics, might be key to stop the spread of the epidemic. We have identified optimal allocation strategies, and once more information about a vaccine characteristics is known, validating our allocation strategies with more complex models is welcome. To avoid confounding effects from different interventions, we optimized vaccine allocation assuming no social distancing interventions in place. In reality, vaccination, at least at the beginning, would take place while some social distancing interventions remain in effect. Under those circumstances, we would need less vaccine to control the epidemic. In that sense, our results are conservative. To keep the optimization from being unreasonably long, our model assumes that vaccination is given all at once and does not capture geographical differences or other heterogeneities.

We utilized mortality and hospitalization rates published by Ferguson et al. (*12*) that were based on the epidemic in Wuhan, China, but these rates may vary vastly in different regions. In particular, it is now known that certain underlying conditions (e.g. obesity, diabetes, heart disease) are important risk factors for developing severe COVID-19, hospitalization and death. As the population makeup of individuals with underlying conditions can be very different in different countries, it is then key to determine region-based estimates of COVID-19 related hospitalizations and deaths, so that models can be adequately parameterized. Further, we compared modeled peak hospitalizations to current state goals for hospital bed occupancy, but it is possible that a lower vaccine effectiveness or a lower vaccination coverage could achieve the same goals because deterministic models tend to overestimate the transmission dynamics. We computed the optimal allocation strategies utilizing age as the sole risk factor. However, other factors, like occupation have been linked to an increased risk of acquisition and severe disease (***?***, *32*). Furthermore, several studies (*33, 34*) have shown that, as a result of health systems with systemic health and social inequalities, people from racial and ethnic minority groups are at increased risk of getting sick and dying from COVID-19 in certain countries. These are crucial considerations that will be included in further studies and can point towards who, within a given age group, should get the vaccine first.

We believe that these results can provide a quantification of the effectiveness of different allocation scenarios under four metrics of disease burden and can be used as an evidence-based guidance to vaccine prioritization.

## Data Availability

Data is available in the manuscript.

https://github.com/lulelita/vaccine_optimization

## Contributors

LM and ERB conceived the study. LM developed the model. LM and JE wrote the optimization algorithms. LM and TL analyzed the data. LM produced the first draft of the manuscript. All authors contributed to the final draft.

## Acknowledgments

LM acknowledges Mia Moore for helpful discussions regarding the model structure and Benjamin E. McGough for help with cluster computing.

## Data and materials availability

All data needed to evaluate the conclusions in the paper are present in the paper and/or the Supplementary Materials. Code available at https://github.com/lulelita/vaccine_optimization.

Additional data related to this paper may be requested from the authors.

## Appendix

### Mathematical model

We developed a deterministic mathematical model with 16 age groups: 0–4, 5–9, 10–14, 15– 19, 20–24, 25–29, 30–34, 35–39, 40–44, 45–49, 50–54, 55–59, 60–64, 65–69, 70–74, and ≥75. For each age-group, the population is divided into the following compartments: those who are susceptible (*S*) to infection; exposed (*E*) but are not yet infectious; infectious; and recovered (*R*). Infectious individuals are classified by severity of symptoms. Those who are infectious may be asymptomatic (*A*) or pre-symptomatic (*P*). As pre-symptomatic individuals become symptomatic, they may not require hospitalization (*I*), require hospitalization (*H*), or require intensive care (*C*). Because of the short duration of our simulation, we did not model any births or deaths. We assumed a population of 7.615 million people, the current population of Washington State (*35*) and parameterized the demographic composition of the population to be similar to that of the United States (*36*).

The model equations (where a dot represents differentiation with respect to time) are:

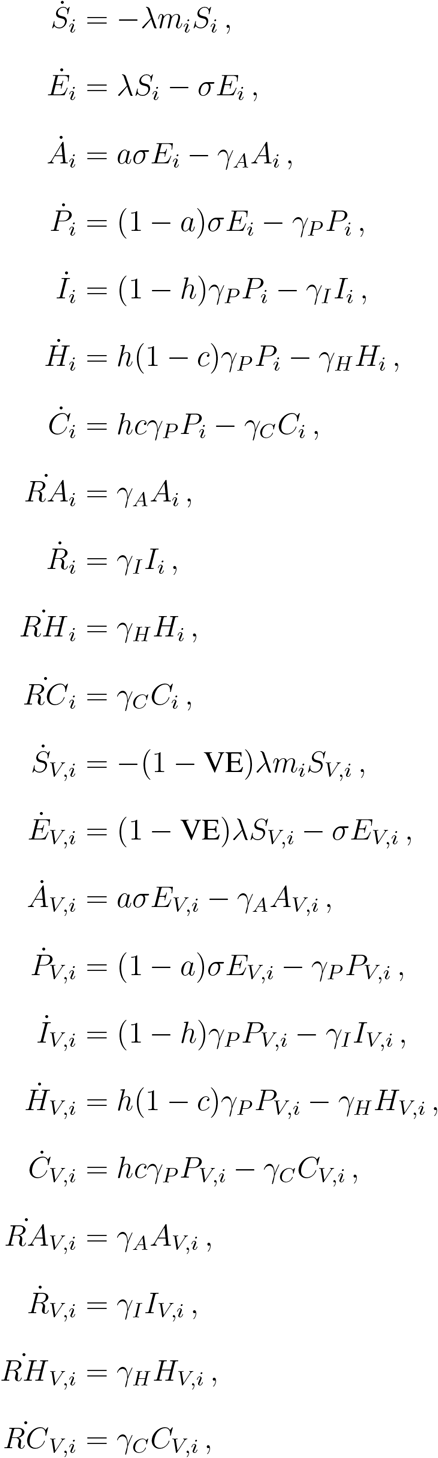

where the force of infection is

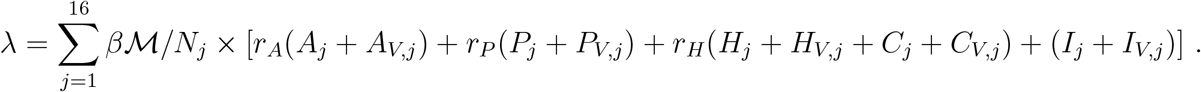

A diagram of the model is shown in Fig.S1.

We used the age-specific contact matrix ℳ for the US given in (*37*) and corrected for reciprocity. We assumed *R*_0_ = 3. We used previously reported age-specific estimates of the severity of infections that require hospitalization, critical care and that lead to death given by Ferguson et al. (*12*). This report has age-specific percentages of symptomatic cases requiring hospitalizations, percentages of hospitalized cases requiring critical care and the infection fatality ratio for the population divided in 10 age groups: 0-9, 10-19, …, 70-80 and ≥ 80. Because we have 16 age groups, we considered the rates in our groups to be similar within a given age group given in (*12*). For example, the rates of hospitalization and mortality for the groups 0-5 and 5-10 in our model are those in the age group 0-9 in (*12*). For the oldest group in our model (≥ 75) we did a weighted average of the rates in (*12*) according to the relative percentages of the population aged 75-80 and ≥ 80. With these rates in hand, we used the total number of infections and symptomatic infections given in our model to compute the number of hospitalizations (and then the number of ICU hospitalizations) and deaths. An average hospital stay lasts 5 days without intensive care, or 10 days if intensive care is required (*11*). We assumed a latent period of three days and an infectious period for non-hospitalized infections of 3 and 5 days respectively (*17, 38*). We assumed that immunity would last at least one year, which corresponds to the duration of our simulations.

A proportion, (1 − *a*), of infections is symptomatic. Of these, a proportion, *h*, requires hospitalization. Further, a proportion, *c*, of hospitalizations requires intensive care (ICU). Symptomatic and asymptomatic individuals are assumed equally infectious. Pre-symptomatic individuals, however, are assumed to be more infectious to match the finding that 40% of transmission occurs prior to symptom onset (*11, 39*). Once hospitalized, individuals are assumed no longer infectious. We determined the optimal allocation strategies assuming both that susceptibility to infection is age-specific and that susceptibility to infection is equal across all age groups. See Table S1 for details.

Simulations were run with initial conditions set to a 20% recovered population distributed proportionally to population size (pro-rata) and disease severity, respectively. In addition, simulations were started assuming 1,000 current infections distributed among the infectious symptomatic and asymptomatic infectious compartments (compartments *A*_*i*_, *A*_*V,i*_, *I*_*i*_, *I*_*V,i*_, *H*_*i*_, *H*_*V,i*_, *C*_*i*_ and *C*_*V,i*_).

#### Vaccination

We assumed a leaky vaccine (*40*) that reduces susceptibility to infection, modeled as a reduced probability of acquiring a SARS-CoV-2 infection. For each vaccination coverage and vaccination strategy considered, we computed within each age-group the fraction of susceptible people among all those individuals in that group who could have sought the vaccine (susceptible, exposed, infected pre-symptomatic, infected asymptomatic, and recovered asymptomatic populations), and utilized that fraction as the fraction of people who were actually vaccinated in each age-group, while assuming that the remaining vaccine would be wasted.

Further, we did not model the vaccination campaign. Our simulations start assuming that the population to be vaccinated has already been vaccinated, and that vaccinated individuals have reached the full protection conferred by the vaccine.

#### Vaccine effectiveness against COVID-19 disease VE_*COV*_

We also explored the optimal allocation strategies with a vaccine that would reduce susceptibility to infection and that would also reduce probability of having a COVID-19 disease (denoted by VE_*COV*_) as follows. We define the vaccine effect for susceptibility VE as a measure of the vaccine to reduce the probability of a person getting infected. We define the vaccine effect for COVID-19 disease and infection as VE_*P*_ as a measure of the vaccine to reduce symptoms given infection and VE_*COV*_ as the vaccine effect for reducing COVID-19 disease (*41*). Then it follows that

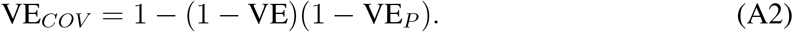

In our analysis, we assumed that VE_*COV*_ = 60, in line with the expected effectiveness in the current vaccine trial protocols (***?***, *10*), Equation (A2) implies that for this value of VE_*COV*_, VE and VE_*P*_ need to be between 0 and 60%. Hence, we restricted VE to this range for this part of the analysis. The equations of vaccinated compartments in the model were then modified as follows:

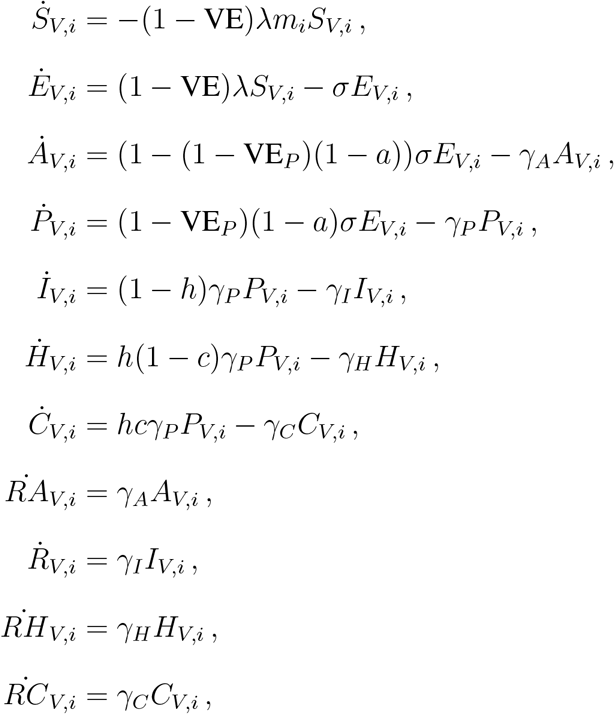

#### Uncertainty analysis

We performed uncertainty analysis in two ways. First, we examined the robustness of the optimal vaccination strategies against the number of initial infections, the fraction of the population with natural immunity at the start of the simulations and the relative susceptibility of children and older adults to infection. In addition, we selected four parameters (the basic reproduction number *R*_0_, the fraction of asymptomatic infections, and the reduction/increase in infectiousness for asymptomatic and pre-symptomatic infections) that we considered would have the most influence in the optimal allocation strategies and re-run the optimization for the 100 VE-vaccination coverage pairs with 36 different combinations of those parameters. Table S2 provides the combinations of values used for this part of the analysis.

Second, we examined the uncertainty in the output measures (number of infections, number of deaths, etc) arising from uncertainty surrounding the model parameters, and chose those parameters for which there is the least agreement. These parameters were: *R*_0_, the mean pre-symptomatic period, the mean infectious period of non-hospitalized symptomatic infections, the proportion of infections that are asymptomatic, and the relative infectiousness of asymptomatic infections. We sampled 1000 parameter sets from pre-determined distributions as follows. For *R*_0_, the proportion of asymptomatic infections, and the multiplier increasing or decreasing the infectiousness of pre-symptomatic infected individuals, we used truncated normals. For the duration of the latent and infectious periods (both pre- and post symptoms) we utilized gamma distributions. Finally, for the relative infectiousness of asymptomatic infections, we chose among three values: 0.2, 0.5 and 1. Table S1 gives the ranges utilized for each of these parameters. We sampled 1000 parameter sets and ran the model with those sets for the optimal allocation strategy for each combination of VE and vaccination coverage pair. Further, we removed the top and bottom 2.5% of the simulations.

#### Optimization

##### Objective functions

We performed the optimization routine to minimize four different objective functions: total symptomatic infections, total deaths, maximum number of hospitalizations not requiring intensive care and maximum number of hospitalizations requiring intensive care. For each of these, we ran the deterministic model for one year. Further, we evaluated the optimal vaccine allocation for 100 VE-vaccination coverage combinations (VE ranging from 10 to 100% and vaccination coverage ranging from 10 to 100% of the total population, in increments of 10%).

##### Optimization routine

For the optimization routine, we collated the population into 5 vaccination groups: 0–19 years old, 20–49 years old, 50–64 years old, 65–74 years old and those aged 75 and older. These vaccination groups were chosen for two reasons; clinically, these groups represent the finest granularity that we could find in the data and computationally, this reduces the dimension of the optimization problem. We defined a decision variable in terms of the proportion total vaccine available as follows: let *v* be a decision vector *v* = (*v*_1_, *v*_2_, …, *v*_5_), where *v*_*i*_ represents the *proportion of the total available vaccine to be given to vaccination group i*. For each objective function, our optimization routine consisted of two steps:

1. We performed an exhaustive search on a coarse grid (*42*) of the unit simplex in the vaccination group space (the set of vectors (*v*_1_, *v*_2_, …, *v*_5_) with non-negative entries such that 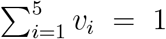). The grid was chosen so that the unit simplex was divided into 0.05 units and was computed in Sage (*43*). For example, a point in this grid is *v* = (0.05, 0.5, 0.1, 0.2, 0.15), that corresponds to utilize 5, 50, 10, 20 and 15% of the available vaccine in vaccination groups 1 through 5 respectively (as noted above, this vector would be repaired if vaccine exceeds the population in any vaccination group).
2. We selected the best 25 decision variables obtained above, the pro-rata allocation vector and an additional 25 decision variables sampled uniformly from the unit simplex (*44*), and used these 51 points as initial points for the Nelder-Mead minimizer implemented in SciPy (*45, 46*). To note, we also ran the optimization using particle swarm optimization algorithm (*47, 48*) and obtained similar optimal solutions, but solutions obtained with Nelder-Mead gave slightly better optimal values and ran faster.

##### Feasibility

We defined the decision variable in terms of proportion of total vaccine available because this allowed us to perform an exhaustive search on a coarse grid of the decision variable space that is independent of the population in each group. With this, our optimizer can be easily adapted to populations with different sizes and with different age group distributions. Further, this allowed us to quickly perform an exhaustive search in the decision variable space. The trade-off of this approach comes with guaranteeing feasibility.

In this setup, a feasible vector is defined as any vector *v* so that 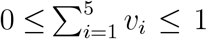, where *v*_*i*_ represents the proportion of vaccine allocated to vaccination group *i*. This guarantees that we are using at most all of the available vaccine. Because Nelder-Mead is an unconstrained optimization method, we had to “repair” any non-feasible solution *v* given by Nelder-Mead. That is, if *v* was a solution with 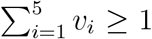, then, we radially mapped *v* to its corresponding feasible vector *v*^*^ in the unit simplex defined above.

However, this will not guarantee that the amount of vaccine given in each group does not exceed the actual population in that group. That is to say, the vaccine proportion vector may not be feasible in terms of the actual population distribution.

Hence, for any given total vaccine available, we further “repaired” any given feasible vaccine proportion vector *v*^*^ by mapping it to a vaccine group population proportion vector *x* = (*x*_1_, *x*_2_, …, *x*_5_), where *x*_*i*_ represents the fraction of the population in vaccination group *i* to be vaccinated (so that excess vaccine for that vaccination group is removed). In this way, we can guarantee feasibility in terms of both the percentage of vaccine available and people in each vaccination group.

To run the deterministic model described above, the decision variable is then transformed into a vector of size 16, where each entry corresponds to one of the 16 age groups of our model. Vaccine in each age-group was distributed proportionally to the fraction of the population that it represents within its vaccination group. For example, in the first group (people under 20 years old), children aged 0–4, 5–9, 10–14 and 15–19 represent 24%, 24.4%, 25.7%, and 25.9% of that group. Hence, within the first vaccination group, vaccine would be allocated to each of the four age groups according to those proportions. The objective functions were then evaluated via the mathematical model.

### Supplemental Figures

**Figure S1:**
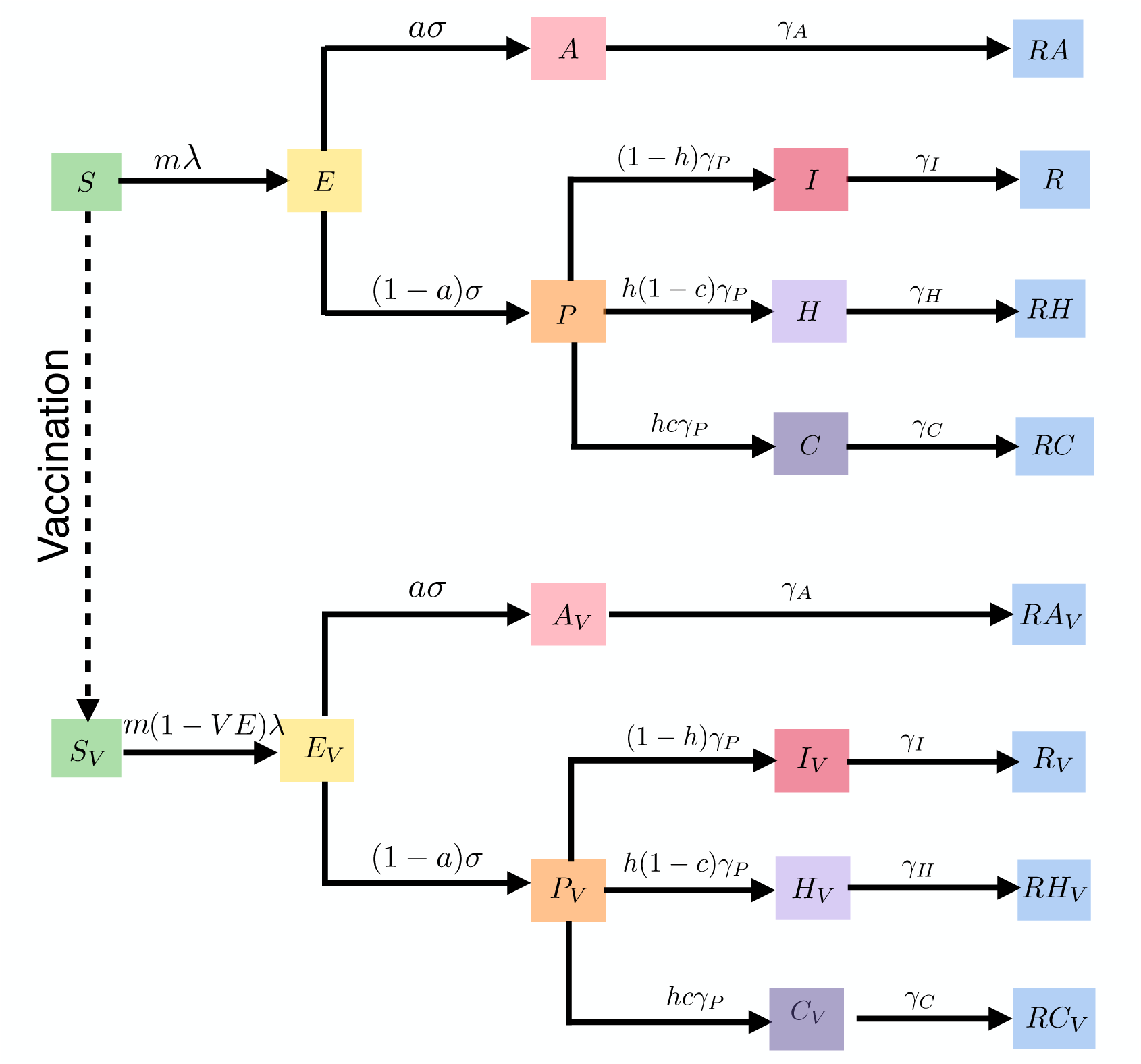
Diagram of the SEIR model without age structure.

**Figure S2:**
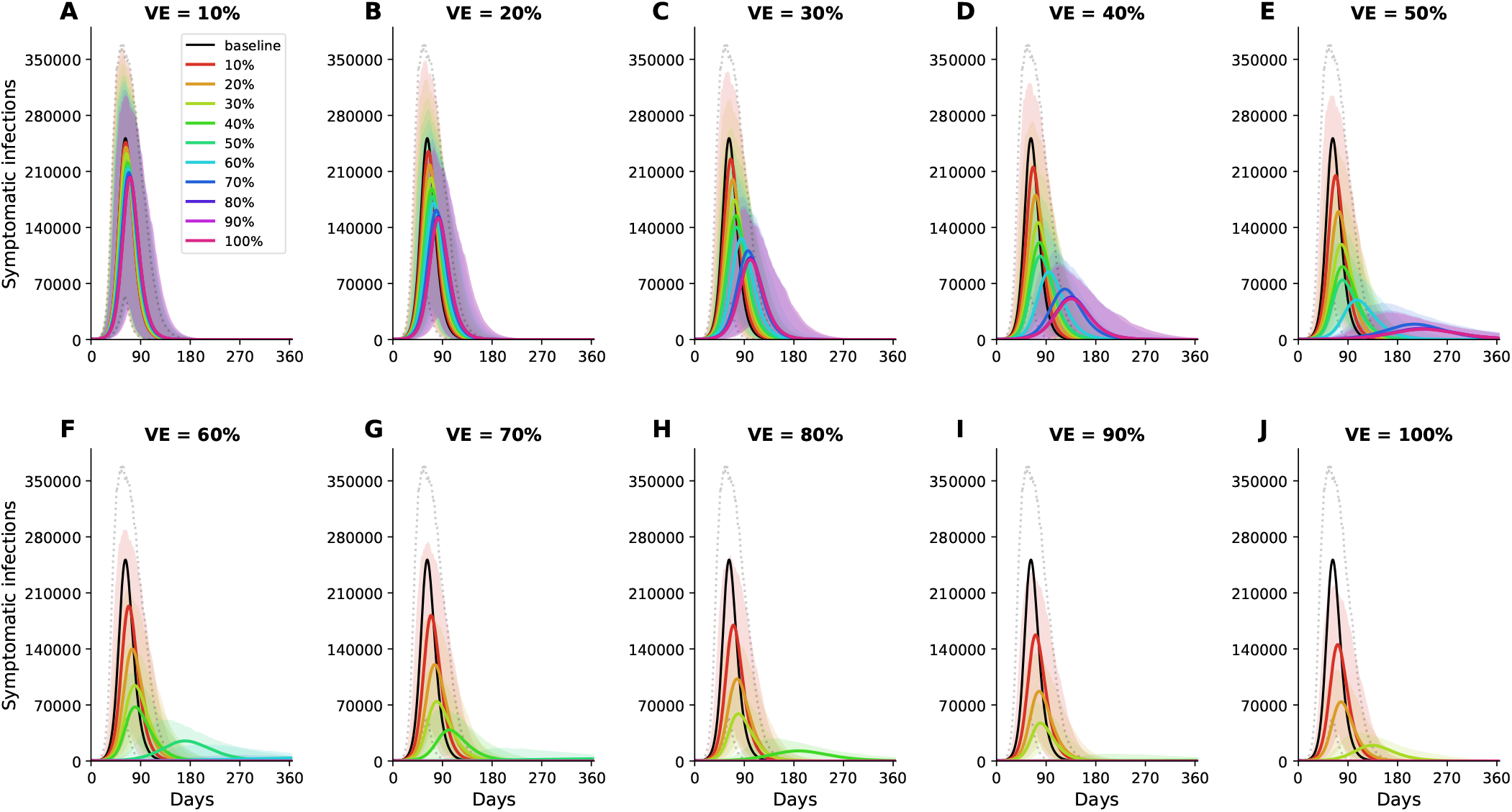
Simulated prevalence of symptomatic COVID-19 infections for VE ranging from 10% (A) to 100% (J) in 10% increments. Colors represent different vaccination coverage, ranging from 0 (black, “baseline”) to 100% (magenta). For each VE and vaccination coverage, the optimal allocation strategy to minimize symptomatic infections was utilized in these simulations. Furthermore, for each vaccination coverage, the shaded area (area encompassed by the dotted lines for baseline) represents results of 1,000 simulations with the top and bottom 2.5% simulations removed.

**Figure S3:**
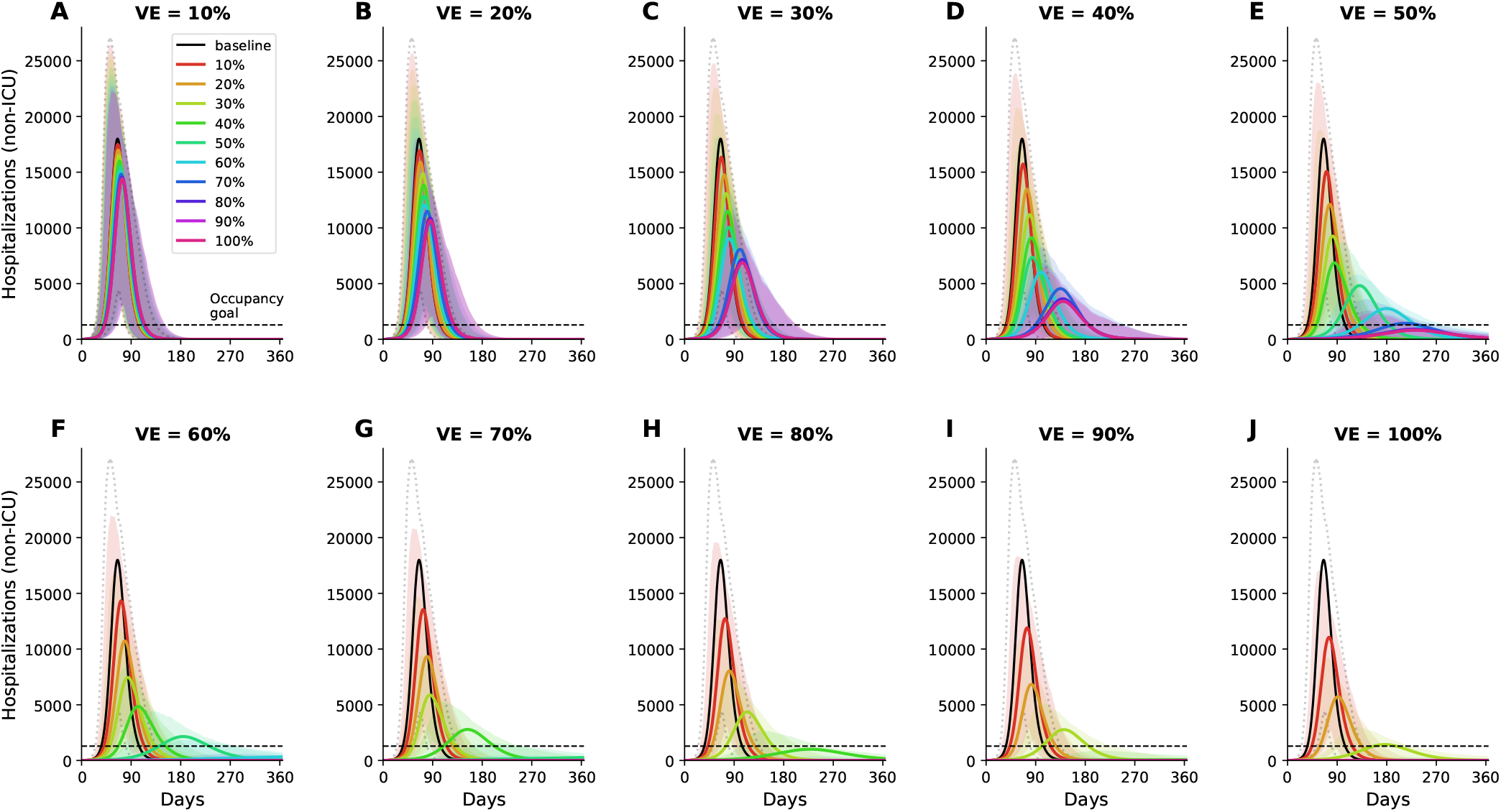
Simulated prevalence of non-ICU hospitalizations for VE ranging from 10% (A) to 100% (J) in increments of 10%. Colors represent different vaccination coverage, ranging from 0 (black, “baseline”) to 100% (magenta). The dashed line represents the current goal of having 10% of licensed general (non-ICU) hospital beds occupied by COVID-19 patients in WA state. For each VE and vaccination coverage, the optimal allocation strategy to minimize non-ICU hospitalizations was utilized in these simulations. Further, for each vaccination coverage, the shaded area (encompassed by the dotted lines for baseline) represents results of 1,000 simulations with the top and bottom 2.5% simulations removed.

**Figure S4:**
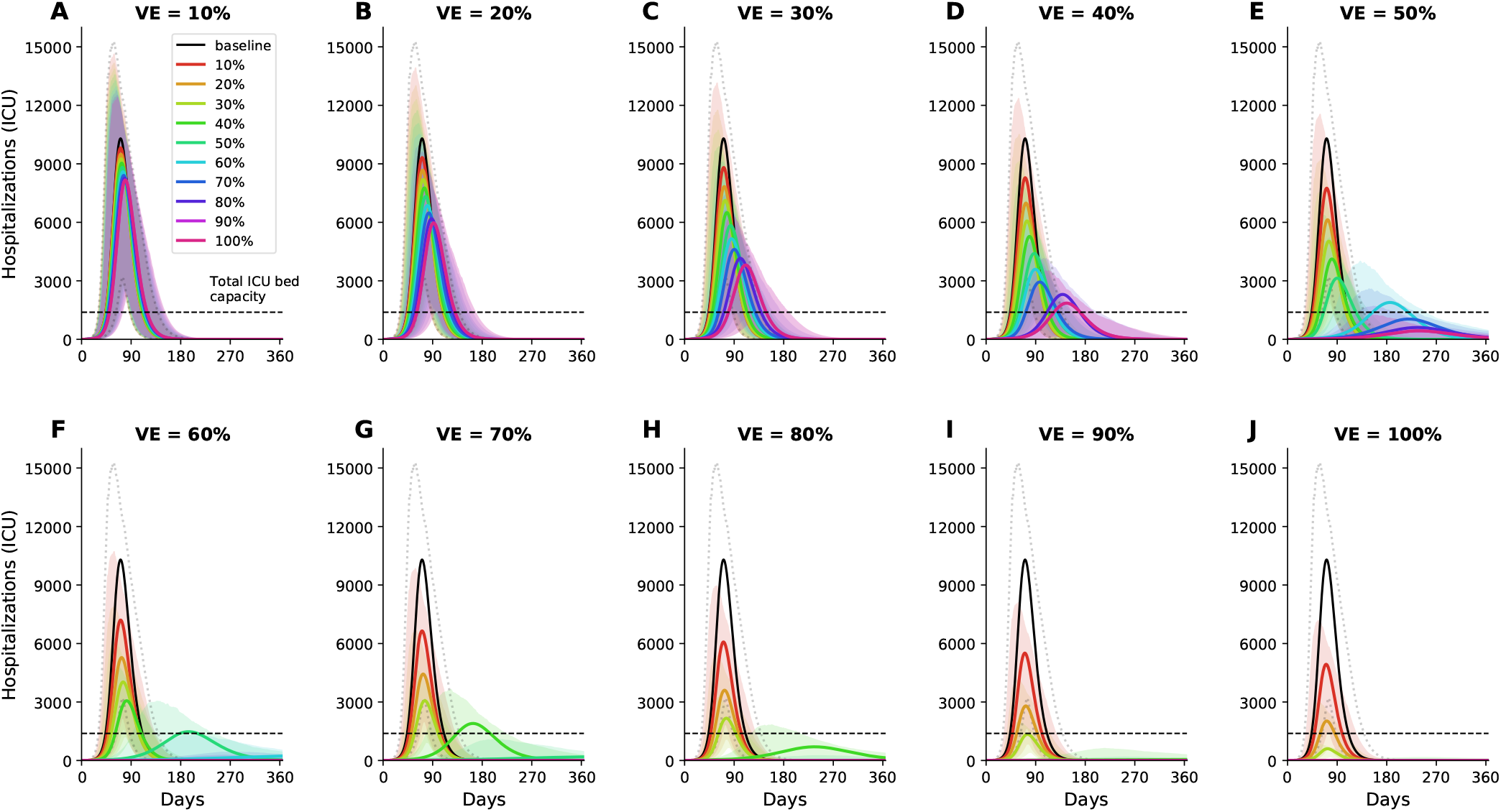
Simulated prevalence of ICU hospitalizations for VE ranging from 10% (A) to 100% (J) in increments of 10%. Colors represent different vaccination coverage, ranging from 0 (black, “baseline”) to 100% (magenta). The dashed line represents the total capacity of ICU beds in WA state. For each VE and vaccination coverage, the optimal allocation strategy to minimize ICU hospitalizations was utilized in these simulations. Furthermore, for each vaccination coverage, the shaded area (area encompassed by the dotted lines for baseline) represents results of 1,000 simulations with the top and bottom 2.5% simulations removed.

**Figure S5:**
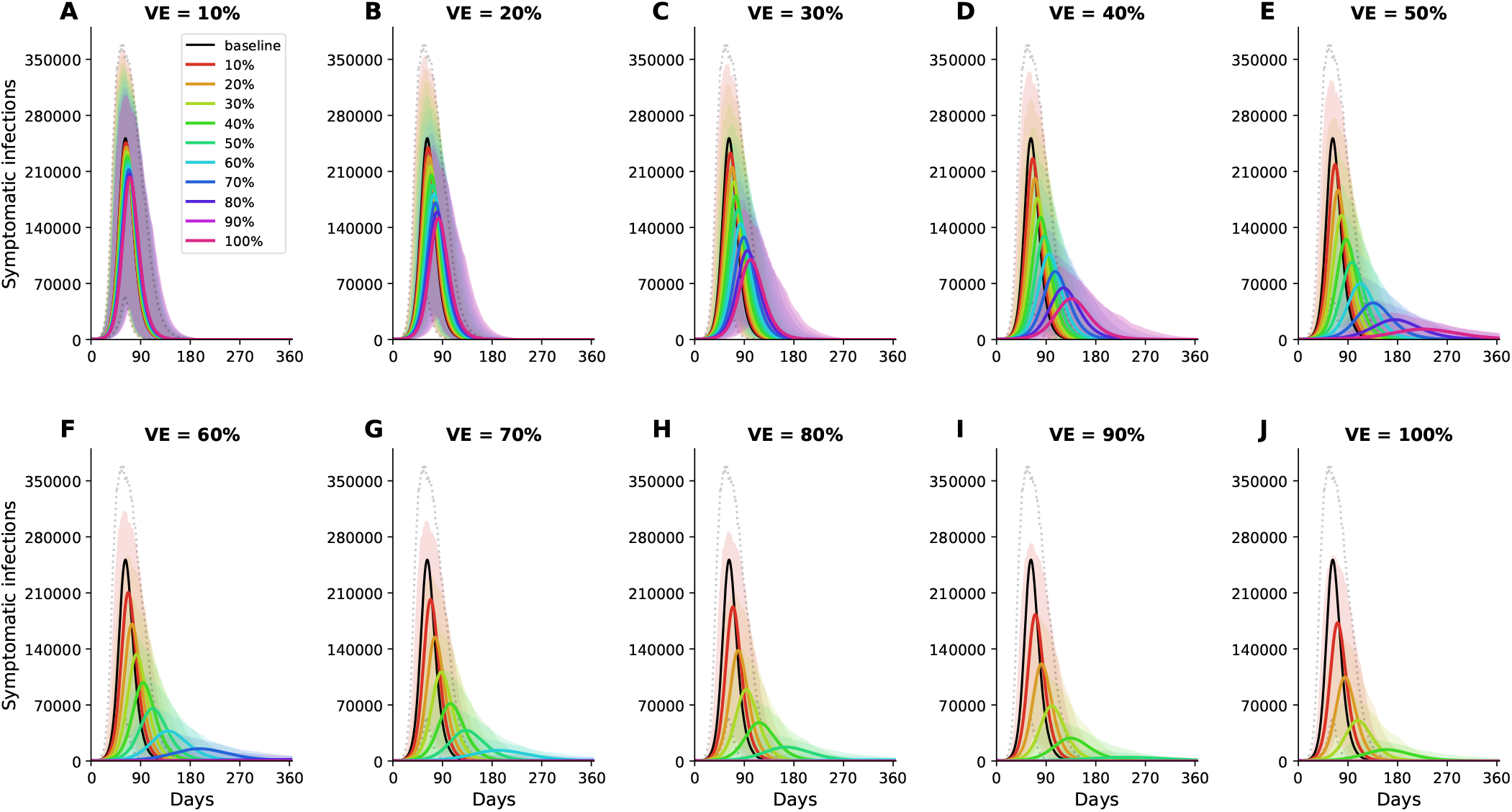
Simulated prevalence of symptomatic COVID-19 infections for VE ranging from 10% (A) to 100% (J) in increments of 10% under a pro-rata distribution of vaccine (vaccine is distributed according to the proportion of the population in each vaccination group). Colors represent different vaccination coverage, ranging from 0 (black, “baseline”) to 100% (magenta). For each vaccination coverage, the shaded area (area encompassed by the dotted lines for baseline) represents results of 1,000 simulations with the top and bottom 2.5% simulations removed.

**Figure S6:**
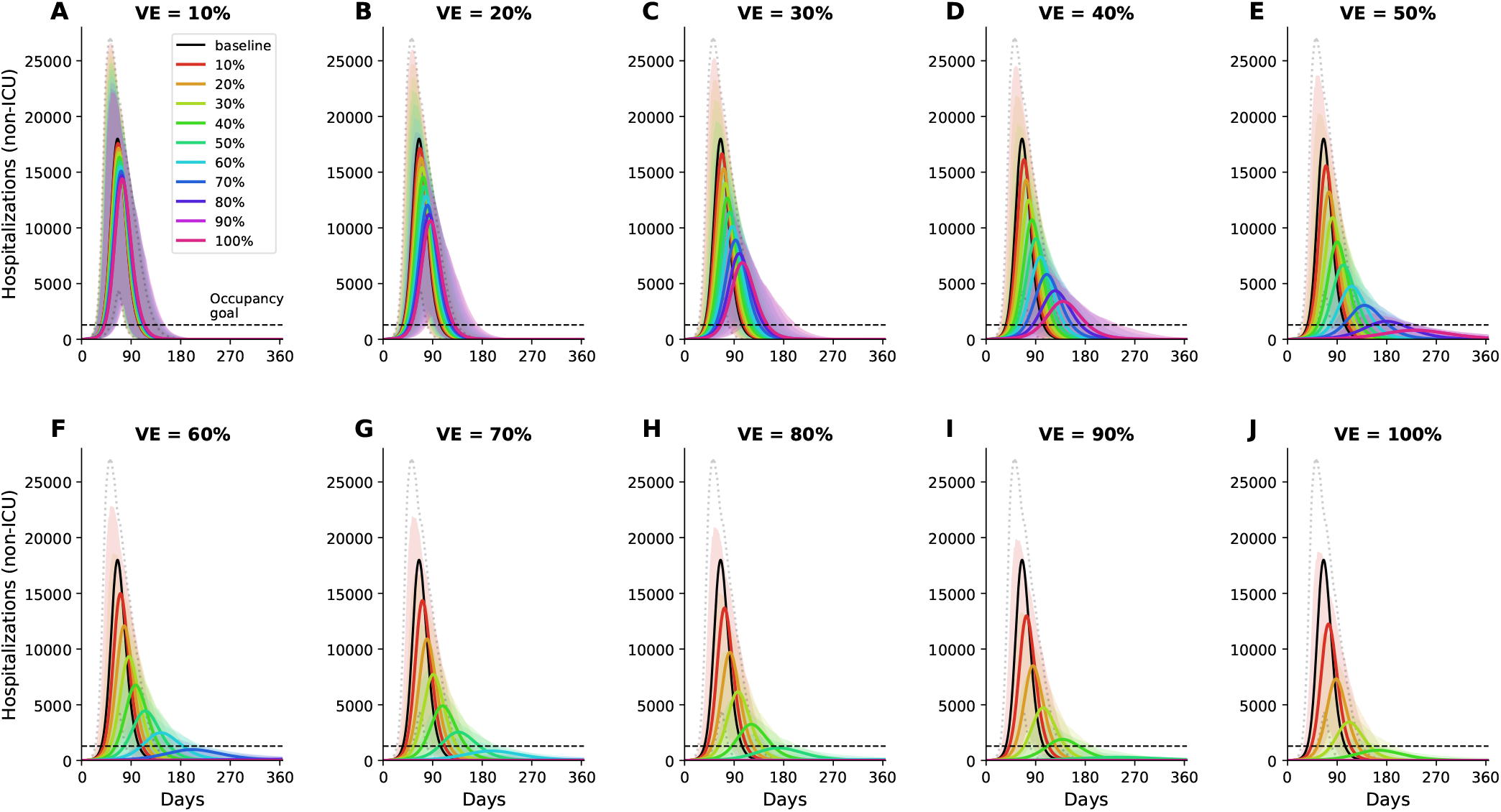
Simulated prevalence of non-ICU hospitalizations for VE ranging from 10% (A) to 100% (J) in increments of 10% under a pro-rata distribution of vaccine (vaccine is distributed according to the proportion of the population in each vaccination group. Colors represent different vaccination coverage, ranging from 0 (black, “baseline”) to 100% (magenta). The dashed line represents the current goal of having 10% of licensed general (non-ICU) hospital beds occupied by COVID-19 patients in WA state. For each vaccination coverage, the shaded area (area encompassed by the dotted lines for baseline) represents results of 1,000 simulations with the top and bottom 2.5% simulations removed.

**Figure S7:**
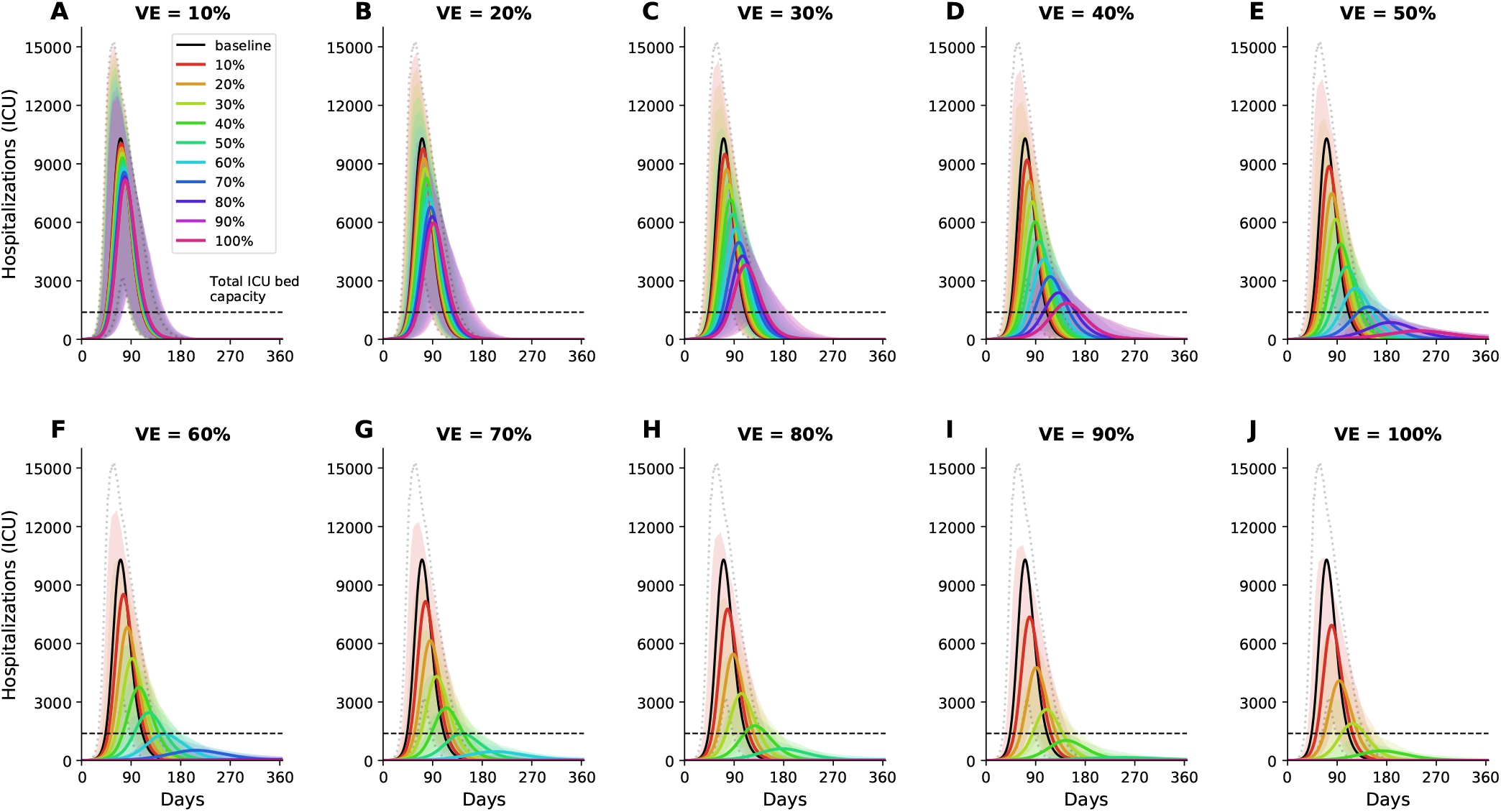
Simulated prevalence of ICU hospitalizations for VE ranging from 10% (A) to 100% (J) in increments of 10% under a pro-rata distribution of vaccine (vaccine is distributed according to the proportion of the population in each vaccination group. Colors represent different vaccination coverage, ranging from 0 (black, “baseline”) to 100% (magenta). The dashed line represents the total capacity of ICU beds in WA state. For each vaccination coverage, the shaded area (area encompassed by the dotted lines for baseline) represents results of 1,000 simulations with the top and bottom 2.5% simulations removed.

**Figure S8:**
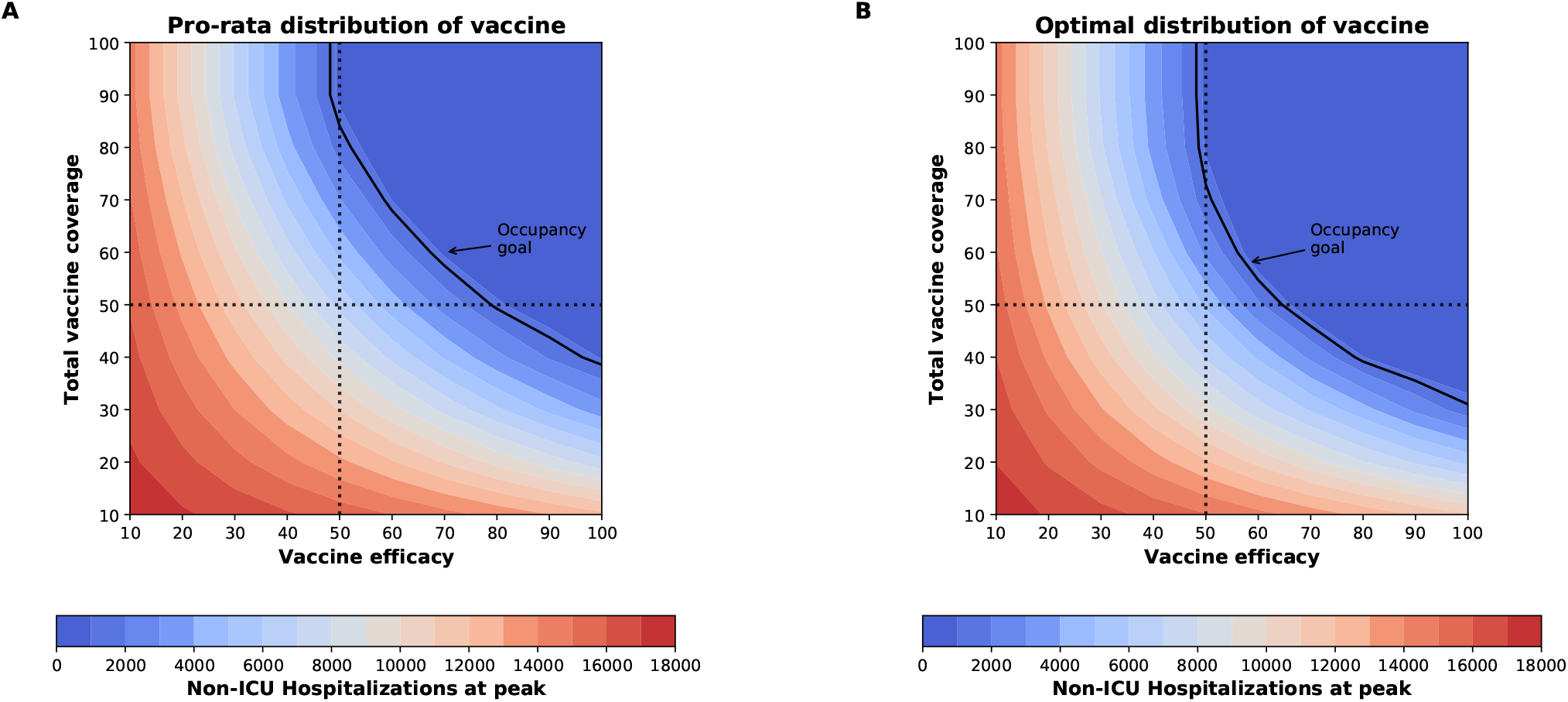
Number of maximum non-ICU hospitalizations as a function of VE and vaccination coverage (total vaccine available as a percentage of the population) for the pro-rata allocation strategy (A) and the optimal allocation strategy (B). The dotted lines correspond to VE = 50% and vaccine available to cover 50% of the population. The isoclines indicate the current goal of having 10% of licensed general (non-ICU) hospital beds occupied by COVID-19 patients in WA state.

**Figure S9:**
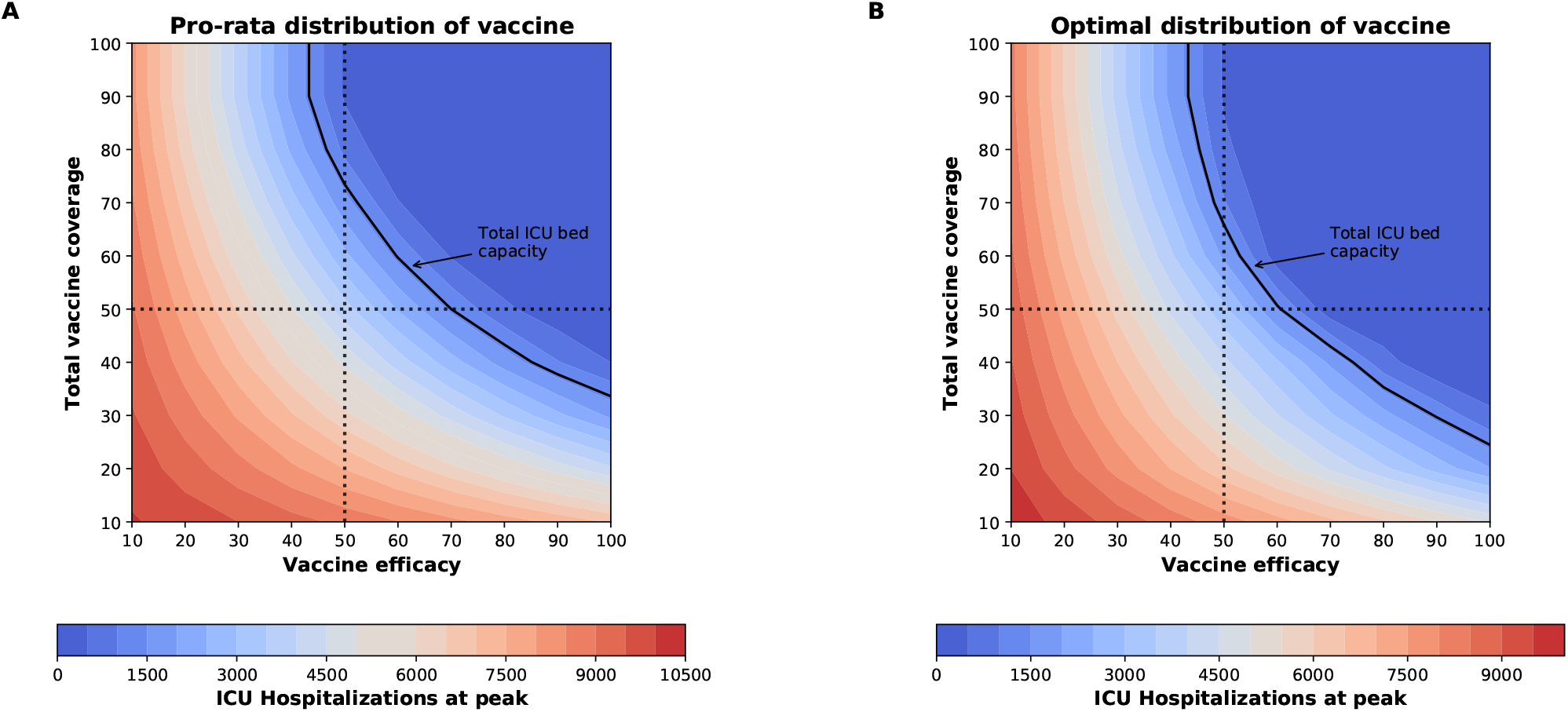
Number of maximum ICU hospitalizations as a function of VE and vaccination coverage (total vaccine available as a percentage of the population) for the pro-rata allocation strategy (A) and the optimal allocation strategy (B). The dotted lines correspond to VE = 50% and vaccine available to cover 50% of the population.The isoclines indicate the number of licensed ICU hospital beds in WA state.

**Figure S10:**
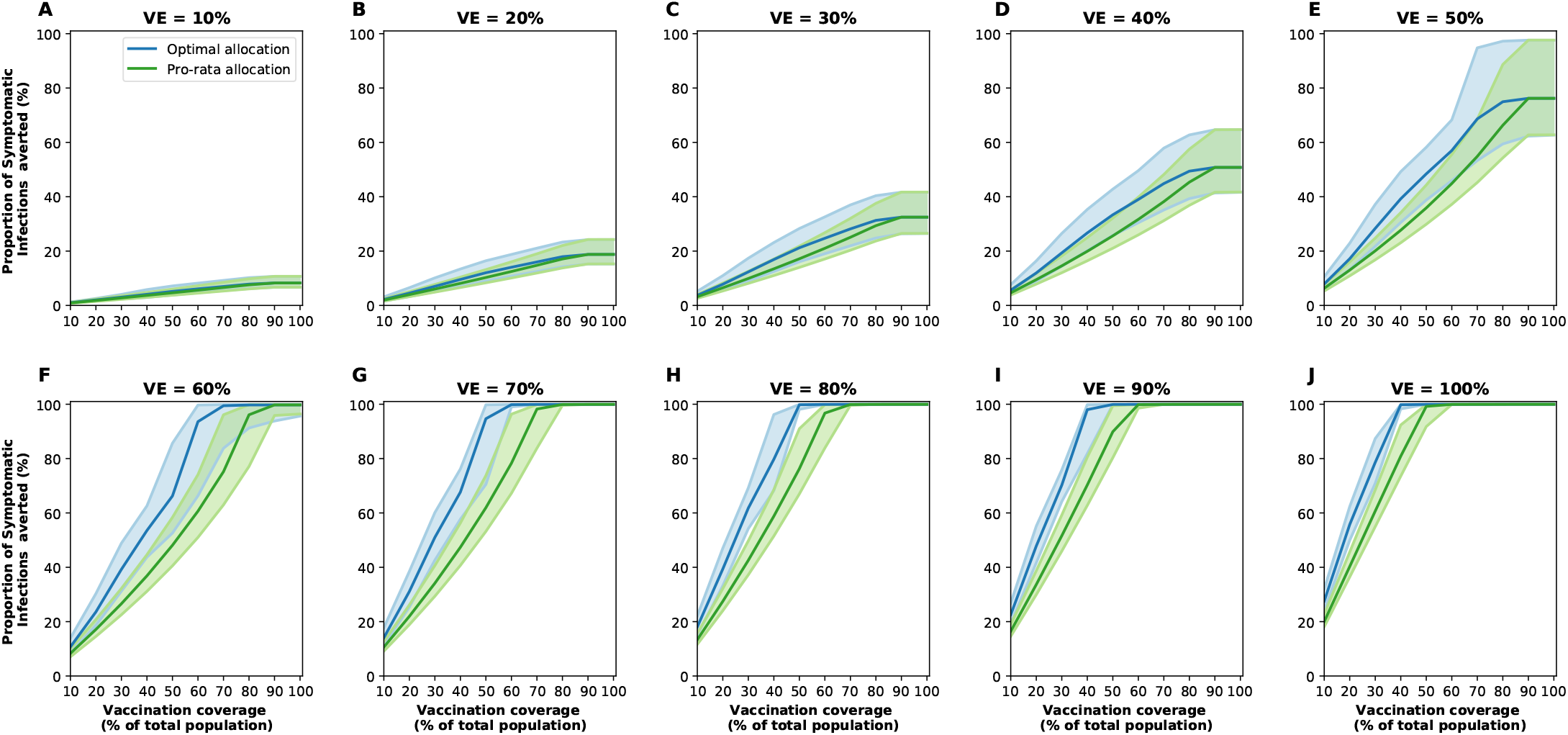
Percentage of symptomatic infections averted for the optimal allocation strategy (blue) and the pro-rata strategy (green) for VE ranging from 10% (A) to 100% (J) in 10% increments and vaccination coverage ranging from 10% to 100% of the total population. The shaded areas represent results of 1,000 parameter simulations with the top and bottom 2.5% simulations removed.

**Figure S11:**
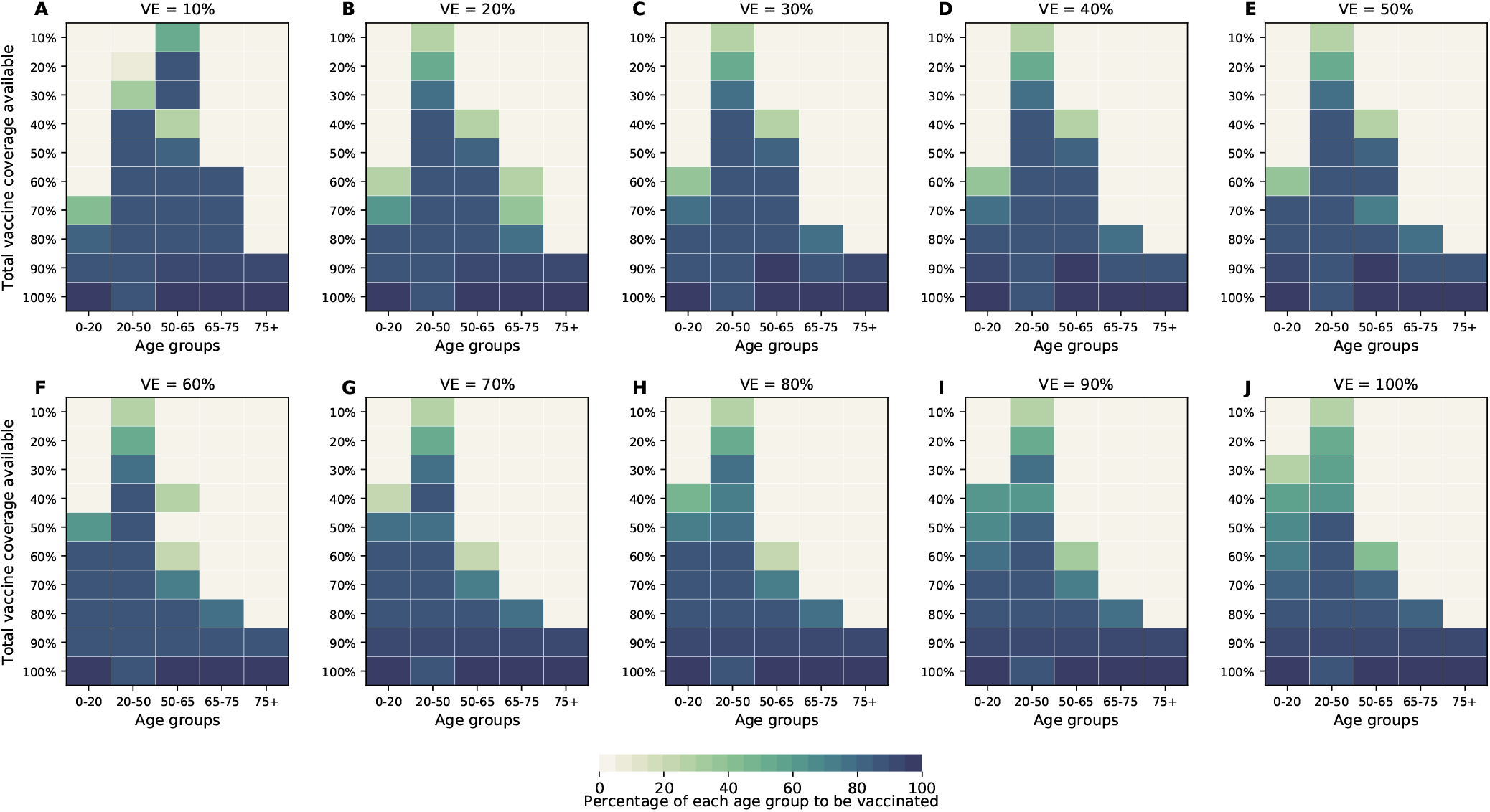
Optimal allocation strategies for minimizing symptomatic infections (with differential susceptibility) for VE ranging from 10% (A) to 100% (J) in 10% increments. For each plot, each row represents the total vaccination coverage available (percentage of the total population to be vaccinated) and each column represents a different vaccination group. Colors represent the percentage of the population in a given vaccination group to be vaccinated.

**Figure S12:**
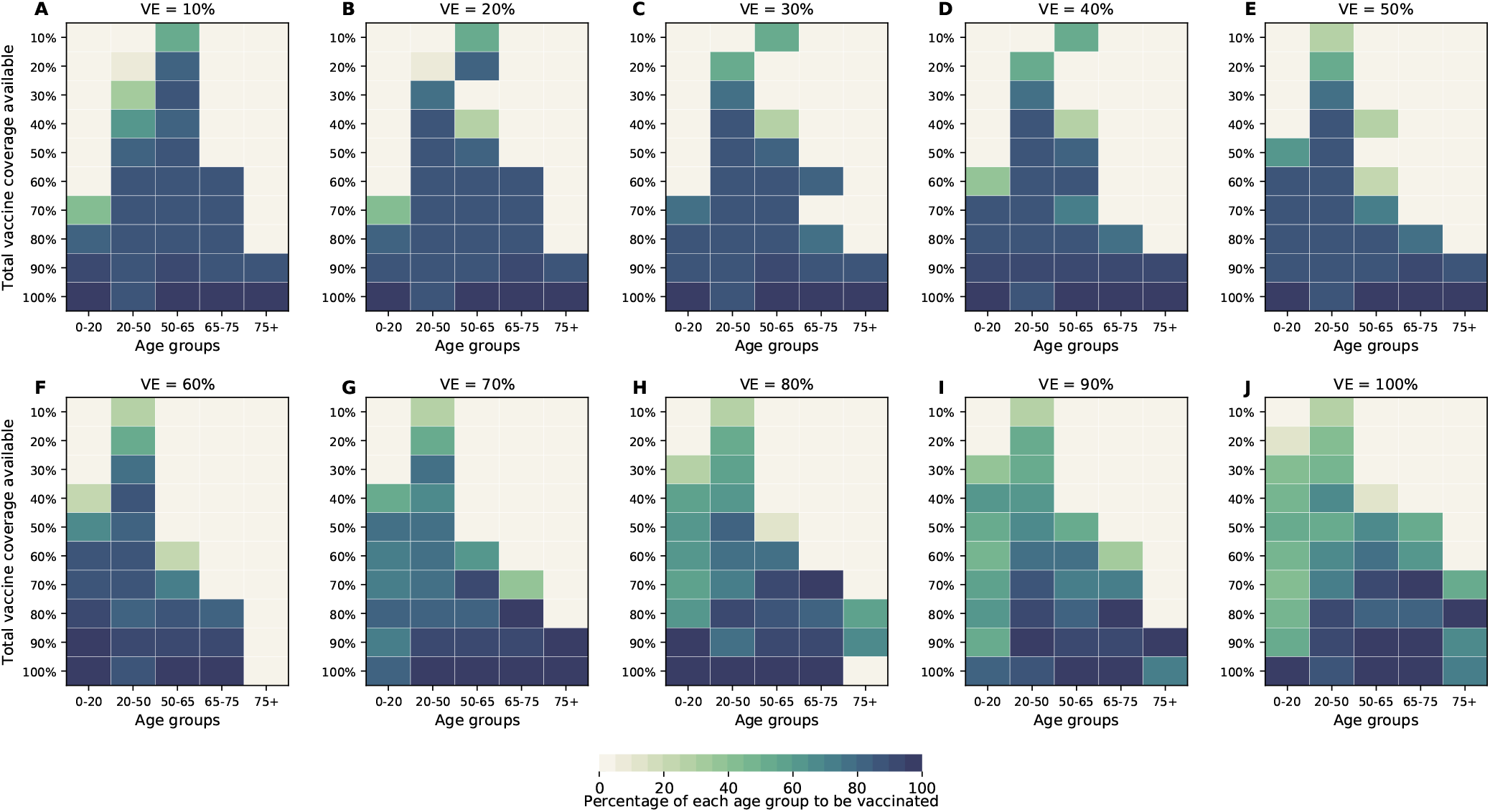
Optimal allocation strategies for minimizing the number of non-ICU hospitalizations at peak (with differential susceptibility across age groups) for VE ranging from 10% (A) to 100% (J) in 10% increments. For each plot, each row represents the total vaccination coverage available (percentage of the total population to be vaccinated) and each column represents a different vaccination group. Colors represent the percentage of the population in a given vaccination group to be vaccinated.

**Figure S13:**
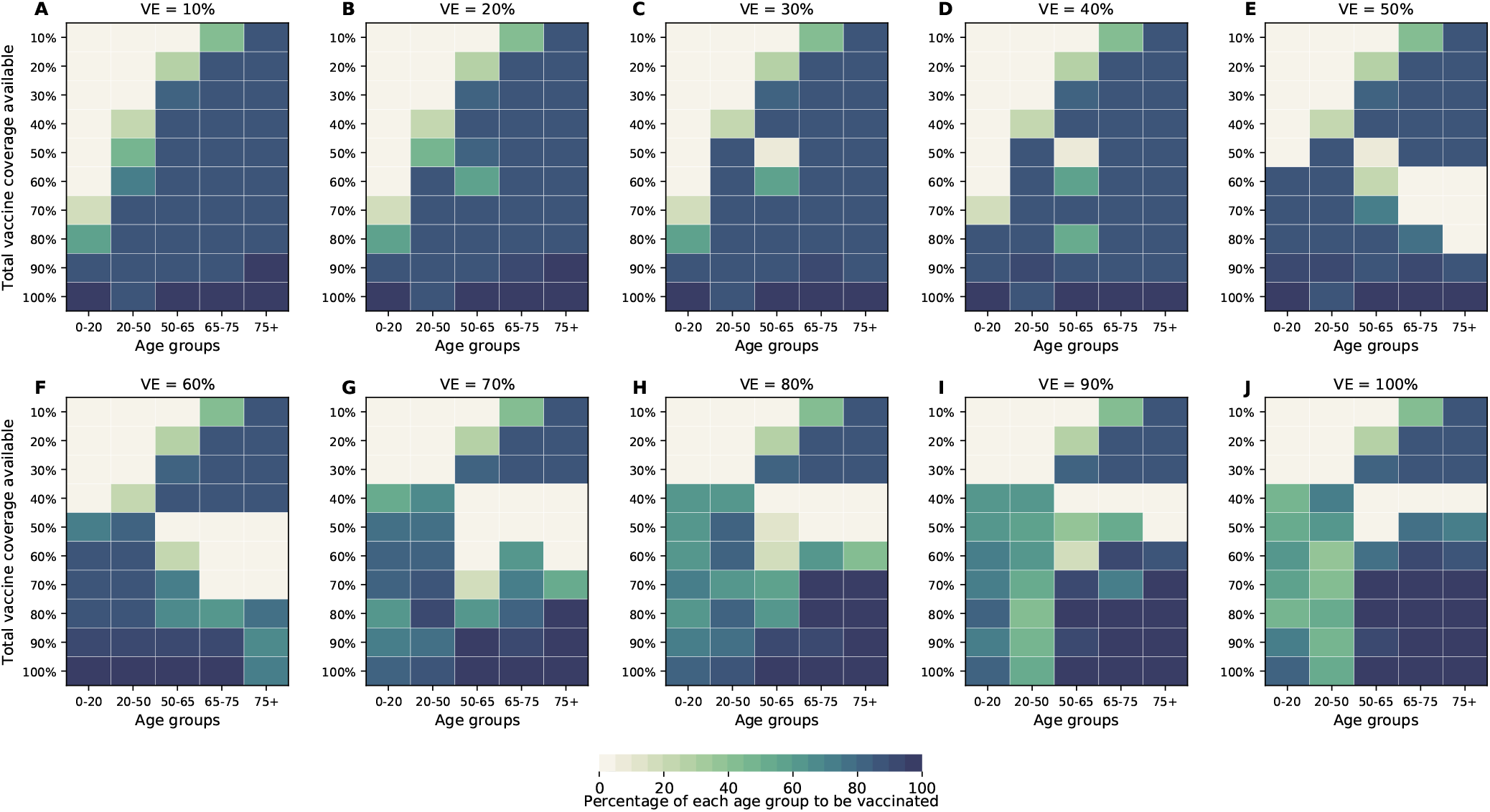
Optimal allocation strategies for minimizing the number of ICU hospitalizations at peak (with differential susceptibility) for VE ranging from 10% (A) to 100% (J) in 10% increments. For each plot, each row represents the total vaccination coverage available (percentage of the total population to be vaccinated) and each column represents a different vaccination group. Colors represent the percentage of the population in a given vaccination group to be vaccinated.

**Figure S14:**
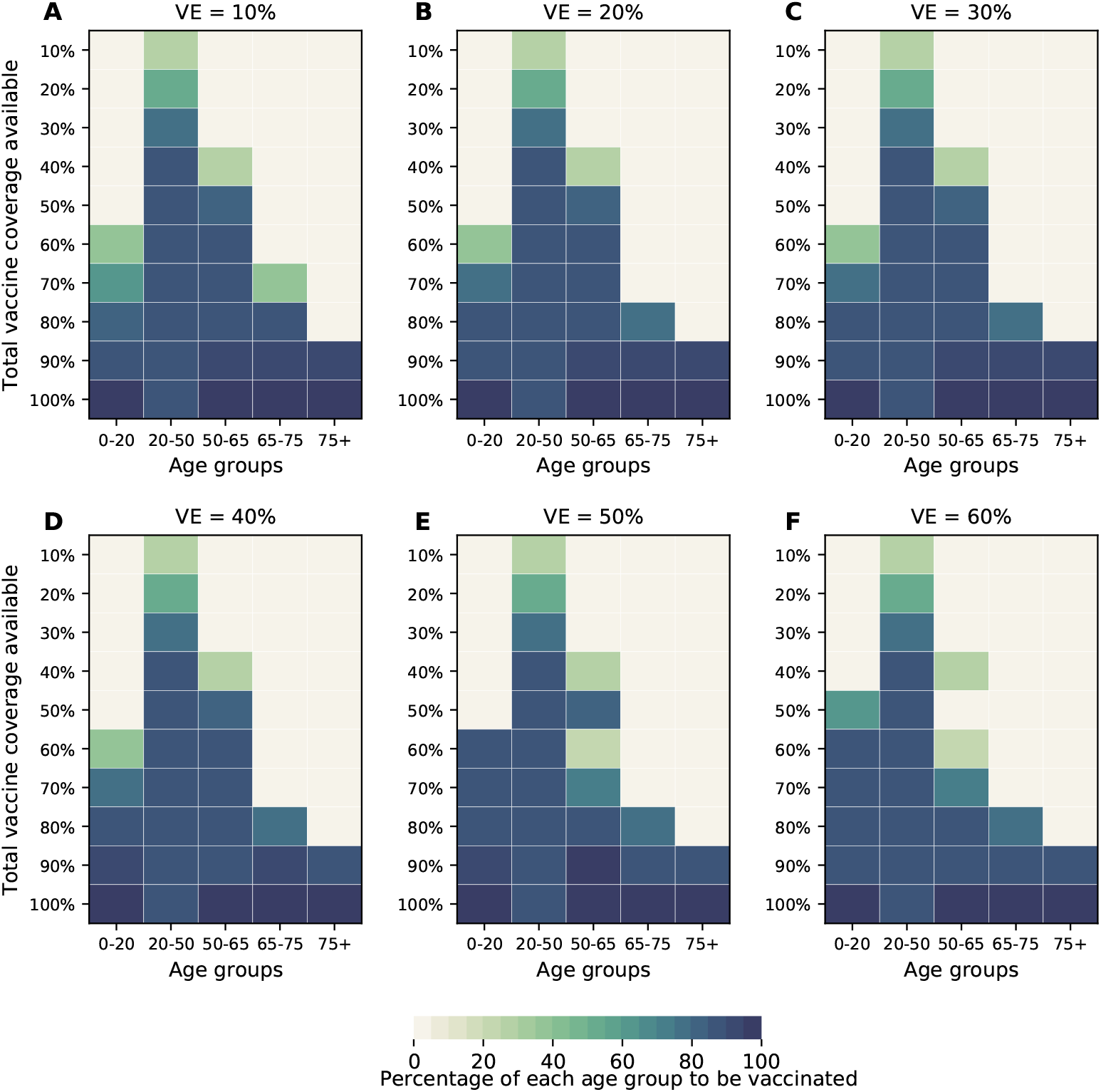
Optimal allocation strategies for minimizing symptomatic infections for VE ranging from 10% (A) to 60% (F) in 10% increments and VE_*COV*_ = 60%. For each plot, each row represents the total vaccination coverage available (percentage of the total population to be vaccinated) and each column represents a different vaccination group. Colors represent the percentage of the population in a given vaccination group to be vaccinated.

**Figure S15:**
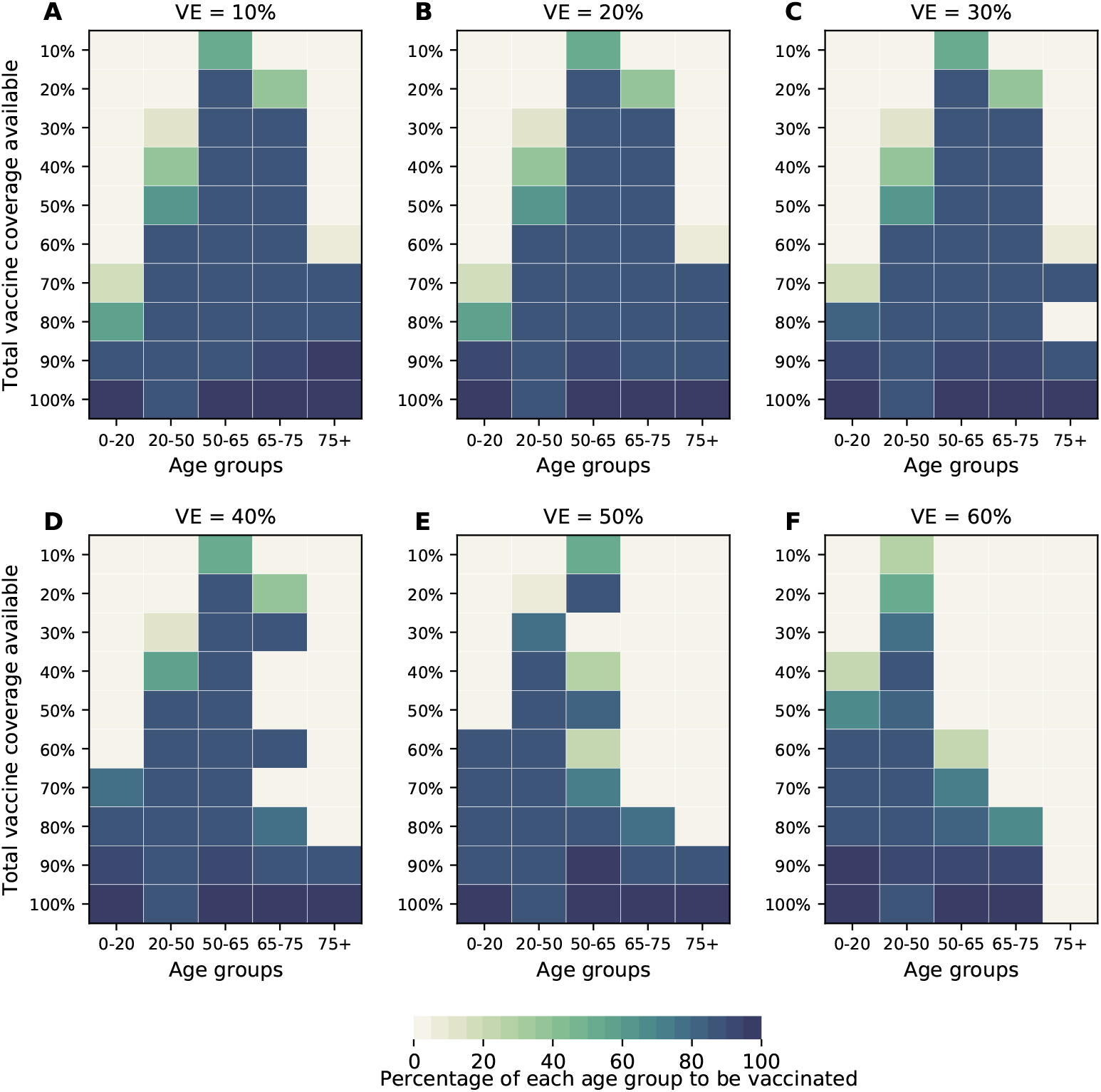
Optimal allocation strategies for minimizing the maximum non-ICU hospitalizations for VE ranging from 10% (A) to 60% (F) in 10% increments and VE_*COV*_ = 60%. For each plot, each row represents the total vaccination coverage available (percentage of the total population to be vaccinated) and each column represents a different vaccination group. Colors represent the percentage of the population in a given vaccination group to be vaccinated.

**Figure S16:**
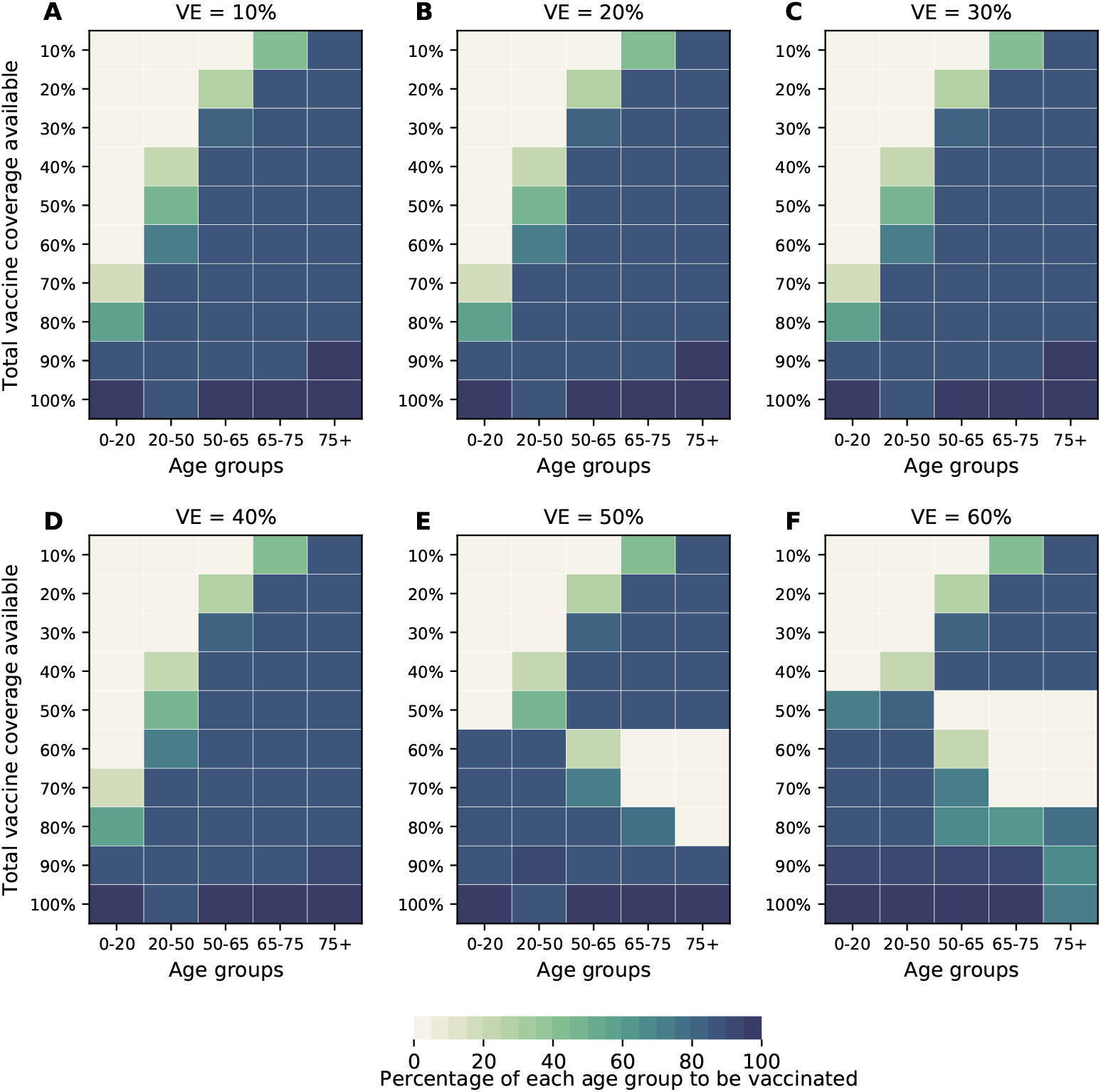
Optimal allocation strategies for minimizing the maximum ICU hospitalizations for VE ranging from 10% (A) to 60% (F) in 10% increments and VE_*COV*_ = 60%. For each plot, each row represents the total vaccination coverage available (percentage of the total population to be vaccinated) and each column represents a different vaccination group. Colors represent the percentage of the population in a given vaccination group to be vaccinated.

**Figure S17:**
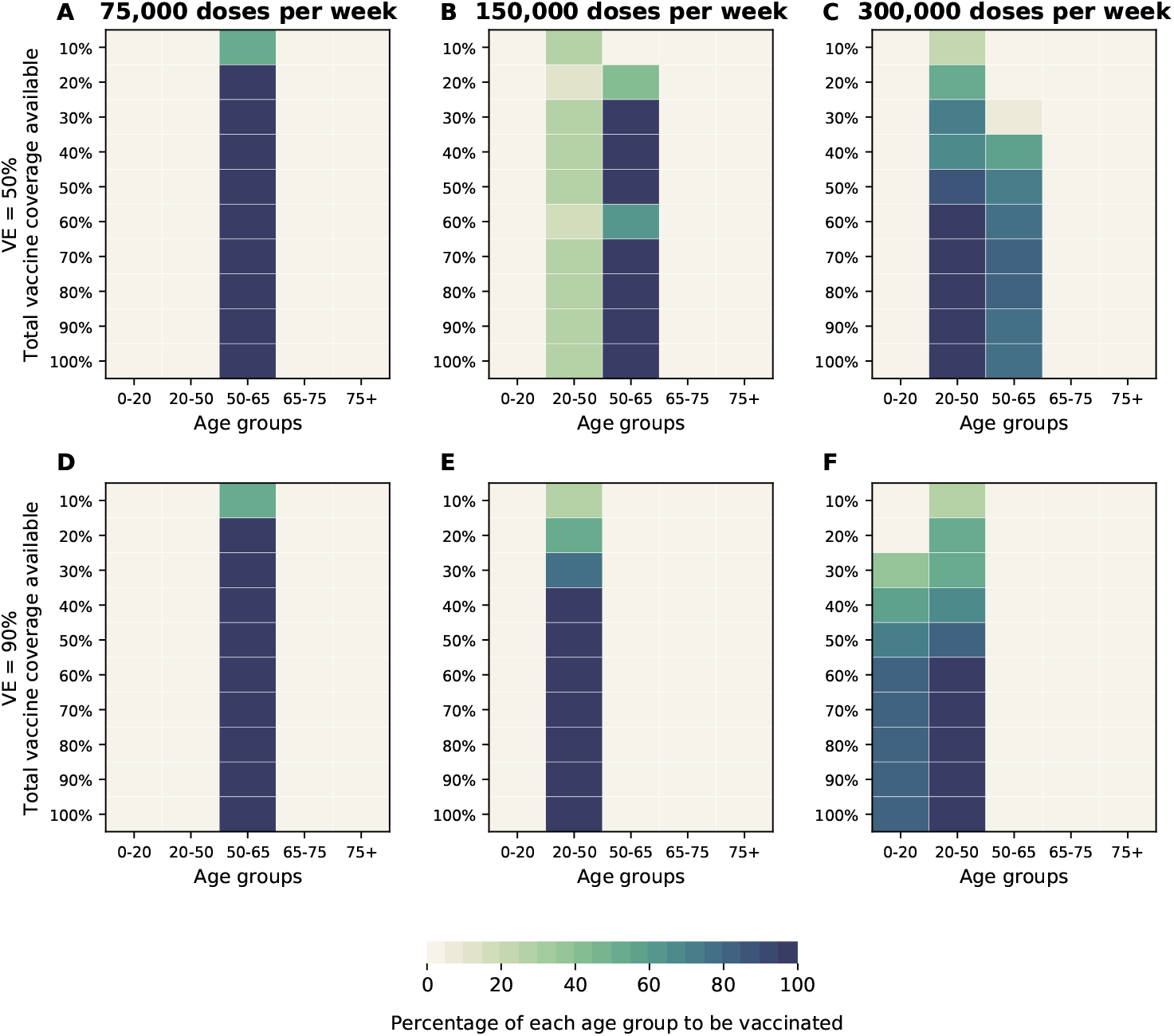
Optimal allocation strategies for minimizing peak non-ICU hospitalizations for VE = 50% (A-C) and VE= 90% (D-F) when including a vaccination campaign with 75 (A and D), 150 (B and E) or 300 (C and F) thousand vaccine doses administered per week. For each plot, each row represents the total vaccination coverage available (percentage of the total population to be vaccinated) and each column represents a different vaccination group. Colors represent the percentage of the population in a given vaccination group to be vaccinated.

**Figure S18:**
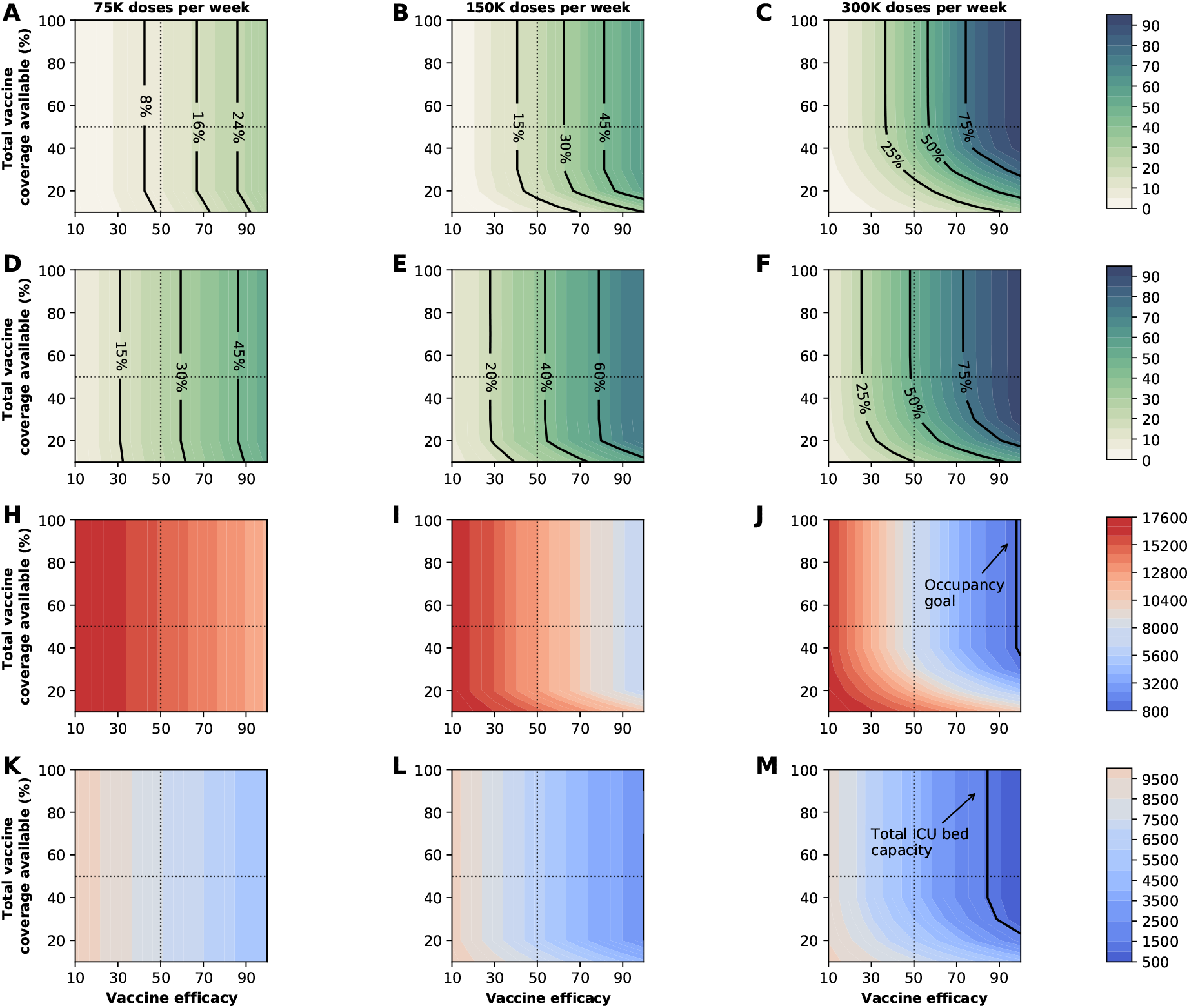
Four key metrics of COVID-19 burden under optimal distribution of vaccine. Percentage of symptomatic infections averted (A-C), deaths averted (D-F), number of maximum non-ICU (H-J) and ICU (K-M) hospitalizations as a function of VE and vaccination coverage (total vaccine available as a percentage of the population) when modeling a vaccination campaign with 75 (left), 150 (middle), or 300 (right) thousand doses of vaccine administered per week. The dotted lines correspond to VE = 50% and vaccine available to cover 50% of the population. The isoclines indicate the current goal for WA state of having 10% of licensed general (non-ICU) hospital beds occupied by COVID-19patients in (C) and total ICU licensed hospital beds in WA state in (D)

**Figure S19:**
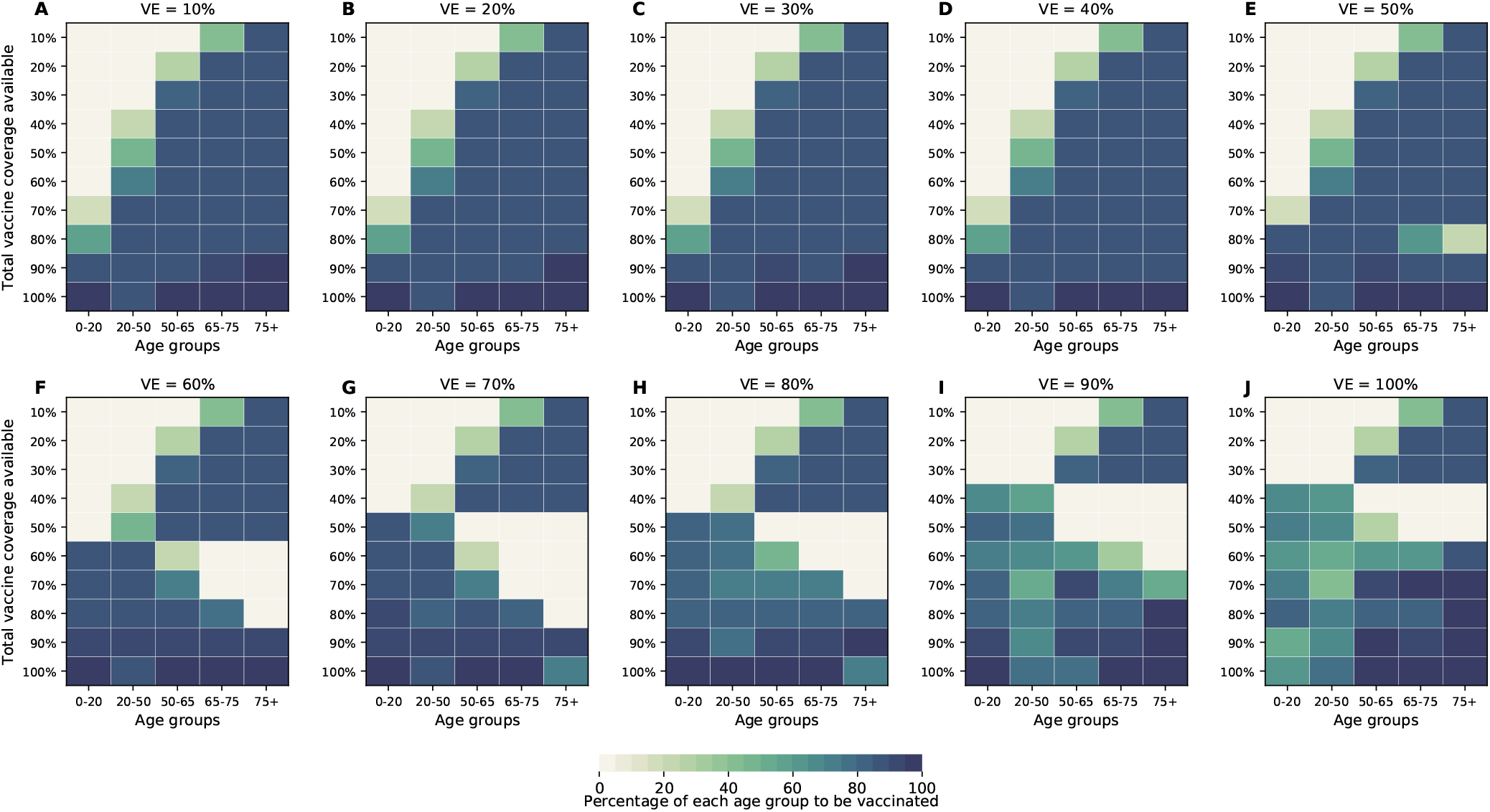
Optimal allocation strategies for minimizing deaths assuming all age groups have equal susceptibility to infection. Here, VE ranges from 10% (A) to 100% (J) in 10% increments. For each plot, each row represents the total vaccination coverage available (percentage of the total population to be vaccinated) and each column represents a different vaccination group. Colors represent the percentage of the population in a given vaccination group to be vaccinated.

**Figure S20:**
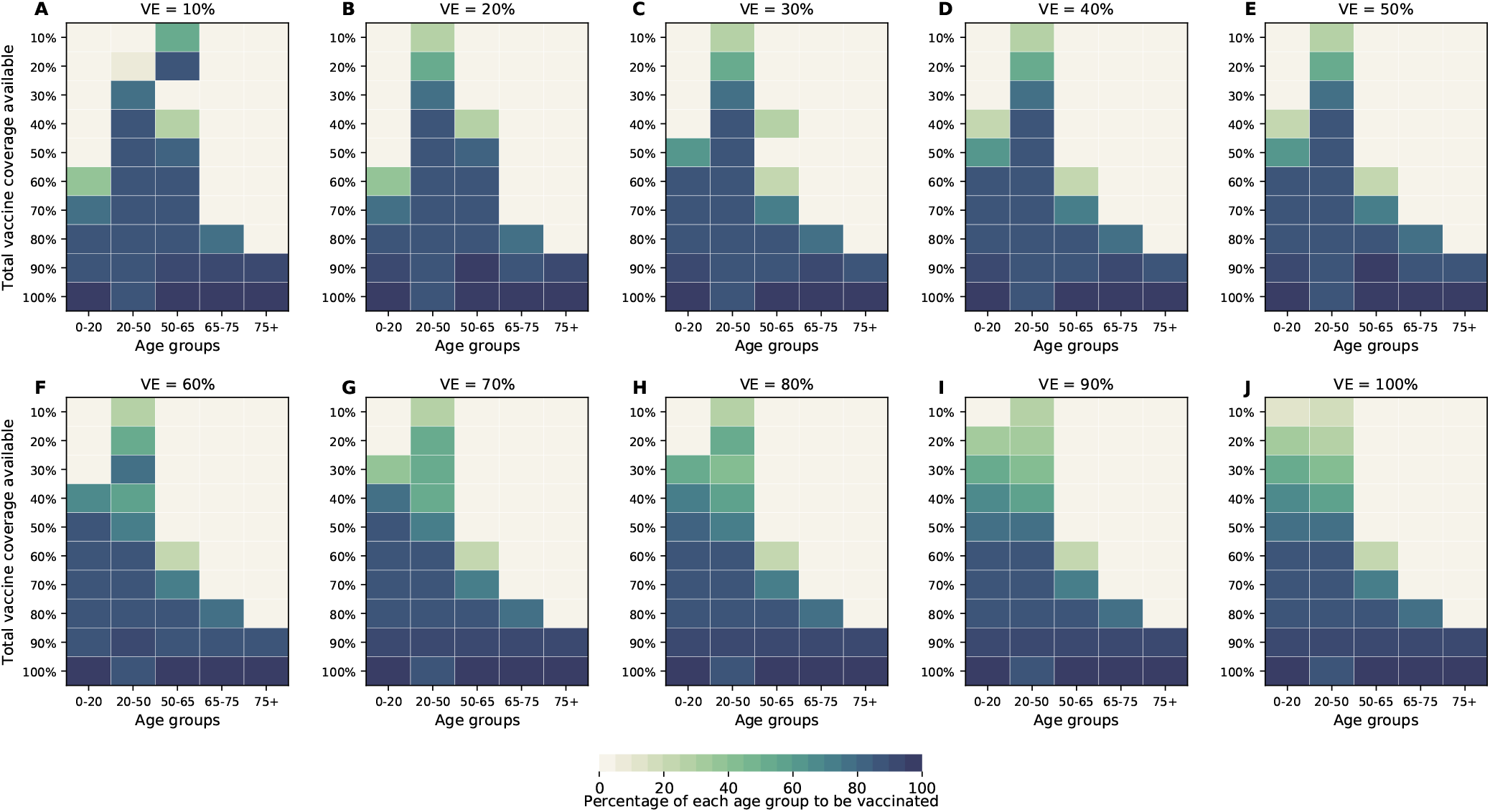
Optimal allocation strategies for minimizing symptomatic infections assuming all age groups have equal susceptibility to infection. Here, VE ranges from 10% (A) to 100% (J) in 10% increments. For each plot, each row represents the total vaccination coverage available (percentage of the total population to be vaccinated) and each column represents a different vaccination group. Colors represent the percentage of the population in a given vaccination group to be vaccinated.

**Figure S21:**
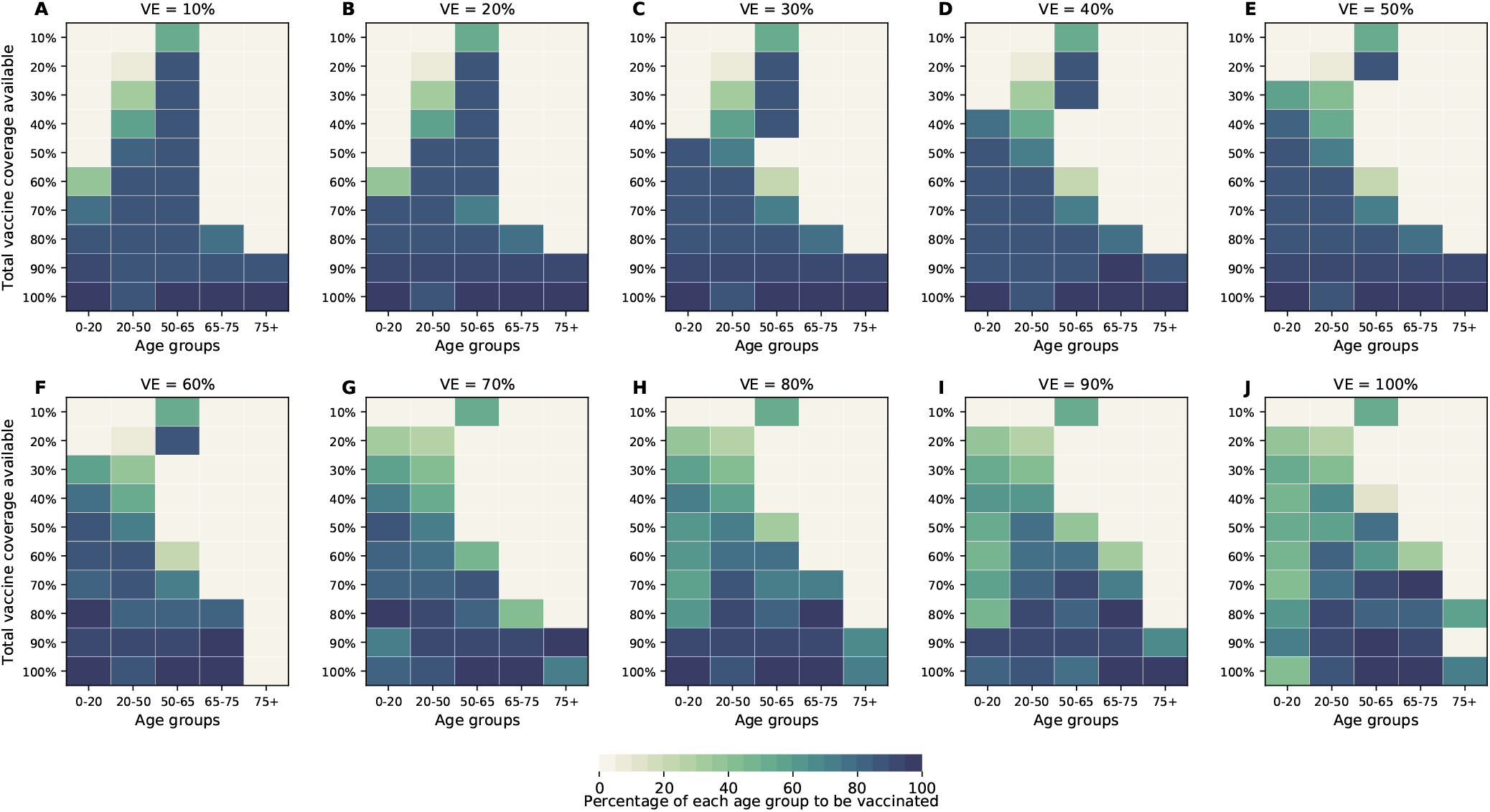
Optimal allocation strategies for minimizing the number of non-ICU hospitalizations at peak assuming all age groups have equal susceptibility to infection. Here, VE ranges from 10% (A) to 100% (J) in 10% increments. For each plot, each row represents the total vaccination coverage available (percentage of the total population to be vaccinated) and each column represents a different vaccination group. Colors represent the percentage of the population in a given vaccination group to be vaccinated.

**Figure S22:**
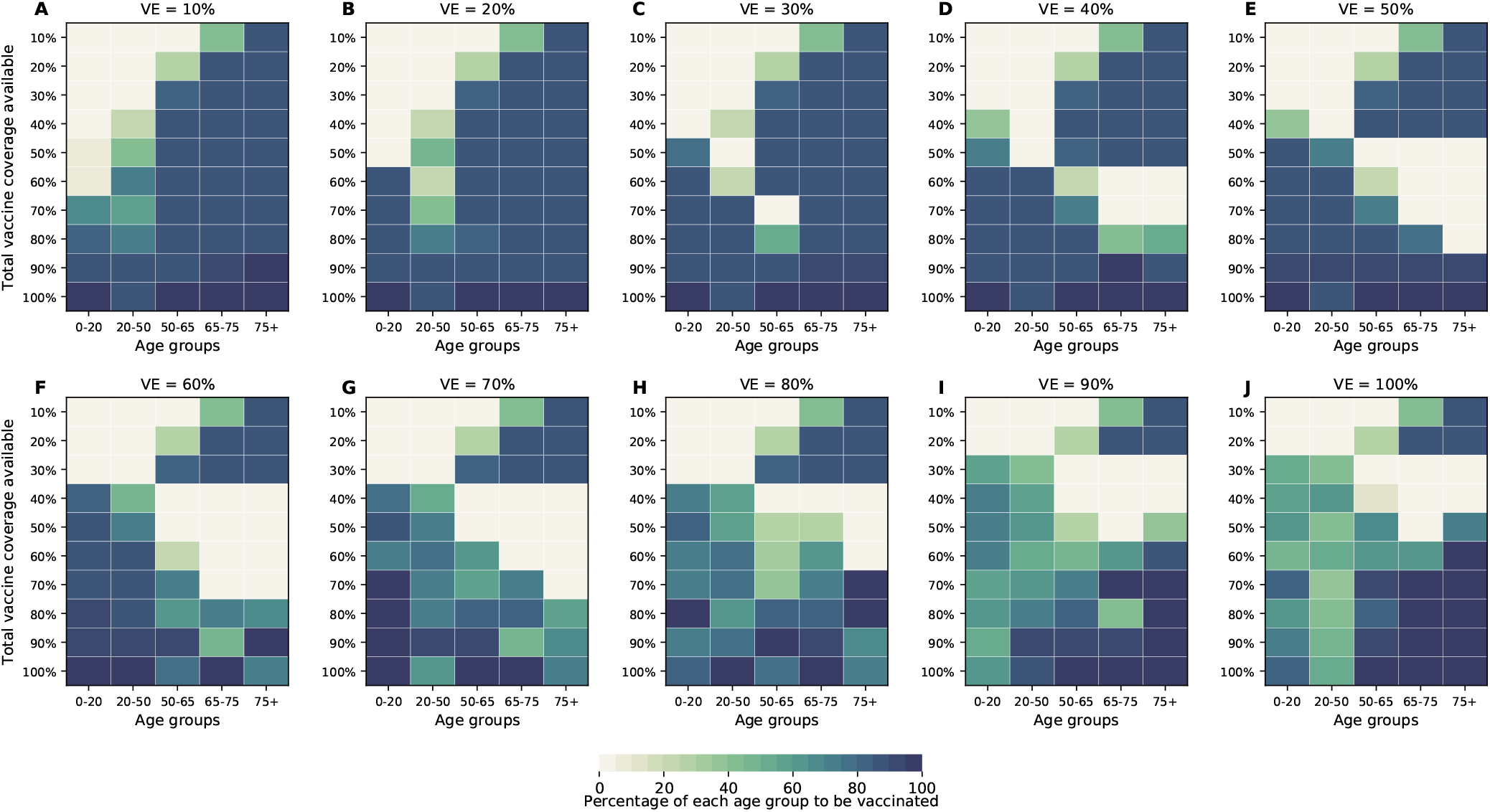
Optimal allocation strategies for minimizing the number of ICU hospitalizations at peak assuming all age groups have equal susceptibility to infection. Here, VE ranges from 10% (A) to 100% (J) in 10% increments. For each plot, each row represents the total vaccination coverage available (percentage of the total population to be vaccinated) and each column represents a different vaccination group. Colors represent the percentage of the population in a given vaccination group to be vaccinated.

**Figure S23:**
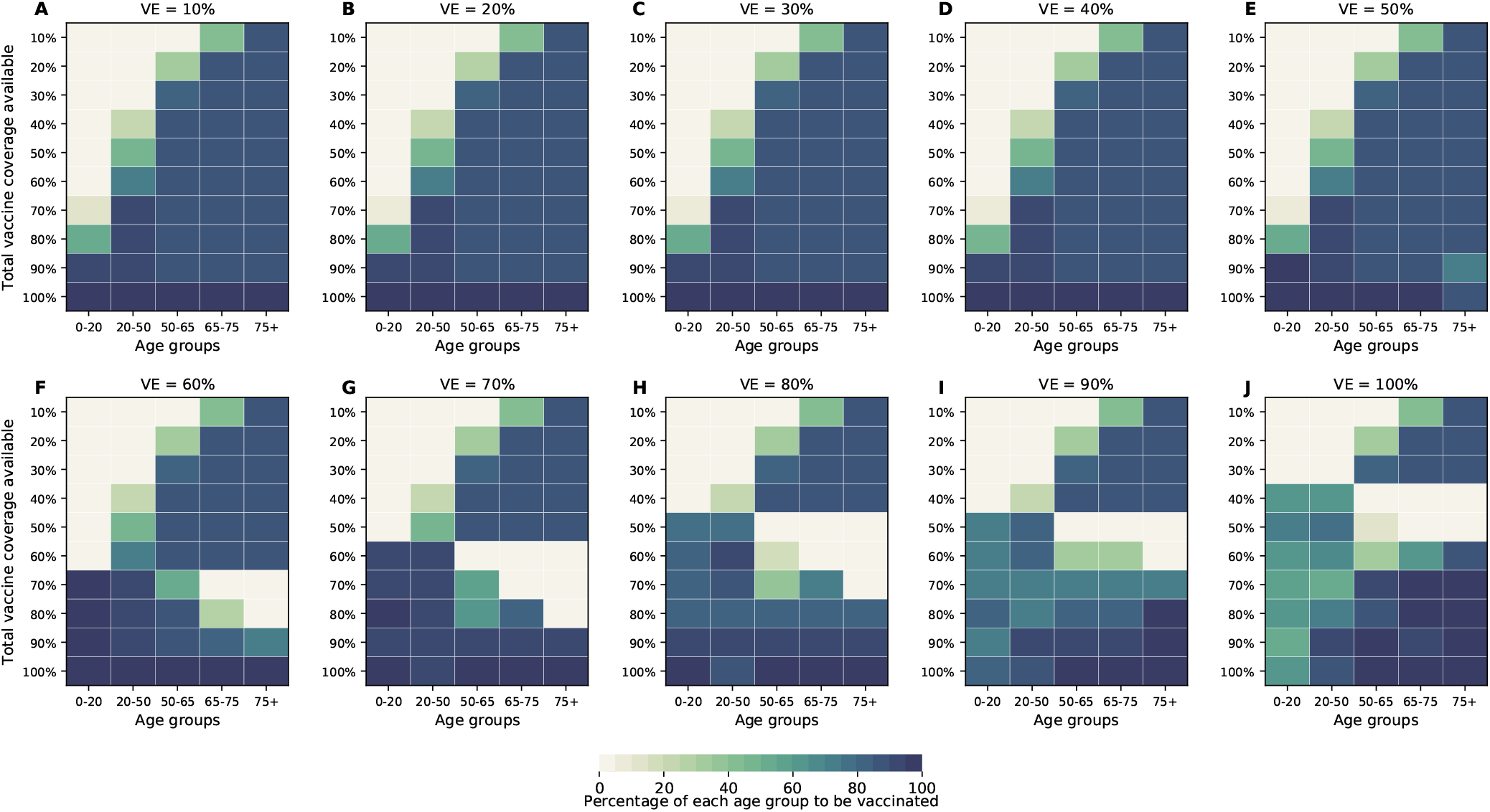
Optimal allocation strategies for minimizing the number of deaths for VE ranging from 10% (A) to 100% (J) in 10% increments assuming age-specific rates for symptomatic infection. For each plot, each row represents the total vaccination coverage available (percentage of the total population to be vaccinated) and each column represents a different vaccination group. Colors represent the percentage of the population in a given vaccination group to be vaccinated.

**Figure S24:**
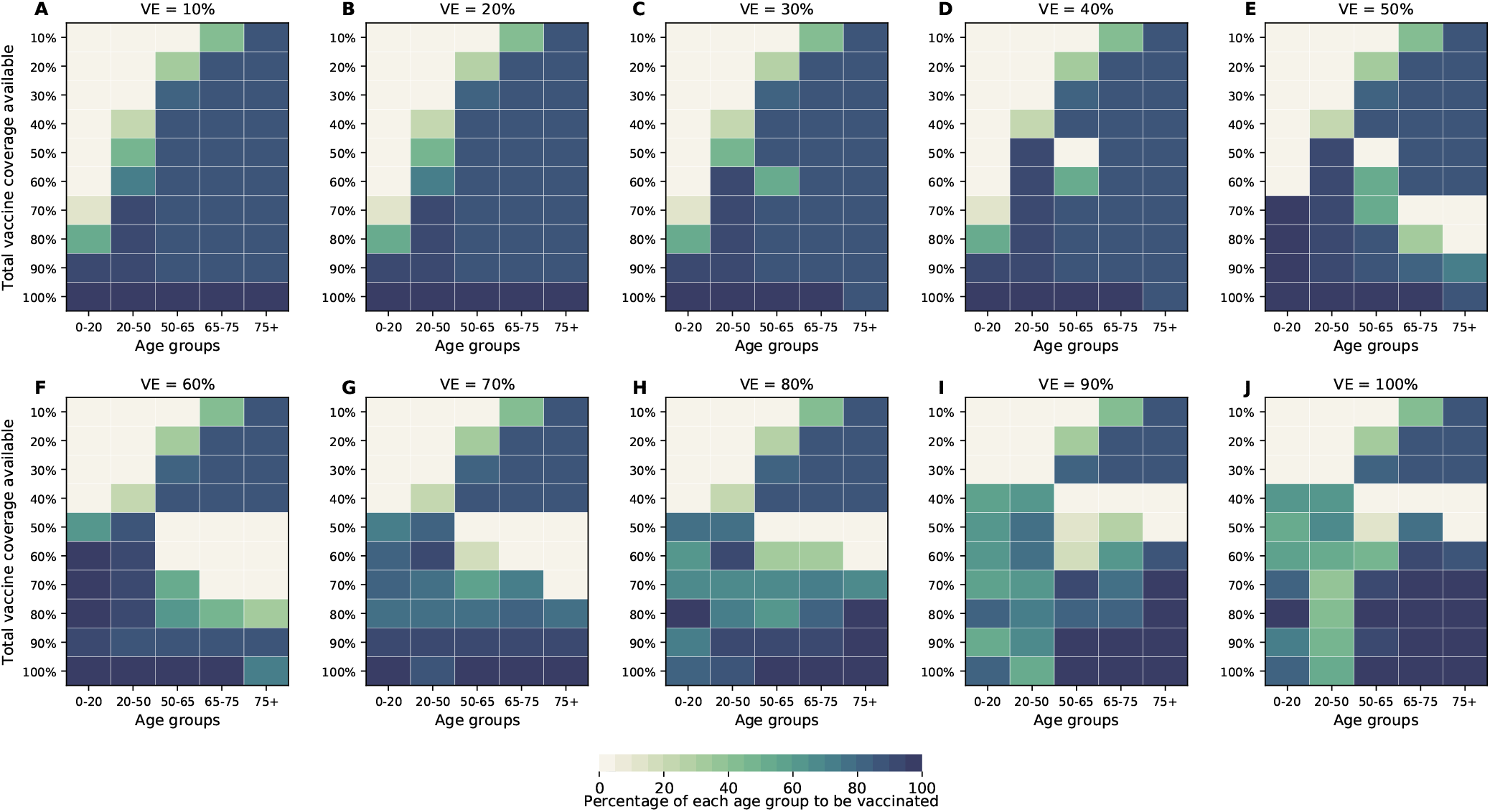
Optimal allocation strategies for minimizing the number of symptomatic infections for VE ranging from 10% (A) to 100% (J) in 10% increments assuming age-specific rates for symptomatic infection. For each plot, each row represents the total vaccination coverage available (percentage of the total population to be vaccinated) and each column represents a different vaccination group. Colors represent the percentage of the population in a given vaccination group to be vaccinated.

**Figure S25:**
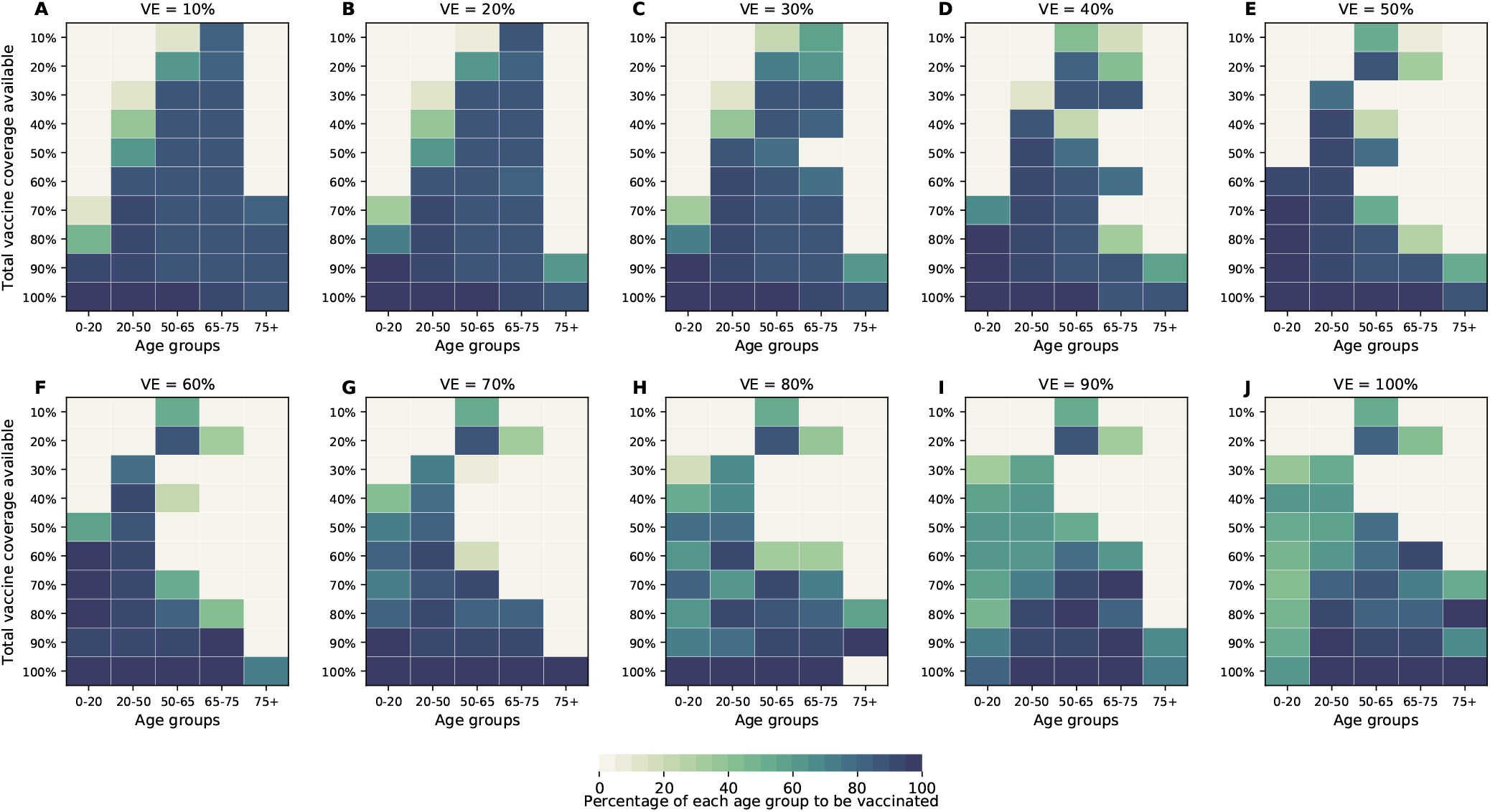
Optimal allocation strategies for minimizing the maximum number of non-ICU hospitalizations for VE ranging from 10% (A) to 100% (J) in 10% increments assuming age-specific rates for symptomatic infection. For each plot, each row represents the total vaccination coverage available (percentage of the total population to be vaccinated) and each column represents a different vaccination group. Colors represent the percentage of the population in a given vaccination group to be vaccinated.

**Figure S26:**
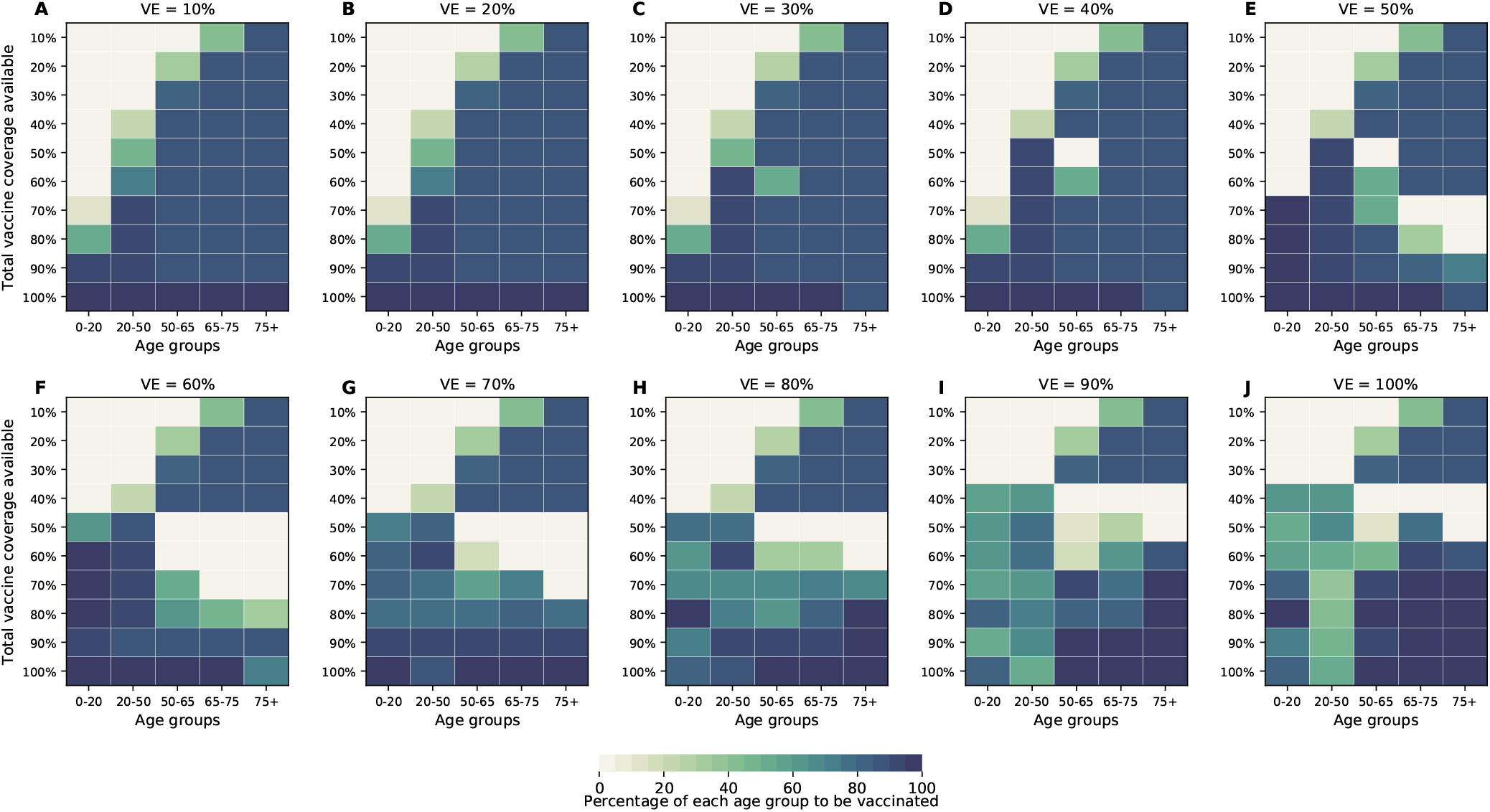
Optimal allocation strategies for minimizing the the maximum number of ICU hospitalizations for VE ranging from 10% (A) to 100% (J) in 10% increments assuming age-specific rates for symptomatic infection. For each plot, each row represents the total vaccination coverage available (percentage of the total population to be vaccinated) and each column represents a different vaccination group. Colors represent the percentage of the population in a given vaccination group to be vaccinated.

**Figure S27:**
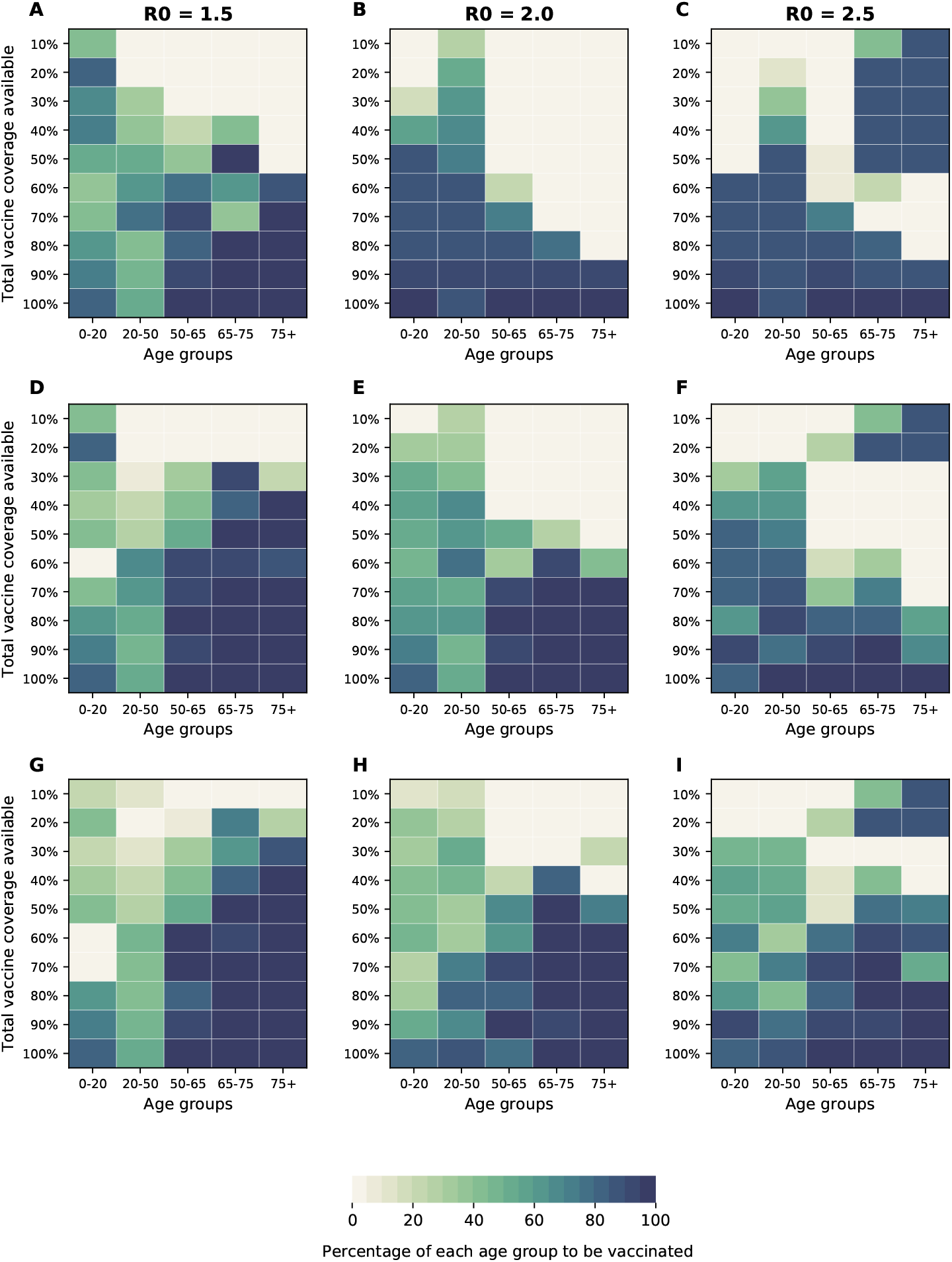
Optimal allocation strategies for minimizing maximum ICU hospitalizations for three different VE: 30% (A-C), 60%(D-F) and 90% (G-I) for three different values of *R*_0_ = 1.5, 2, and 2.5. For each plot, each row represents the total vaccination coverage available (percentage of the total population to be vaccinated) and each column represents a different vaccination group. Colors represent the percentage of the population in a given vaccination group to be vaccinated.

**Figure S28:**
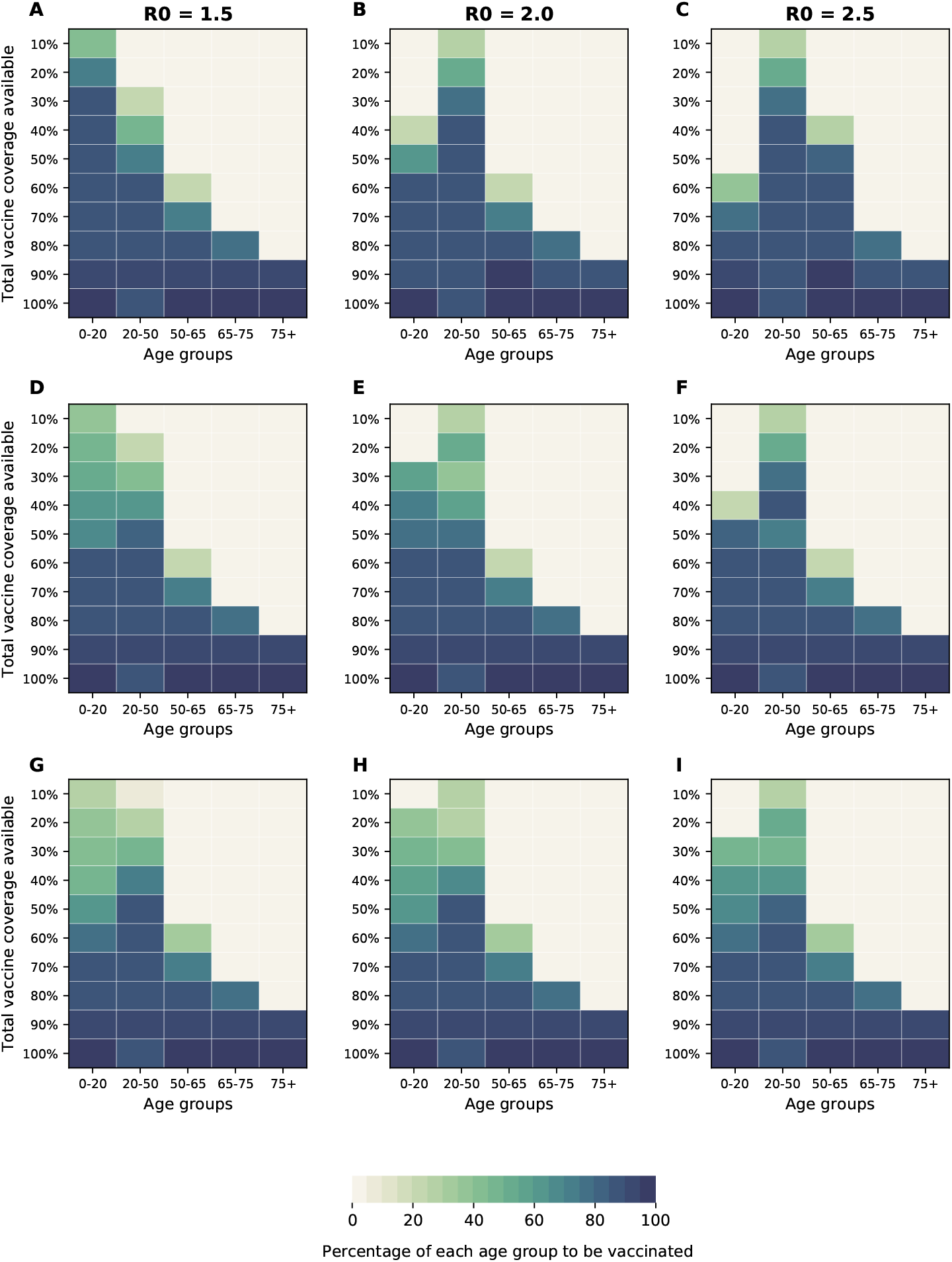
Optimal allocation strategies for minimizing symptomatic infections for three different VE: 30% (A-C), 60%(D-F) and 90% (G-I) for three different values of *R*_0_ = 1.5, 2, and 2.5. For each plot, each row represents the total vaccination coverage available (percentage of the total population to be vaccinated) and each column represents a different vaccination group. Colors represent the percentage of the population in a given vaccination group to be vaccinated.

**Figure S29:**
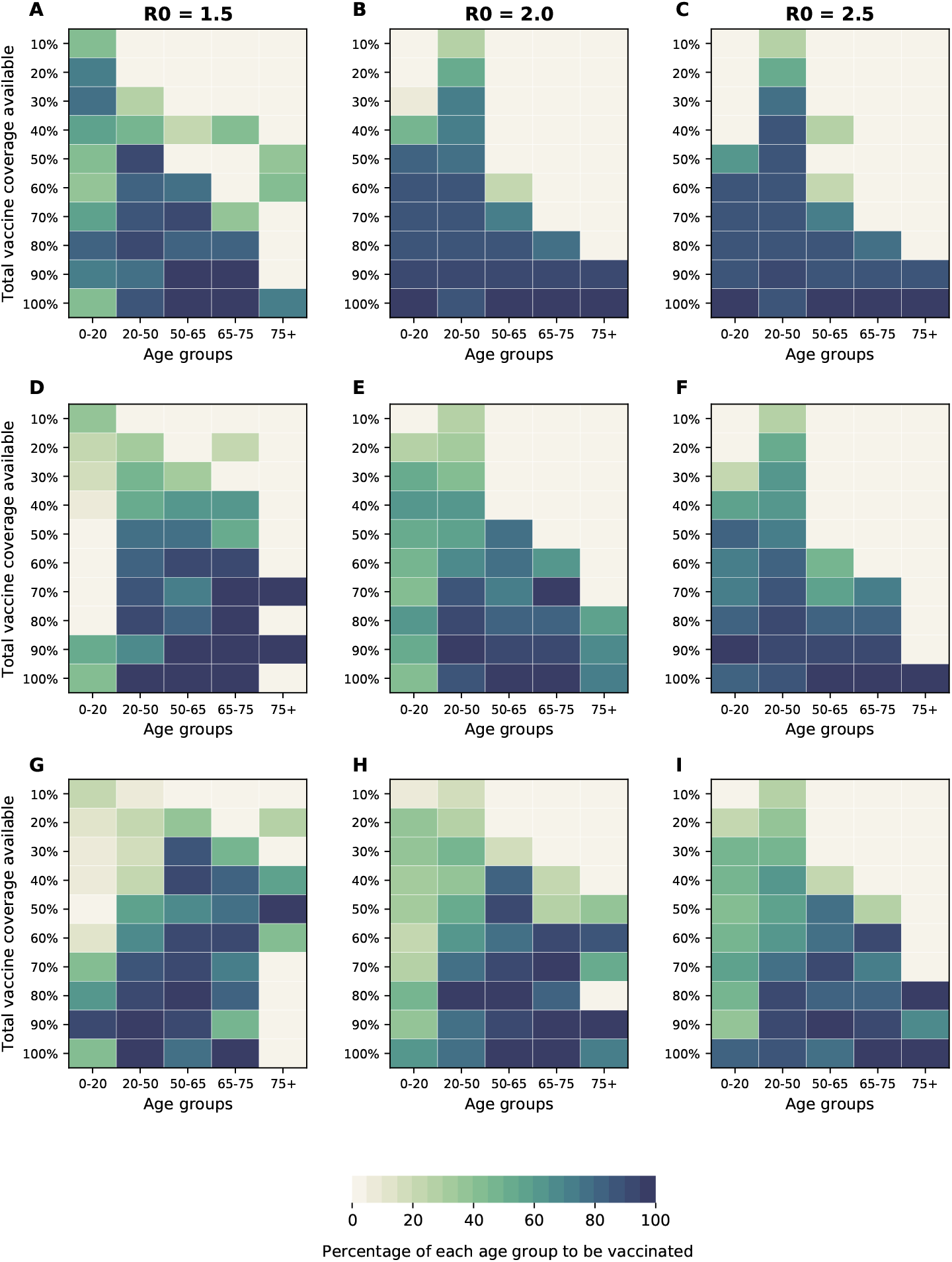
Optimal allocation strategies for minimizing maximum non-ICU hospitalizations for three different VE: 30% (A-C), 60%(D-F) and 90% (G-I) for three different values of *R*_0_ = 1.5, 2, and 2.5. For each plot, each row represents the total vaccination coverage available (percentage of the total population to be vaccinated) and each column represents a different vaccination group. Colors represent the percentage of the population in a given vaccination group to be vaccinated.

**Figure S30:**
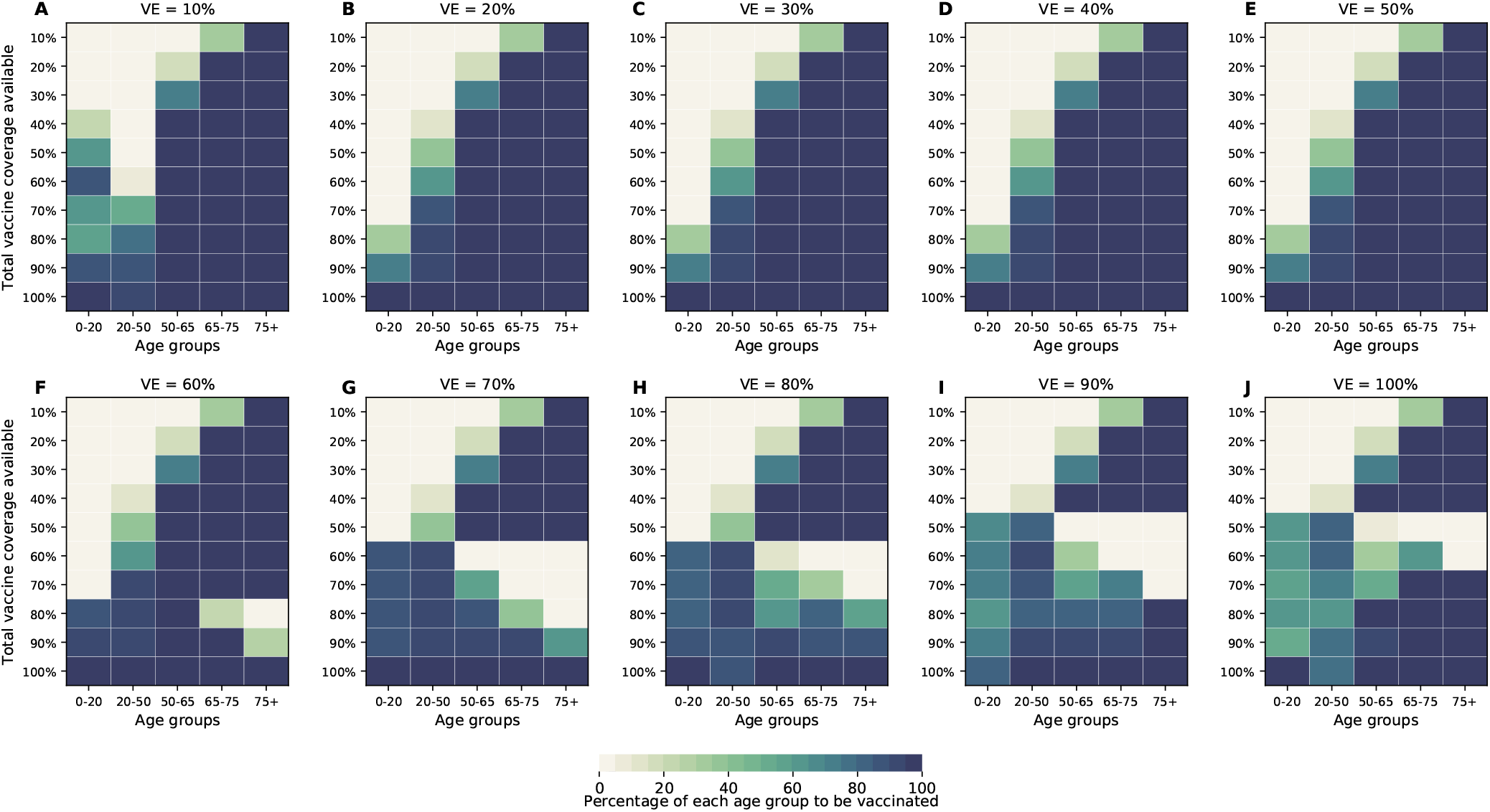
Optimal allocation strategies for minimizing the number of deaths for VE ranging from 10% (A) to 100% (J) in 10% increments assuming a different distribution of pre-existing immunity in the population (see Main for details). For each plot, each row represents the total vaccination coverage available (percentage of the total population to be vaccinated) and each column represents a different vaccination group. Colors represent the percentage of the population in a given vaccination group to be vaccinated.

**Figure S31:**
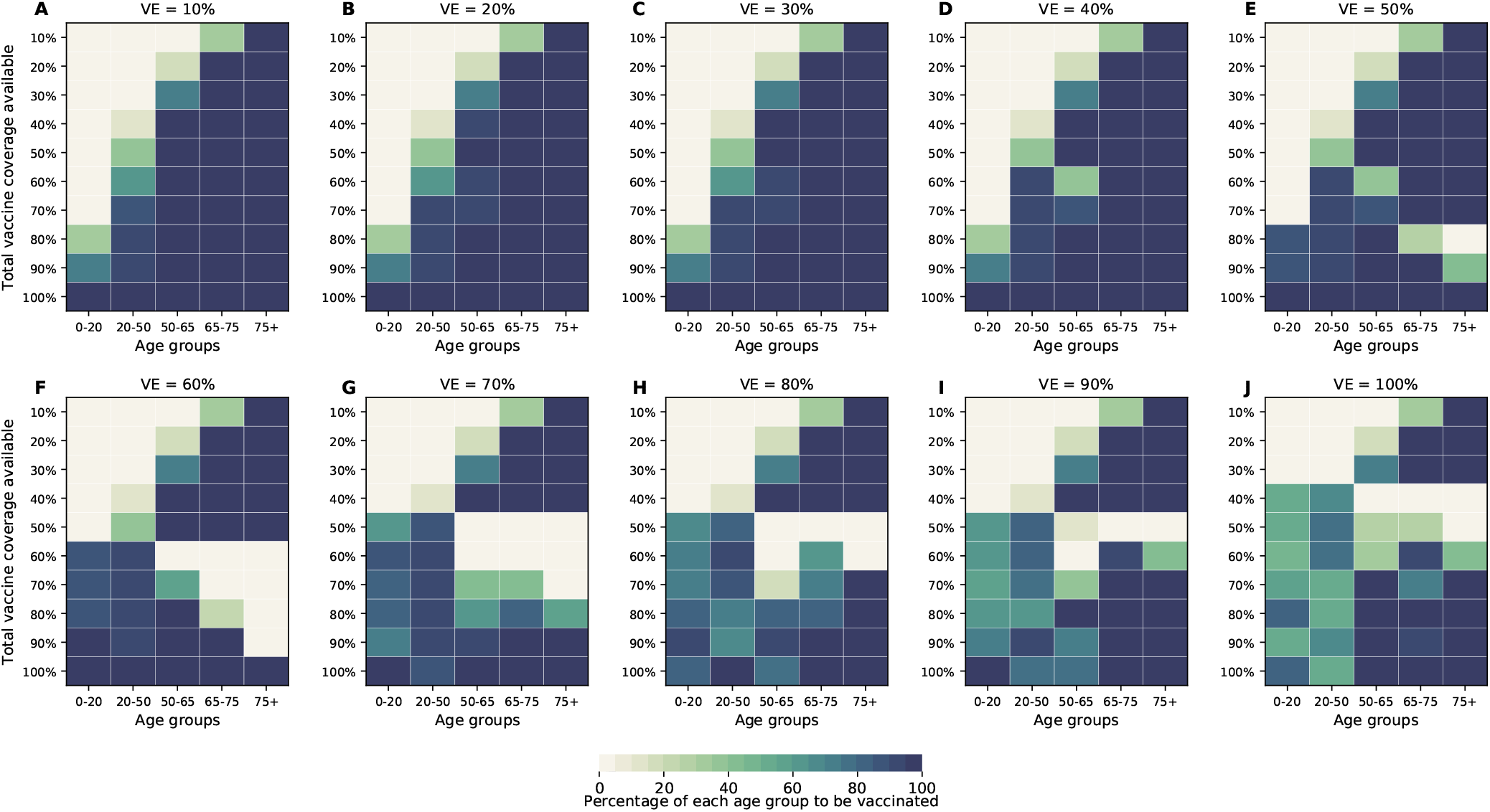
Optimal allocation strategies for minimizing the number of symptomatic infections for VE ranging from 10% (A) to 100% (J) in 10% increments assuming a different distribution of pre-existing immunity in the population (see Main for details). For each plot, each row represents the total vaccination coverage available (percentage of the total population to be vaccinated) and each column represents a different vaccination group. Colors represent the percentage of the population in a given vaccination group to be vaccinated.

**Figure S32:**
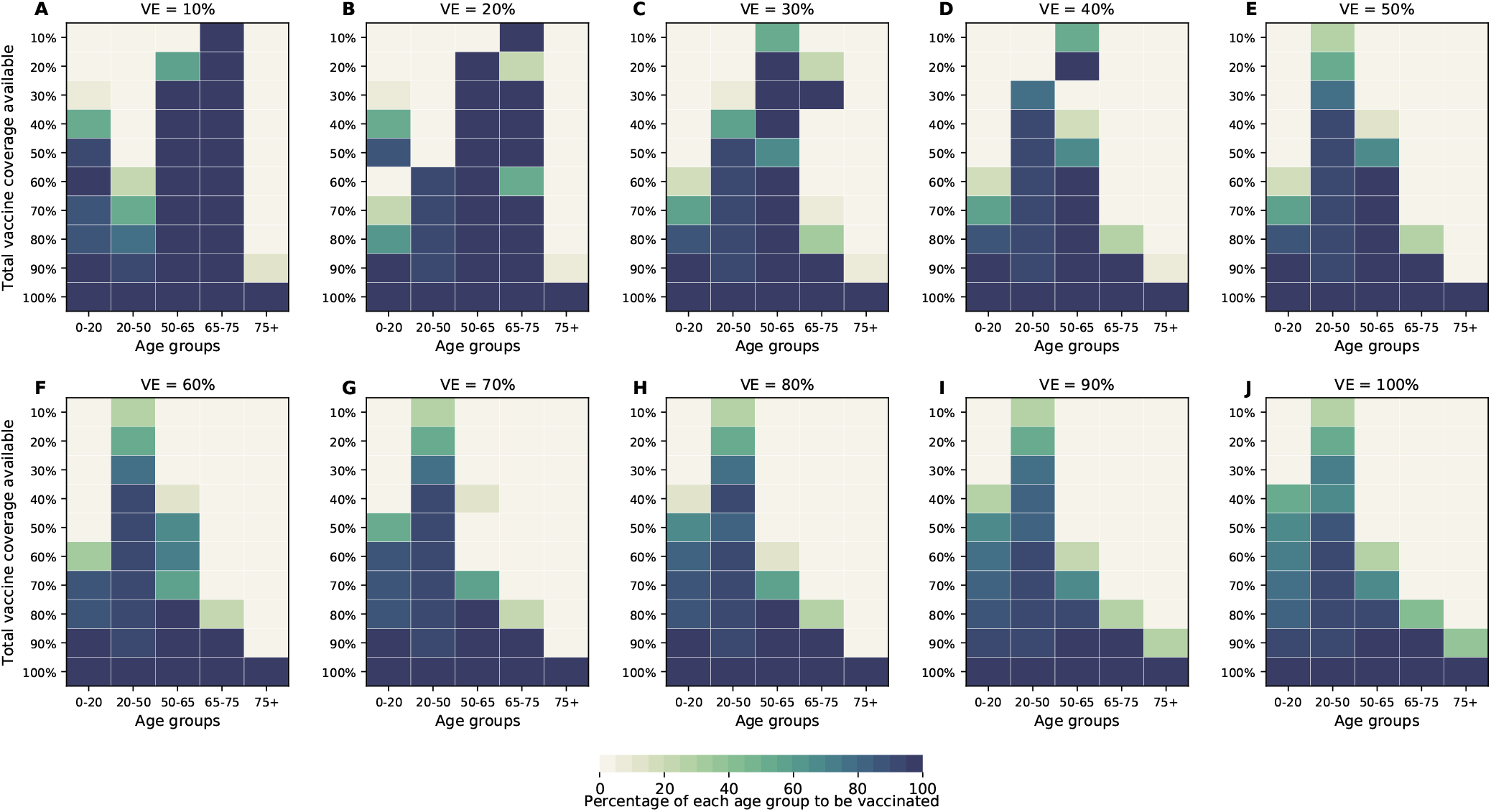
Optimal allocation strategies for minimizing the maximum number of non-ICU hospitalizations for VE ranging from 10% (A) to 100% (J) in 10% increments assuming a different distribution of pre-existing immunity in the population (see Main for details). For each plot, each row represents the total vaccination coverage available (percentage of the total population to be vaccinated) and each column represents a different vaccination group. Colors represent the percentage of the population in a given vaccination group to be vaccinated.

**Figure S33:**
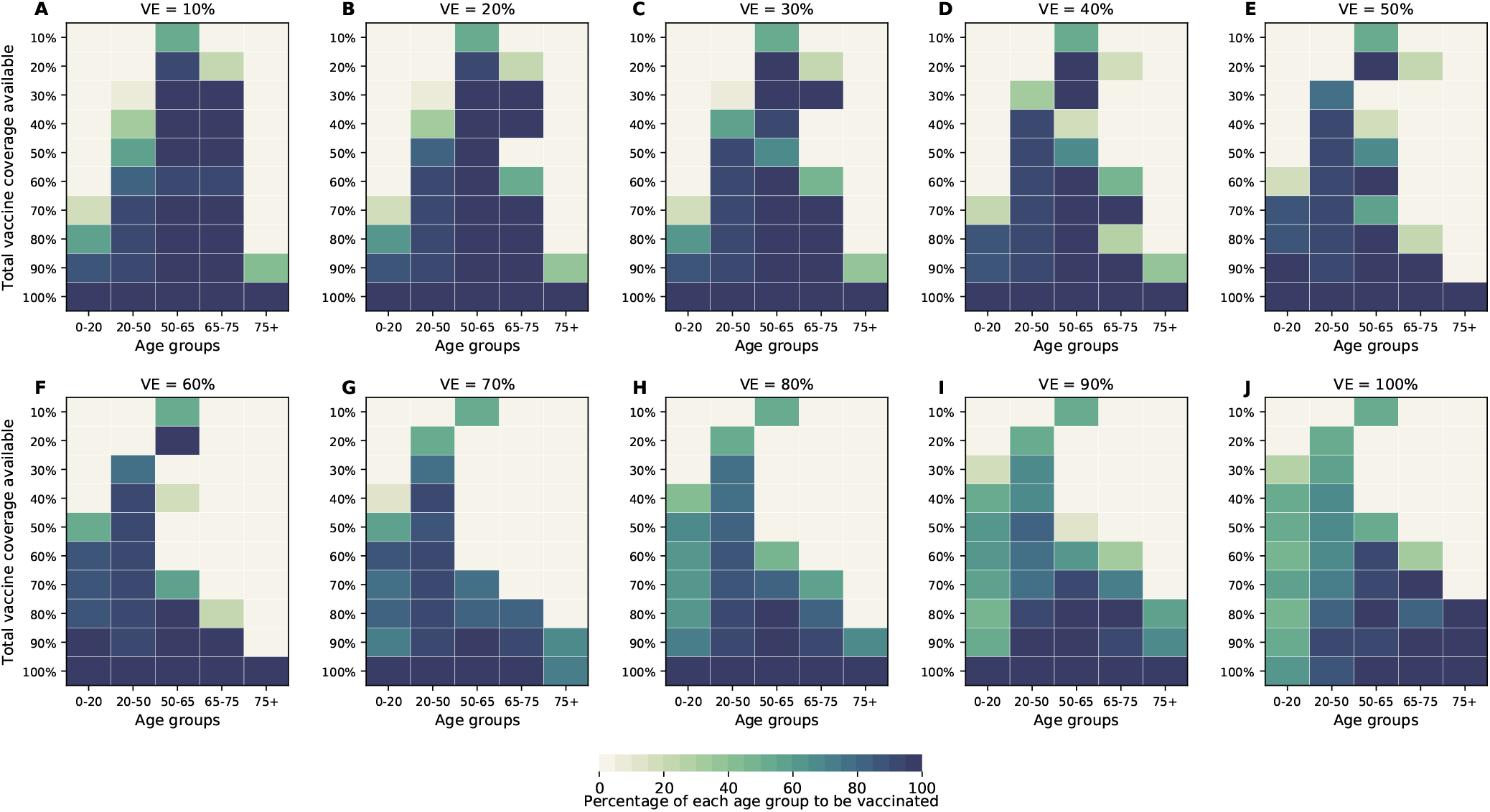
Optimal allocation strategies for minimizing the maximum number of ICU hospitalizations for VE ranging from 10% (A) to 100% (J) in 10% increments assuming a different distribution of pre-existing immunity in the population (see Main for details). For each plot, each row represents the total vaccination coverage available (percentage of the total population to be vaccinated) and each column represents a different vaccination group. Colors represent the percentage of the population in a given vaccination group to be vaccinated.

**Figure S34:**
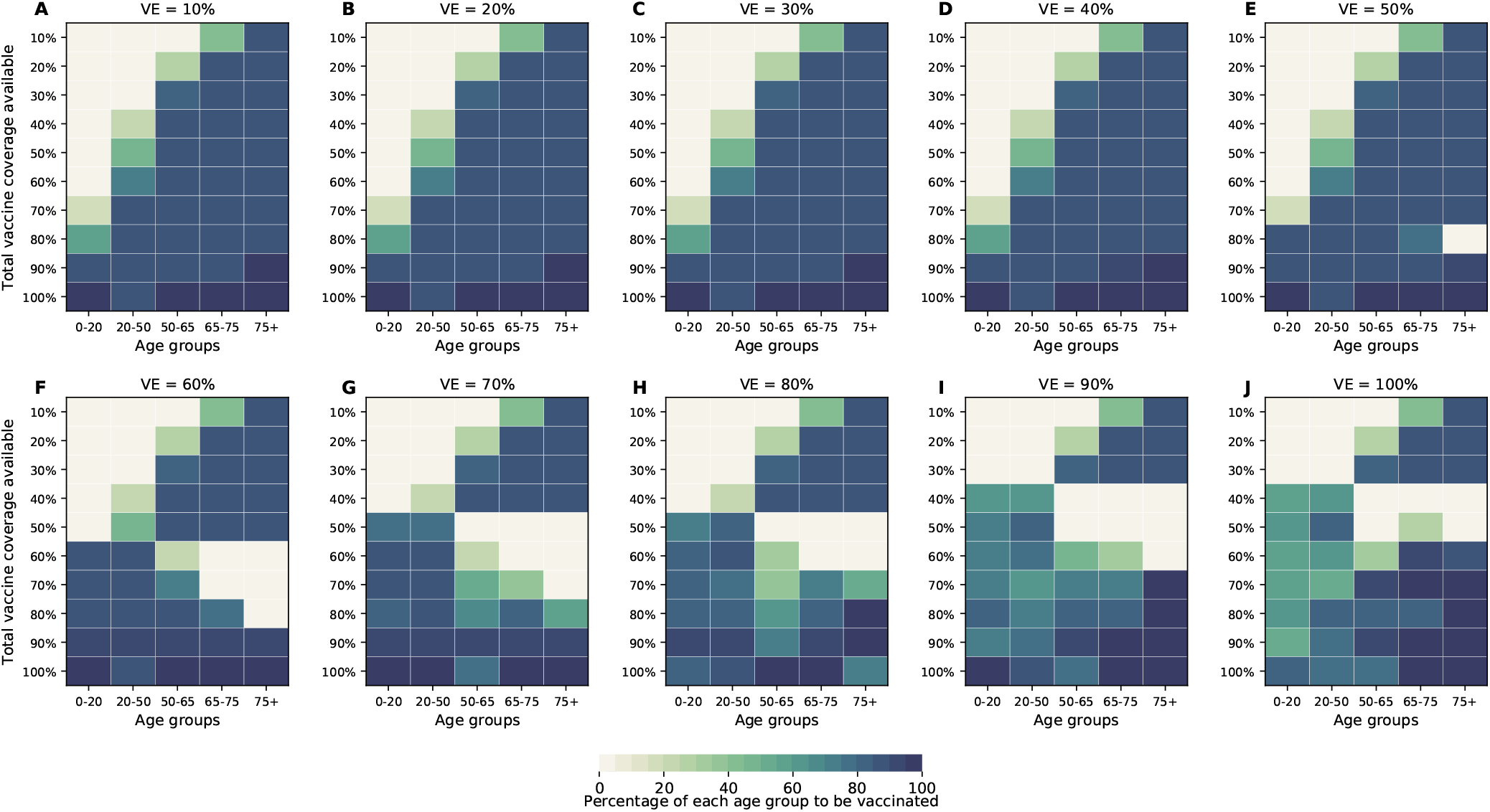
Optimal allocation strategies for minimizing the number of deaths for VE ranging from 10% (A) to 100% (J) in 10% increments assuming a latent period of 5 days. For each plot, each row represents the total vaccination coverage available (percentage of the total population to be vaccinated) and each column represents a different vaccination group. Colors represent the percentage of the population in a given vaccination group to be vaccinated.

**Figure S35:**
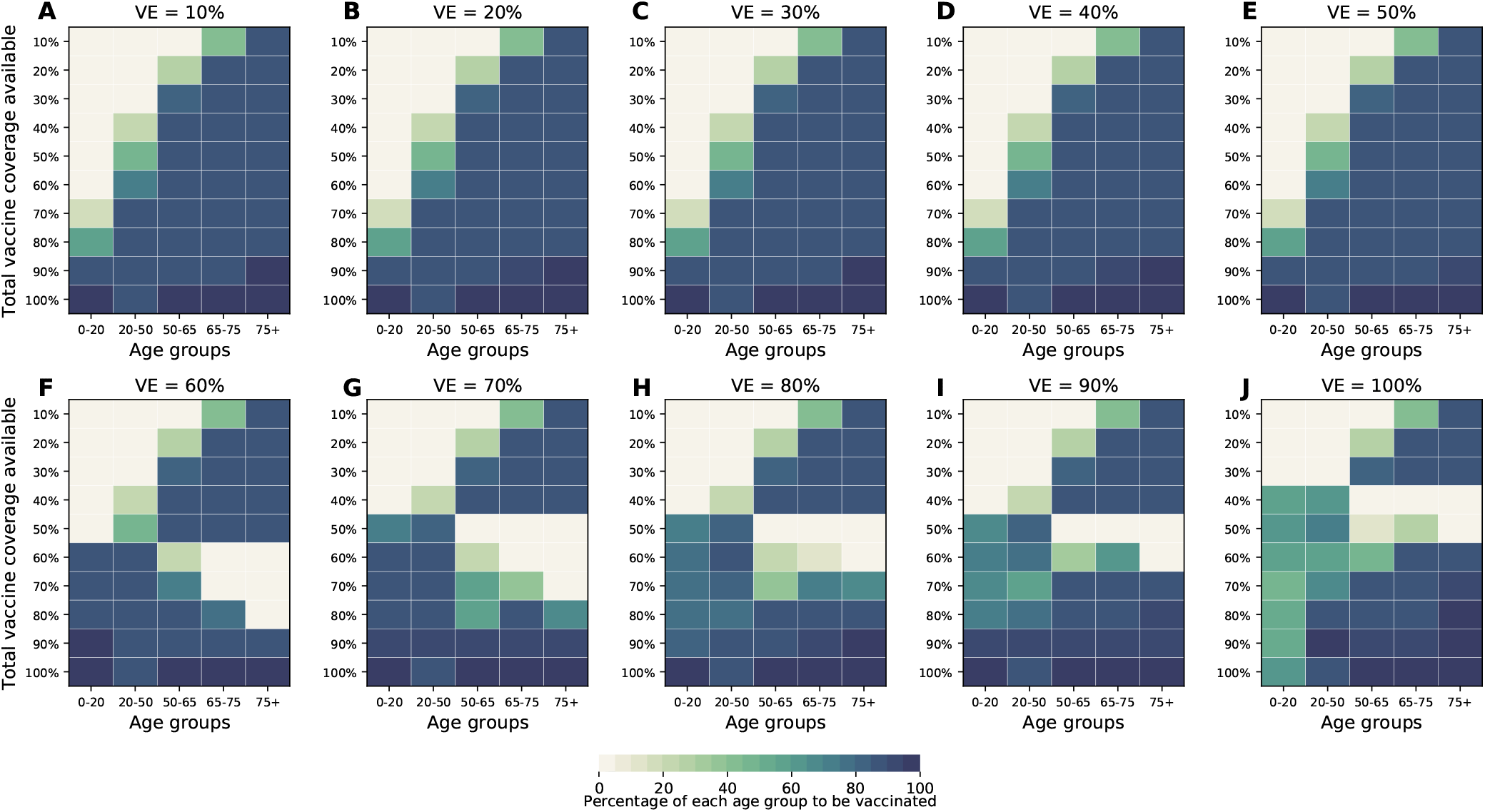
Optimal allocation strategies for minimizing deaths (with differential susceptibility) for VE ranging from 10% (A) to 100% (J) in 10% increments, with 10,000 infections at the start of simulation. For each plot, each row represents the total vaccination coverage available (percentage of the total population to be vaccinated) and each column represents a different vaccination group. Colors represent the percentage of the population in a given vaccination group to be vaccinated.

### Supplemental Tables

**Table S1:**
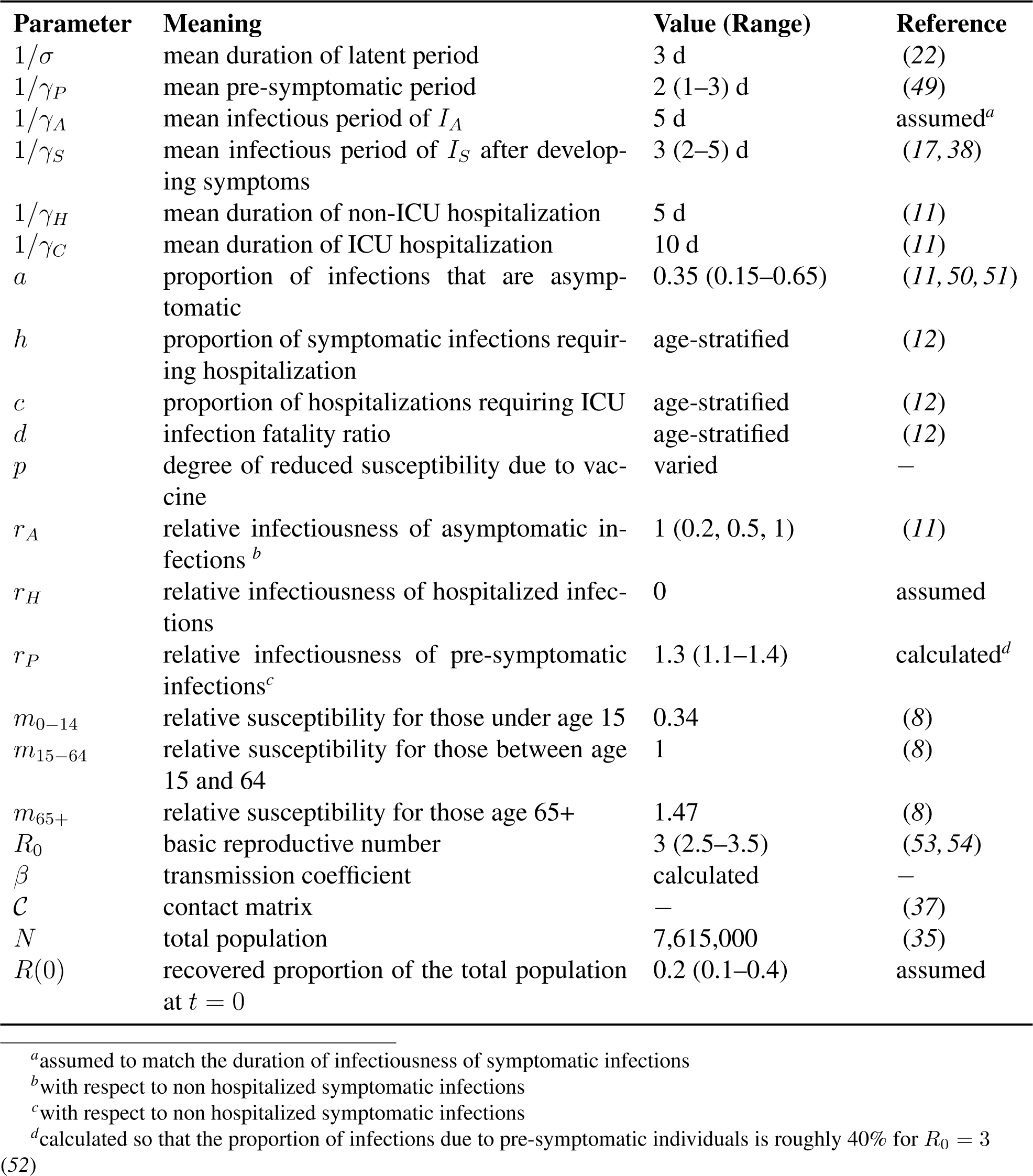
Description of parameters used in the model.

**Table S2:** Combinations of parameters used in the robustness analysis.

| $R_0$ | $a$ | $r_A$ | $r_P$ |
| --- | --- | --- | --- |
| 2.50 | 0.15 | 0.50 | 1.00 |
| 2.50 | 0.15 | 0.50 | 1.50 |
| 2.50 | 0.15 | 1.00 | 1.00 |
| 2.50 | 0.15 | 1.00 | 1.50 |
| 2.50 | 0.35 | 0.50 | 1.00 |
| 2.50 | 0.35 | 0.50 | 1.50 |
| 2.50 | 0.35 | 1.00 | 1.00 |
| 2.50 | 0.35 | 1.00 | 1.50 |
| 2.50 | 0.55 | 0.50 | 1.00 |
| 2.50 | 0.55 | 0.50 | 1.50 |
| 2.50 | 0.55 | 1.00 | 1.00 |
| 2.50 | 0.55 | 1.00 | 1.50 |
| 3.00 | 0.15 | 0.50 | 1.00 |
| 3.00 | 0.15 | 0.50 | 1.50 |
| 3.00 | 0.15 | 1.00 | 1.00 |
| 3.00 | 0.15 | 1.00 | 1.50 |
| 3.00 | 0.35 | 0.50 | 1.00 |
| 3.00 | 0.35 | 0.50 | 1.50 |
| 3.00 | 0.35 | 1.00 | 1.00 |
| 3.00 | 0.35 | 1.00 | 1.50 |
| 3.00 | 0.55 | 0.50 | 1.00 |
| 3.00 | 0.55 | 0.50 | 1.50 |
| 3.00 | 0.55 | 1.00 | 1.00 |
| 3.00 | 0.55 | 1.00 | 1.50 |
| 3.50 | 0.15 | 0.50 | 1.00 |
| 3.50 | 0.15 | 0.50 | 1.50 |
| 3.50 | 0.15 | 1.00 | 1.00 |
| 3.50 | 0.15 | 1.00 | 1.50 |
| 3.50 | 0.35 | 0.50 | 1.00 |
| 3.50 | 0.35 | 0.50 | 1.50 |
| 3.50 | 0.35 | 1.00 | 1.00 |
| 3.50 | 0.35 | 1.00 | 1.50 |
| 3.50 | 0.55 | 0.50 | 1.00 |
| 3.50 | 0.55 | 0.50 | 1.50 |
| 3.50 | 0.55 | 1.00 | 1.00 |
| 3.50 | 0.55 | 1.00 | 1.50 |

### Supplemental Files

**SF1:** Robustness analysis for the optimization strategies when minimizing symptomatic infections. Each page of the document represents one VE. Within each page, each subplot represents the optimal allocation strategy for that particular VE and for the combination of parameters given in the row of Table S2 indicated in the subtitle.

**SF2:** Robustness analysis for the optimization strategies when minimizing total deaths. Each page of the document represents one VE. Within each page, each subplot represents the optimal allocation strategy for that particular VE and for the combination of parameters given in the row of Table S2 indicated in the subtitle.

**SF3:** Robustness analysis for the optimization strategies when minimizing peak non-ICU hospitalizations. Each page of the document represents one VE. Within each page, each subplot represents the optimal allocation strategy for that particular VE and for the combination of parameters given in the row of Table S2 indicated in the subtitle.

**SF4:** Robustness analysis for the optimization strategies when minimizing peak ICU hospitalizations. Each page of the document represents one VE. Within each page, each subplot represents the optimal allocation strategy for that particular VE and for the combination of parameters given in the row of Table S2 indicated in the subtitle.

